# Rapid protocols to support Covid-19 clinical diagnosis based on hematological parameters

**DOI:** 10.1101/2021.06.21.21259252

**Authors:** Juliana Carneiro Gomes, Valter Augusto de Freitas Barbosa, Maíra Araújo de Santana, Clarisse Lins de Lima, Raquel Bezerra Calado, Cláudio Roberto Bertoldo Júnior, Jeniffer Emidio de Almeida Albuquerque, Rodrigo Gomes de Souza, Ricardo Juarez Escorel de Araújo, Giselle Machado Magalhães Moreno, Luiz Alberto Lira Soares, Luiz Alberto Reis Mattos Júnior, Ricardo Emmanuel de Souza, Wellington Pinheiro dos Santos

## Abstract

**Purpose:** In December 2019, the Covid-19 pandemic began in the world. To reduce mortality, in addiction to mass vaccination, it is necessary to massify and accelerate clinical diagnosis, as well as creating new ways of monitoring patients that can help in the construction of specific treatments for the disease.

**Objective:** In this work, we propose rapid protocols for clinical diagnosis of Covid-19 through the automatic analysis of hematological parameters using Evolutionary Computing and Machine Learning. These hematological parameters are obtained from blood tests common in clinical practice.

**Method:** We investigated the best classifier architectures. Then, we applied the particle swarm optimization algorithm (PSO) to select the most relevant attributes: serum glucose, troponin, partial thromboplastin time, ferritin, D-dimer, lactic dehydrogenase, and indirect bilirubin. Finally, we used decision trees to build four rapid protocols for Covid-19 clinical diagnosis.

**Results:** We developed a web system for Covid-19 diagnosis support. Using a 100-tree Random Forest, we obtained results for accuracy, sensitivity and specificity superior to 99

**Conclusion:** By using a reduced set of hematological parameters common in clinical practice, it was possible to achieve results of accuracy, sensitivity and specificity comparable to those obtained with RT-PCR. It was also possible to automatically generate clinical decision protocols, allowing relatively accurate clinical diagnosis even without the aid of the web decision support system.

## 1 Introduction

### 1.1 Motivation and problem characterization

The world that emerged after the Second World War was marked by a rapid process of globalization. An interconnected world has emerged, totally connected both by advanced means of transport, such as airplanes and ships, and by means of information and communication technologies. Trade necessarily integrates nations and intensifies the movement of people across the globe. However, from the point of view of Epidemiology, a fully connected world is also a world more susceptible to several threats, including health threats, such as epidemics and pandemics [28, 85]. The pathways through which international trade flows are also the pathways used by infectious disease vectors.

Coronaviruses are viruses of the Coronaviridae family, known since the 1960s, responsible for respiratory infections in humans and animals. In 2002, there was an outbreak in China of Severe Acute Respiratory Syndrome (SARS), a viral respiratory disease of zoonotic origin transmitted by the SARS-CoV coronavirus. SARS-CoV has spread rapidly to more than a dozen countries in North America, South America, Europe and Asia, infecting more than 8,000 people and causing about 800 deaths. The SARS epidemic was controlled in 2003, but the disease has become endemic in many countries [35]. In 2012, the Middle East Respiratory Syndrome (MERS) appeared, a respiratory disease transmitted by the MERS-CoV coronavirus. This coronavirus was unknown as an agent of human disease until its identification, initially in Saudi Arabia and, later, in other countries in the Middle East, in Europe and Africa [3, 20, 70]. MERS-CoV has its origins in nature in camels and bats [3, 20, 70]. Confirmed cases of MERS developed into respiratory disease with fever, cough and dyspnea. Most patients had pneumonia. Some immunodepressed patients initially had fever and diarrhea and, in some cases, pneumonia. The most frequent complications in the evolution of the cases were respiratory failure, adult respiratory distress syndrome, septic shock, renal failure, disseminated intravascular coagulation, and pericarditis [3, 20, 70]. About 30% of patients confirmed with MERS-CoV died [2, 8].

In December 2019, in the city of Wuhan, China, the most critical outbreak in the last hundred years began: Coronavirus Disease 2019 (Covid-19), transmitted by the SARS-CoV-2 virus, a virus of zoonotic origin until then unknown, present in bats and pangolins [5, 21, 31, 89, 111]. SARS-CoV-2, when compared to its predecessors, proved to be much more resistant and infectious. The most common symptoms are fever, dry cough and tiredness [21, 31, 42, 89, 111]. Pain and discomfort, sore throat, diarrhea, conjunctivitis, headache, loss of taste or smell, rash on the skin, or discoloration of the fingers or toes may also appear [15, 21, 78, 89, 107, 111]. Its severe symptoms are difficulty in breathing or shortness of breath, pain or pressure in the chest, and loss of speech or movement [15, 21, 31, 42, 78, 89, 107, 111]. Despite the lower lethality, the virus spreads very quickly, producing a large volume of deaths and leaving sequels that are often permanent [15, 31, 78, 107]. Due to their high rate of contagion, public health system resources are rapidly depleted [31]. The Covid-19 pandemic is one of the biggest health crises in decades. In March 2021, SARS-CoV had already infected almost 130 million people, more than 70 million of whom recovered, while almost 3 million died [75]. In this context, it is not enough to invest in the opening of new hospital beds for the treatment of patients. It is necessary to have tests that guarantee fast and reliable diagnoses; specific treatments to decrease the lethality of the disease; efficient and low-cost vaccines applied to a considerable portion of the population; and social isolation and quarantine policies to seek to control the disease vector while vaccines and specific treatments are not available for Covid-19.

Accurate diagnosis plays an important role against Covid-19. The test established as the gold standard for the diagnosis of Covid-19 is the Reverse Transcription Polymerase Chain Reaction (RT-PCR), used to search for the presence of the SARS-CoV-2 virus, translated from RNA into DNA, in samples of saliva and human secretions [36]. However, the RT-PCR process can take hours or even days, considering both the volume of tests required and the logistics of transporting the samples, given the pandemic situation [36]. Late diagnosis can result in late patient care, which can make recovery difficult. In addition, non-isolated infected people can spread the virus.

The most common rapid tests are based on the identification of serological evidence of the presence of the virus as antibodies or antigens. Therefore, these tests are nonspecific because they do not detect the presence of the virus directly. Given that other viral infections can produce responses of antibodies and similar antigens, such as common flu, the performance of the tests depends on other factors, such as the time of onset of the disease and the viral concentration in the sample of interest [60, 74]. For example, tests based on IgM/IgG antibodies performed in the first week of the disease have 18.8% sensitivity and 77.8% specificity [67]. However, in the second week, IgG/IgM tests are performed with 100% sensitivity and 50% specificity [67]. Therefore, when the viral load is high, the IgG/IgM tests reach high sensitivities and specificities. But in these cases, the disease is at advanced levels [46, 50]. That is why the World Health Organization does not recommend the use of this type of test for clinical decision-making [74].

Several studies have sought to highlight the nature of Covid-19 as a disease that mainly affects the cardiovascular system [16, 37, 38, 65, 98, 113]. Coronaviruses, such as SARS-CoV and SARS-CoV-2, have the angiotensin-converting zinc metallopeptidase 2 (ACE2), an enzyme present in the cell membranes of the arteries, heart, lungs and other organs as a functional receptor. ACE2 is involved in cardiac function, hypertension and diabetes [102]. The MERS-CoV and SARS-CoV coronaviruses can cause acute myocarditis and heart failure [113]. Some of the impacts of coronaviruses on the cardiovascular system are increased blood pressure and increased levels of troponin I (hs-cTnI) [113]. Covid-19 patients may also develop lymphopenia, i.e. low level of lymphocytes [37, 65, 98]; and leukopenia, i.e. few leukocytes. Covid-19 patients may also experience decreased hemoglobin levels, Absolute Lymphocyte Count (ALC) and Absolute Monocyte Count (AMC) [37]. Patients who have developed severe forms of the disease have significantly higher levels of Interleukin-6 and D-dimer than patients who have developed a moderate form of Covid-19 [38]. Therefore, considering that Covid-19 is a disease that affects blood parameters, hematological tests can be used to help diagnose the disease.

Given that the result of the RT-PCR test can take hours or even days, given the pandemic situation that increases the demand for tests with the high speed of contamination, clinical diagnosis assumes a fundamental role in determining the treatment and the type of care correct for mild, moderate and, mainly, severe cases. In this sense, the observation of complementary parameters, such as the hematological parameters obtained from common tests in clinical practice, takes on a very important role. Several works have been using machine learning techniques to diagnosis diseases by analyzing hematological parameters automatically [5, 45, 69, 100]. Blood tests are commonly used during medical screening. The most common blood tests, like complete blood count, bilirubin, serum gligose, c-reactive protein, urea, and others, are easily available at relatively low-cost compared with other diagnosis methods. Therefore, intelligent systems can be used to automatically analyze hematological parameters and use them to support Covid-19’s clinical diagnosis and to suggest appropriate patient care [5].

In this work we propose the following rapid protocols to support the clinical diagnosis of Covid-19 based on hematological parameters: (1) rapid diagnosis based on automatic analysis by an intelligent web system based on Particle Swarm Optimization and Random Forests; (2) rapid diagnosis by statistical inference by decision trees. In the first approach, an intelligent web system was developed to support decision making. This system was put into operation in the city of Paudalho, Brazil, to support the clinical diagnosis and assessment of the severity of the disease in patients admitted to intensive and semi-intensive care units, in the year 2020. In the second approach, we used decision trees to infer that hematological parameters would be statistically more relevant for clinical diagnosis without the support of an intelligent system. The most relevant parameters were ferritin and prothrombin partial time. The results of general accuracy, sensitivity and specificity were quite high for both approaches, demonstrating that rapid diagnosis is possible using only well-known and low-cost clinical exams, with the potential support of an intelligent decision support system. The high correlation between ferritin, prothrombin time and a positive diagnosis for Covid-19 in symptomatic patients also points to the need for more research on treatments to combat early clotting and the main symptoms of the disease.

### 1.2 Related works

Several studies emphasize the importance of hematological parameters to support Covid-19 clinical diagnosis. Some of them point to the relevance of using hematological analysis as an indicative of the severity degree of Covid-19. Fan et al. [37] analyzed hematological parameters of 69 patients with Covid-19. The study was conducted with subjects from the National Center for Infectious Diseases (NCID) in Singapore. 65 of these patients underwent complete blood count (CBC) on the day of admission. 13.4% of patients needed intensive care unit (ICU) care, especially the elderly. During the first exams, 19 patients had leukopenia (few white blood cells) and 24 had lymphopenia (low level of lymphocytes in the blood), with 5 cases classified as severe (Absolute Lymphocyte Count (ALC) *<* 0.5 × 10^9^*/L*). The study also pointed out that patients who needed to be admitted to the ICU had lower ALC and a higher rate of Lactate Dehydrogenase (LDH). These data indicated that monitoring these parameters can help to identify patients who need assistance in the ICU. The authors found that the patients who were in the ICU had a significant decrease in their hemoglobin levels, ALC and Absolute Monocyte Count (AMC) levels, when compared to the non-ICU group. ICU patients also tend to neuthophilia. The platelet count did not prove to be a factor for discrimination between the type of hospitalization.

The work from Tan et al. [98] also assessed the complete blood count of patients. They used data from both cured patients and patients who died from Covid-19. The main purpose of the study was to obtain key indicators of disease progression, in order to support future clinical management decisions. In the case of patients who died, blood tests were continuously monitored throughout the treatment process. Similar to the previous study, the authors observed lymphopenia in this group. Based on this, the study then outlined a model (Time-LYM% model, TLM) for classifying disease severity and predicting prognosis. Thus, the blood lymphocyte percentage (LYM%) was divided into two cases, considering the first 10-12 days of symptoms: LYM% *>* 20% are classified as moderate cases and with a high chance of recovery. LYM% *<* 20% are classified as severe cases. In a second exam, 17-18 days after the first symptoms, patients with LYM% *>* 20% are recovering, patients with 5 *<*LYM% *<* 20% are in danger, and LYM% *<* 5% are in critical condition. In order to validate the model, the authors evaluated 90 patients with Covid-19. The consistency between Guideline and TLM-based disease classification was verified using kappa statistic (Kappa = 0.48). These results indicate that probably LYM% should be used together with other parameters for a better evaluation of Covid-19.

Gao et al. [38] assessed hematological characteristics of 43 patients at Fuyang Second People’s Hospital. The patients had diagnosis confirmed by the Covid-19 ground truth test, the fluorescent reverse transcription-polymerase chain reaction (RT-PCR). They were divided into two groups: the moderate group with 28 patients, and the severe group with 15 patients. The groups have no significant difference in age and sex. The blood tests observed were: Routine blood tests (white blood cell [WBC] count, lymphocyte count [LYM], mononuclear count [MONO], neutrophils count [NEU]) were performed on the blood samples. Blood biochemistry parameters (aspartate aminotransferase [AST], alanine aminotransferase [ALT], glucose [GLU], urea, creatinine [Cr], cystatin [Cys-c], uric acid [UA], and C-reactive protein [CRP]); Coagulation functions (D-dimer [d-D], thrombin time [TT], prothrombin time [PT], fibrinogen [FIB], activated partial thromboplastin time [APTT]); rocalcitonin (PCT); and Interleukin-6 (IL-6). Using statistical tests, the study noted that the levels of GLU, CRP, IL-6, TT, FIB, and d-D were significantly higher in the severe group than in the mild group. Performing this analysis with ROC curves, the authors pointed out that the best indicators for predicting severity were IL-6 and d-D combined, with AUC of 0.840. The combination also achieved specificity of 96.4% and sensitivity of 93.3%, using tandem and parallel testing, respectively. These results indicate that patients with severe conditions would have abnormal coagulation.

Liu et al. [65] reported that lymphopenia and inflammatory cytokine storm are abnormalities commonly found in other infections caused by coronavirus, such as SARS-Cov and MERS-Cov. With that in mind, they studied 40 patients diagnosed with Covid-19 confirmed by throat-swab specimens analyzed with RT-PCR. The patients were treated at Wuhan Union Hospital in January, 2020. The information provided was: epidemiological, demographic, clinical manifestations and laboratory tests. Similar to the previous study, patients were divided into two groups: mild patients, with symptoms such as epidemiological history, fever or respiratory symptoms, and abnormalities in imaging tests; the second group with severe patients, patients should additionally have symptoms such as shortness of breath, oxygen saturation *<* 93%, respiratory *>* 30 times/min, or PaO_2_*/*FiO_2_ *<* 300 mmHg. 27 patients were classified in the first group, while 13 were classified in the second. The study reported that levels of fibrinogen, D-dimer, total bilirubin, aspartate transaminase, alanine transaminase, lactate dehydrogenase, creatine kinase, C-reactive protein (CRP), ferritin and serum amyloid A protein were significantly higher in the severe group. Futhermore, most severe patients presented lymphopenia, that can be related to the significantly decreased absolute counts of T cells, especially CD8+ T cells, while white blood cells and neutrophils counts were higher.

These studies have pointed out that hematological parameters can be indicators of the risk factors and degree of severity of Covid-19. The identification of these parameters can be essential to optimize clinical care for each group of patients. In this sense, the development of intelligent systems based on blood tests is useful. Faced with the pandemic scenario, in which most hospitals are full, decision support systems can facilitate clinical management. Thus, it can increase the assertiveness in the treatment for each case and, consequently, the number of lives saved.

Gunčar et al. [45] proposed a system based on machine learning for analyzing blood tests and predicting hematological diseases. Their database was acquired between the years 2005 and 2015 at the University Medical Center of Ljubljana. In this case, 43 diseases and 181 parameters or features were selected to generate a first model (SBA-HEM181). In addition to it, a second model with 61 parameters was also developed (SBA-HEM061). The selection criteria was based on the frequency of use. Regarding the missing values (about 75%), the authors filled in with median values for each attribute. As classification methods, the authors tested classic approaches, such as Support Vector Machines, Naive Bayes and Random Forest. The simulations were repeated 10 times using 10-fold cross validation. Finally, the models SBA-HEM181 and SBA-HEM061 reached an accuracy of 57% considering all the diseases chosen. By restricting the prediction to five classes, the systems achieved an accuracy of 88% and 86%, respectively. These results were achieved when using Random Forest for classification. This study also pointed to the possibility of effectively detecting diseases through blood tests using classic intelligent classifiers.

Barbosa et al. [5] proposed an intelligent system to support Covid-19 diagnosis using blood exams. After testing several machine learning methods, they achieved high classification performance using Bayes network as classifier. The intelligent system was built using a 5,644-subjects public database from the Hospital Israelita Albert Einstein, Brazil [53], and showed average accuracy of 95.2% ± 0.7, with kappa index of 0.90 ± 0.01, sensitivity of 0.968 ± 0.007, precision of 0.94 ± 0.01 and specificity of 0.94 ± 0.01. The authors were able to minimize costs by selecting only 24 blood tests from the set of 107 available exams. Although the study achieved good results, the set of selected tests did not considered all the exams indicated by subsequent recommendations from the Brazilian Ministry of Health when dealing with Covid-19 patients [13].

Similarly, Soares et al. [95] use a method based on artificial intelligence to identify Covid-19 through blood tests. As Barbosa et al. [5] did, they used the database from the Hospital Israelita Albert Einstein. However, since the database has many missing data, they chose to include only the subjects that had most of the data. This procedure reduced the dataset from 5,644 samples to 599 samples. By using Support Vector Machines as a classifier and SMOTEBoost technique to perform oversampling, they achieved average specificity of 85.98%, Negative Predictive Value (NPV) of 94.92%, average sensitivity of 70.25% and Positive Predictive Value (PPV) of 44.96%.

These intelligent systems based on blood tests may play an important role in the process of diagnosing Covid-19, since many studies are confirming evidences of how this disease affects hematological parameters. Positive cases can be referred to further highly sensitive testing such as RT-PCR, computerized tomography scans and radiography [42, 43].

Negri et al. [73] have been using elevated D-dimer as a predictor of severity and mortality in Covid-19 patients. They also observed that heparin use during in-hospital stay has been associated with decreased mortality. Covid-19 patient autopsies have revealed thrombi in the microvasculature, suggesting that hypercoagulability is a prominent feature of organ failure. Negri et al. [73] performed a clinical study involving 27 Covid-19 patients admitted to Sirio-Libanes Hospital in São Paulo, Brazil. They treated these patients with heparin in therapeutic doses tailored to clinical severity. Their results suggest hypercoagulative state and microthrombosis as the main mechanisms of organ failure in Covid-19 and the potential response to early anticoagulation therapy. Following the steps of [73], several studies confirmed the importance of D-dimer as severity predictor and specific treatment based on anticoagulants as capable to reduce Covid-19 mortality and improve prognosis [16, 71, 90, 93, 99, 105, 108].

According to Klok et al. [57], Covid-19 may predispose patients to both venous and arterial thromboembolism due to excessive inflammation, hypoxia, immobilisation and diffuse intravascular coagulation. The incidence of thrombotic complications in ICU patients with COVID-19 infections is considerably high. The authors’ findings are strongly suggestive of increasing the prophylaxis towards high-prophylactic doses, even in the absence of randomized evidence. For Long et al. [68] and Panigada et al. [76], D-Dimer and Prothrombin Time are most significant indicators of severe Covid-19 cases and poor prognosis due to hypercoagulation hypercoagulability together with severe inflammatory states. Other clinical studies confirm evidences to this findings and point out to partial thromboplastin time as other important biomarker [24, 51, 61, 77, 96, 110].

According to Vargas-Vargas and Cortés-Rojo [104], ferritin is a key mediator of immune dysregulation, especially under extreme hyperferritinemia, via direct immune-suppressive and pro-inflammatory effects, contributing to the cytokine storm. Fatal outcomes by Covid-19 are accompanied by cytokine storm syndrome. Disease severity is dependent of the cytokine storm syndrome. A possible strategy to decrease ferritin levels might be the treatment with iron chelators like deferoxamine, since it is a non-toxic iron chelator clinically approved, effective for long-term iron chelation therapy in beta-thalassemia and other maladies involving iron overload. For Vargas-Vargas and Cortés-Rojo [104], manipulations decreasing dietary iron should be also considered as they have been shown to modify serum ferritin levels. Vargas-Vargas and Cortés-Rojo [104] suggest this procedure could reduce Covid-19 severity, specially in individuals with high ferritin levels morbidities like diabetes. According to Gómez-Pastora et al. [44], Rosário et al. [88], in inflammatory diseases, there is a large production of ferritin. Macrophages are responsible for most immune cells in the lung parenchyma. Macrophages also produce cytokines and may be responsible for the secretion of serum ferritin [88]. Several inflammatory stimuli, including IL-6 cytokines, can induce ferritin synthesis [88]. High concentrations of IL-6 in patients with Covid-19 are correlated with disease severity [66, 114]. Liu et al. [64] discuss the use of IL-6 blocking drugs to treat Covid-19. Liu et al. [64] discuss the pathogenesis of severe acute respiratory syndrome (SARS)-induced cytokine release syndrome (CRS). They also compare the CRS in Covid-19 with that in SARS and Middle East respiratory syndrome (MERS), and summarize the existing therapies for CRS. They propose to utilize interleukin-6 (IL-6) blockade to manage COVID-19-induced CRS and discuss several factors that should be taken into consideration for its clinical application.

For Gómez-Pastora et al. [44], as ferritin can be actively secreted at the site of infection, it is possible that ferritin can take on other functions in addition to its classic role as an iron storage protein. Ferritin has been shown to be a signaling molecule and direct mediator of the immune system [88]. Complex feedback mechanisms may exist between ferritin and cytokines in the control of pro-inflammatory and anti-inflammatory mediators, as cytokines can induce ferritin expression. Ferritin can also induce the expression of pro-and anti-inflammatory cytokines [88]. However, the pathogenic role of ferritin during inflammation remains unclear [56]. For Gómez-Pastora et al. [44], it is necessary to investigate the structure of plasma ferritin in patients with Covid-19. Ferritin is composed of 2 different subunits, H and L. Different studies have suggested that the expression of the H subunit is driven by inflammatory stimuli and ferritin H can function as an immunomodulatory molecule, exhibiting pro-inflammatory and immunosuppressive functions [56, 88]. Figure 1 illustrates the ferritin role during inflammation provoked by Covid-19 infection. Active ferritin production by macrophages and cytokines may lead to hyperferritinemia. It can induce the production of several pro-inflammatory (IL-1*β*) and anti-inflammatory cytokines (IL-10).

**Fig. 1:**
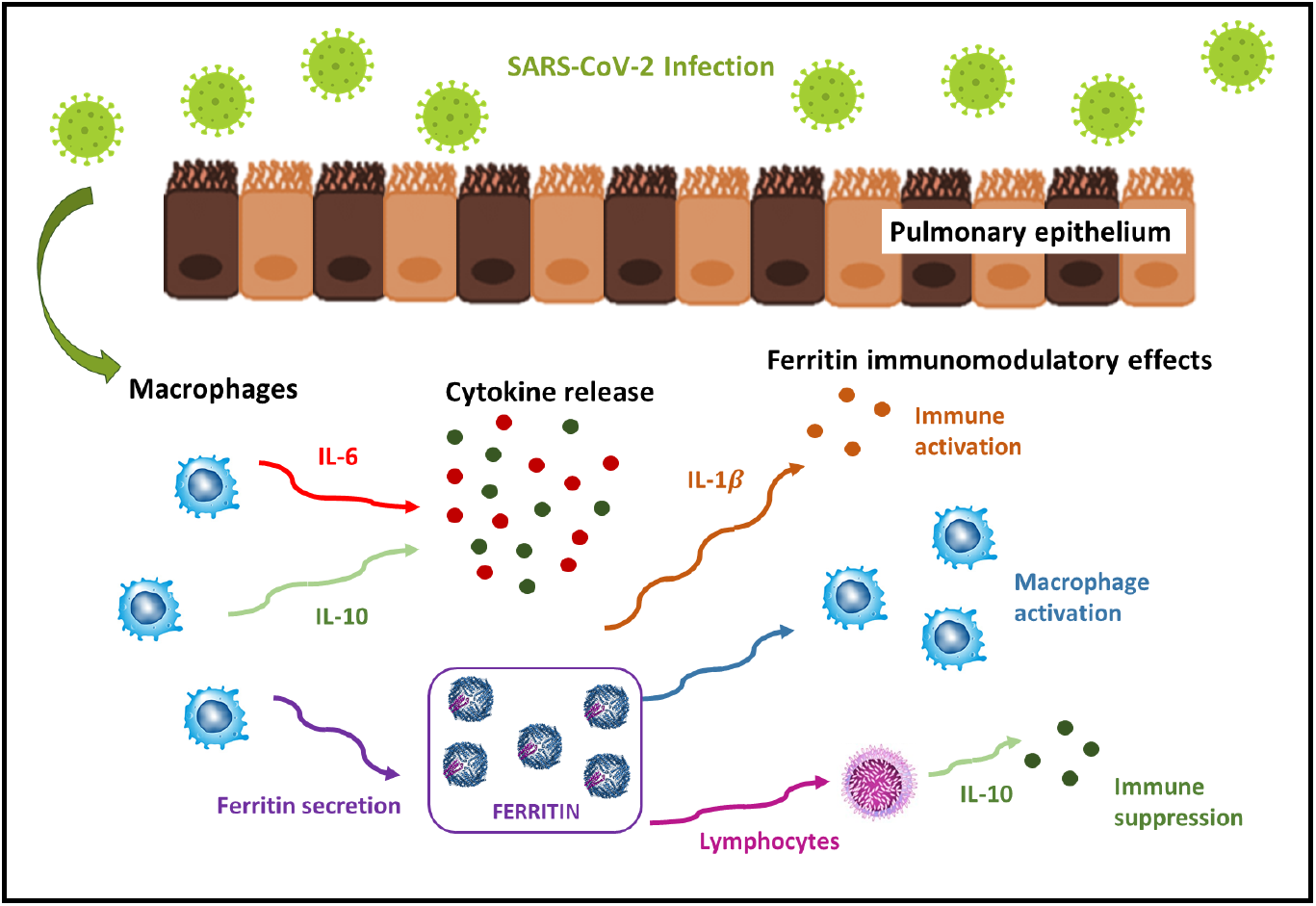
Ferritin role during inflammation provoked by Covid-19 infection. Active ferritin production by macrophages and cytokines may lead to hyperferritinemia. It can induce the production of several pro-inflammatory (IL-1*β*) and anti-inflammatory cytokines (IL-10). Illustration based on Gómez-Pastora et al. [44], Kernan and Carcillo [56], Rosário et al. [88].

For Vargas-Vargas and Cortés-Rojo [104], there is a strong evidence supporting the hypothesis that ferritin levels might be a crucial factor influencing the severity of Covid-19. Lin et al. [63] found an association between hyperferritinemia and disease severity in patients with Covid-19. Lin et al. [63] conducted a retrospective study on 147 confirmed Covid-19 patients in Changsha, China. The overall proportion of severe disease was 16.32% (24/147). The severe patients had higher levels of serum ferritin than the nonsevere patients. Multivariate logistic regression analysis indicated that the serum ferritin level on admission was an independent risk factor for disease severity in Covid-19 patients. C-reactive protein and lymphocyte counts were found to be two additional independent risk factors for disease severity through the multivariate logistic regression model. Higher serum ferritin was able to predict an increased risk of disease severity in patients with Covid-19. Serum ferritin levels positively correlate with levels of c-reactive protein, and inversely correlate with lymphocytic counts. Consequentely, the authors concluded these are critical factors considered to be associated with the disease severity in Covid-19 patients. Other studies agree on the importance of serum ferritin as a Covid-19 biomarker [7, 19, 30, 54, 101]

## 2 Materials and methods

### 2.1 Proposed method

In this work we propose two approaches of rapid protocols to support the clinical diagnosis of Covid-19 based on hematological parameters: (1) rapid diagnosis based on automatic analysis by an intelligent web system based on Particle Swarm Optimization and Random Forests; (2) rapid diagnosis by statistical inference by decision trees. In the first approach, we developed an intelligent web system for decision Covid-19 diagnostic support: Heg.IA web. This system was based on a standalone solution based on Windows and Linux PCs, Heg.IA [5]. The system started operating in the city of Paudalho, Brazil, as a prototype and has been used to support clinical diagnosis and assessment of disease severity in patients admitted to intensive and semi-intensive care units in public health units in the city since 2020.

In this approach, we used a knowledge base composed of 6215 patient records seen in health units in the city. The records were represented by age and up to 43 hematological parameters. We do not use biological sex because it is not relevant to the Covid-19 diagnostic problem. Using the Particle Swarm Optimization method, with 20 individuals evolving in 50 generations, we reduced the dimension of the attribute vectors from 43 to 8 statistically more relevant attributes. Both the original knowledge base (43 features) and the simplified knowledge base (8 features) was used to build machine learning models based on the following architectures: Multilayer Perceptron neural networks (MLP), Support Vector Machines (SVM), Naive Bayes Classifier, Bayesian Network, Decision Trees, and Random Forests. The diagram in the Figure 2 shows a summary of this solution. General method proposed: Patients with characteristic symptoms of Covid-19 are referred to a health unit and are evaluated by a medical team. This team orders blood tests. After obtaining the results, the health professional accesses Heg.IA web. On the website, he must login. Then he can enter the results of the patient’s blood tests. Upon completion, the system will generate a report with a positive or negative diagnosis for Covid-19, in addition to the hospitalization forecast. This report can be printed and used by the medical team to define the final clinical conduct.

**Fig. 2:**
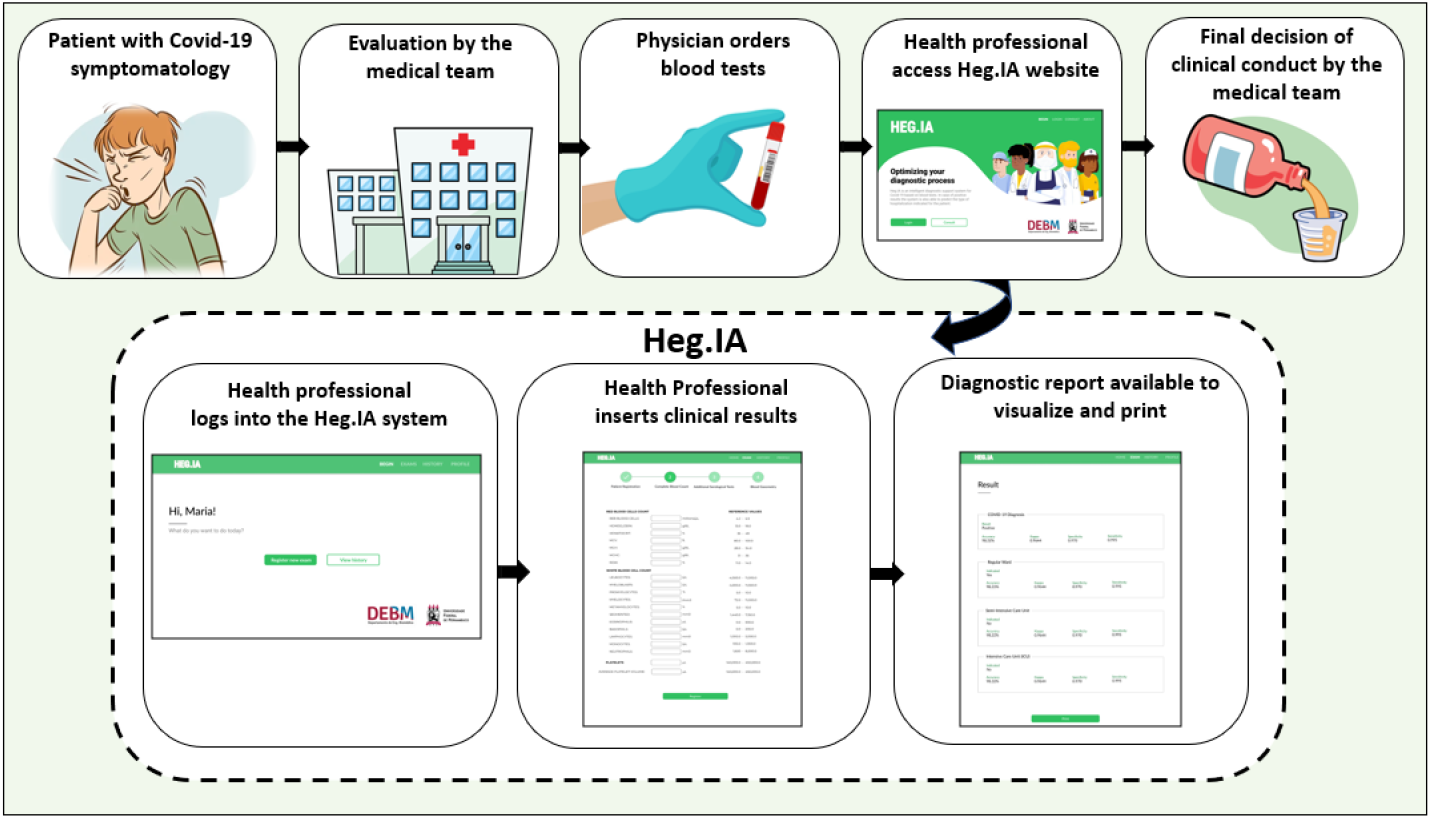
General method proposed: Patients with characteristic symptoms of Covid-19 are referred to a health unit and are evaluated by a medical team. This team orders blood tests. After obtaining the results, the health professionals access the HegIA web. On the website, they must login. Then these health professionals can enter the results of the patient’s blood tests. Upon completion, the system will generate a report with a positive or negative diagnosis for Covid-19, in addition to the hospitalization forecast. This report can be printed and used by the medical team to define the final clinical conduct.

In the second approach, we use decision trees to find the most significant relationships between hematological parameters and decision making, i.e. whether Covid-19 is positive or not. This process was used to infer that hematological parameters are statistically more relevant for clinical diagnosis without the support of an intelligent system.

All experiments were performed with 30 repetitions of 10-fold cross-validation, resulting in 300 computational experiments. As metrics of quality, we use accuracy, kappa, sensitivity, specificity, area under the ROC curve, F1-score, and precision.

### 2.2 Database

The city of Paudalho is located 38 km from the coastal capital Recife, State of Pernambuco, Brazil, has a semi-arid climate [106] and is inserted in the Atlantic Forest biome. It has a territorial area of 269,651 km^2^ and its population was around 51,357 in 2010 (last census conducted in Brazil). It is estimated that in 2020 it had 56,933 inhabitants [52]. Its demographic density of 185.06 inhabitants / km^2^ places it among the 40 most populated municipalities in the state.

Some social determinants of health provide a more functional view of the municipality in question. Directly related to health care, the municipality has 21 SUS health establishments, encompassing primary, secondary and tertiary care [52]. SUS is an acronym for “Unique Public Health System”, the Brazilian public health system. It presents only 31.5% of households with adequate sanitation (position 132 in the State of Pernambuco) [52] and infant mortality rate of 13.55 / 1,000 live births [52].

This retrospective study used the medical records of patients provided by the Health Secretariat of the city of Paudalho as a database. All procedures for this research were approved by the Research Ethics Committee at the Federal University of Pernambuco under number CAAE 34932020.3.0000.5208.

We used 6,215 records of patients seen in outpatient clinics, emergency rooms and the emergency department of SUS in the city in question, from August 30, 2019 to August 17, 2020. Of these records, 57.61% were women (3,581) and 0.27% were newborns (17) and did not have their gender identified. The mean age was 41.79 (SD = 22.94). Among men, 4.54% tested positive for Covid-19 and proportionally the most affected age group was over 90 years, where 20% of patients were victims of the disease. Among the women, 3.88% had the diagnosis of Covid-19 confirmed, with 11.86% of the patients aged 50 to 60 years and 11.88% of the age group 80 to 90 years tested positive.

The graph in Figure 3 shows the demographic stratification by sex and age of all records used in this study. We highlight the positive cases of Covid-19 from male (COVID-19 M) and female (COVID-19 F) patients using the colors blue and red, whilst the total of males and females patients are associated to orange and green, respectively. Table 1 shows the demographic details of the database, in which 2617 records are related to males, whilst 3581 are females; from these records, 119 (4.54%) are males positive for Covid-19 and 139 (3.88%) are females with Covid-19 as well, in a total of 258 Covid-19 patients distributed among moderate and severe Covid-19 cases.

**Fig. 3:**
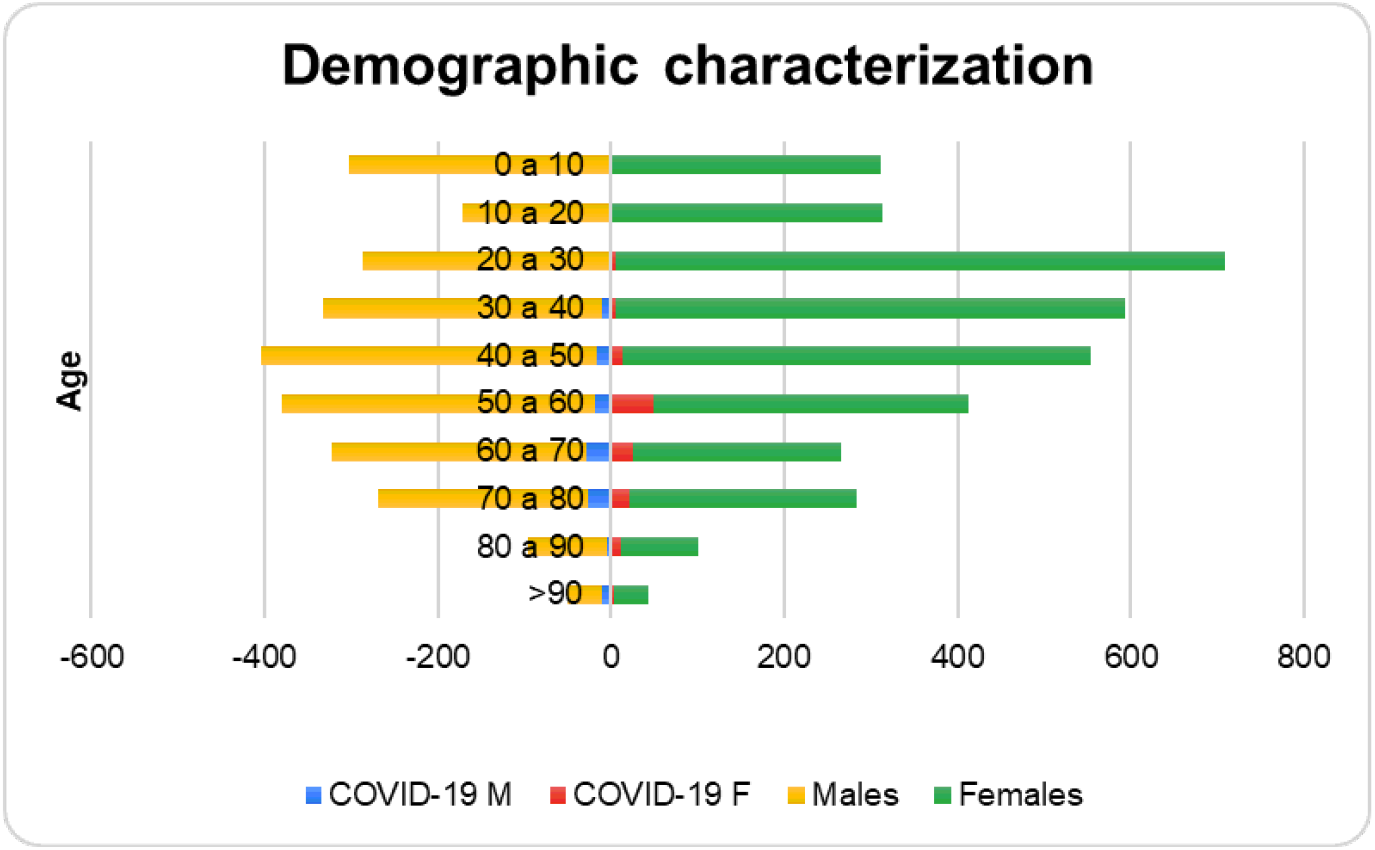
Demographic stratification by sex and age of all records used in this study. We highlight the positive cases of Covid-19 from male (COVID-19 M) and female (COVID-19 F) patients using the colors blue and red, whilst the total of males and females patients are associated to orange and green, respectively.

**Table 1:** Demographic details of the database: distribution of the number of patients by gender and by diagnosis of Covid-19, with a percentage of the population with Covid-19 by gender and by age group

| Age (years) | Males (total) | COVID-19 males | Females (total) | COVID-19 females |
| --- | --- | --- | --- | --- |
| 0 - 10 | 302 | 0 | 311 | 1 (0.32%) |
| 10 - 20 | 171 | 0 | 313 | 2 (0.63%) |
| 20 - 30 | 287 | 2 (0.69%) | 709 | 5 (0.7%) |
| 30 - 40 | 332 | 11 (3.31%) | 594 | 6 (1.01%) |
| 40 - 50 | 405 | 17 (4.19%) | 549 | 14 (2.55%) |
| 50 - 60 | 380 | 19 (5%) | 413 | 49 (11.86%) |
| 60 - 70 | 323 | 29 (8.97%) | 265 | 26 (9.81%) |
| 70 - 80 | 270 | 26 (9.62%) | 284 | 21 (7.39%) |
| 80 - 90 | 97 | 5 (5.15%) | 101 | 12 (11.88%) |
| > 90 | 50 | 10 (20%) | 42 | 3 (7.14%) |
| **Total** | **2617** | **119 (4.54%)** | **3581** | **139 (3.88%)** |

Since not all 6215 records correspond to medical records with tests that cover all the hematological parameters provided for in this work, the database has several missing values. To fill in these missing values, we used mean predictors. For each class (positive or negative for Covid-19), the sample mean for each attribute was calculated. This mean was used as a predictor for the missing values in the attribute with missing value in the vector of attributes of the corresponding class. The total of medical records with blood tests considered incomplete does not exceed 30% of the data set.

To perform this research, we initially selected 41 hematological parameters. These parameters correspond to blood exams recommended by the Ministry of Health of Brazil as an initial clinical approach and part of the Covid-19 diagnostic process [13]. Therefore, considering that health centers must already perform these tests, there is no financial loss or time spent on additional tests. On the contrary, the diagnostic process can be optimized with the system proposed here. The list of 41 hematological parameters is shown in the Figure 4. The Complete Blood Count (CBC) with differential comprises 20 of these parameters, while arterial blood gas analysis includes 9 parameters. The remaining 12 exams are those of total, indirect and direct Bilirubin; Serum Glucose; Lipase dosage; Urea; D-Dimer; Lactic Dehydrogenase; C-Reactive Protein (CRP); Creatinine; and Partial thromboplastin time (PTT) and Prothrombin time Activity from coagulogram. To this 41-item list we added patient’s age and a redundant Lymphocytes parameter. This additional Lymphocytes item corresponds to a different electronic patient record specification that used to categorize atypical lymphocytes, but was abandoned since the beginning of 2020.

**Fig. 4:**
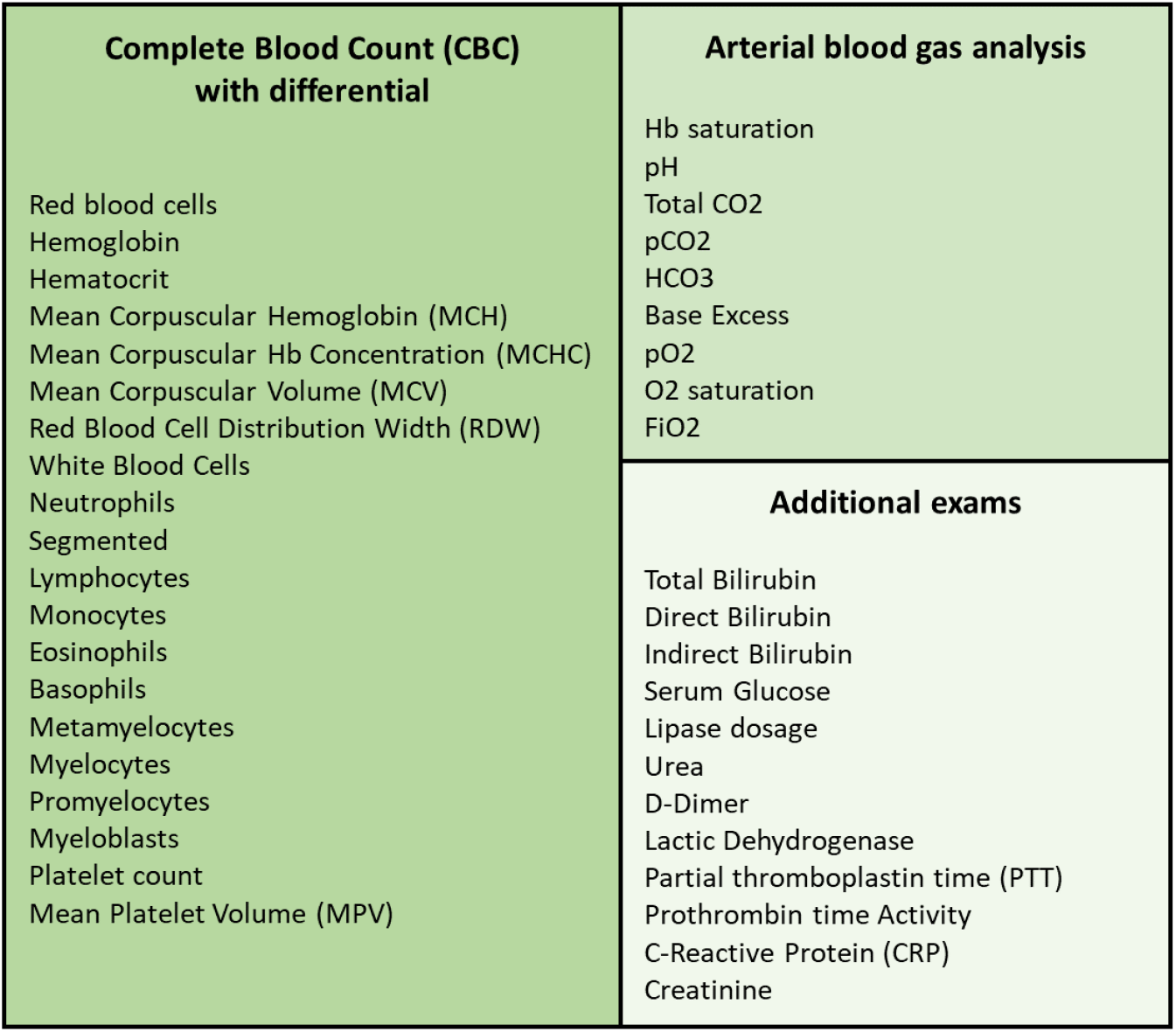
List of tests specified by the Ministry of Health of Brazil and used both in the initial assessment of symptomatic patients suspected of Covid-19 and for the control and assessment of disease progress

### 2.3 Feature Selection

The database constructed during the pre-processing was submitted to feature selection using Particle Swarm Optimization (PSO) [55, 84, 109]. As objective function, we employed a simple decision tree to guide the optimization process. We set the algorithm to 20 individuals and 500 iterations. The goal of the attribute selection is to find the most significant exams for classification tasks and also to reduce the number of required exams for diagnostic support. We chose the PSO algorithm because it is a well-established search and optimization algorithm with few parameters to be defined [55, 84, 109]. The PSO algorithm was created taking inspiration from the behavior of flocks of birds [55, 84, 109]. These flocks of birds were modeled as dynamic systems where there is a global leader, who guides the flock in a direction that optimizes a given measure of performance, and local leaders. While the global leader governs the overall dynamics of the flock of birds, local leaders are defined as those who perform best among their neighbors. Each bird is modeled using pairs of spatial position and velocity vectors. Throughout the evolution of the system, it is possible for a local leader to become a global leader, as it is also possible for leaders to become simple members of the flock of birds. Each bird represents a candidate for solving the problem of maximizing or minimizing a given objective function. In the case of maximization problems, the candidates for the solution that correspond to the global maximum and local minimums are the respective global and local leaders. Figure 5 illustrates the metaphor that inspires the PSO algorithm. PSO algorithm uses a population of randomly generated particles. In this approach, each particle corresponds to randomly generated solution and have an associated velocity and position. For each particle, each position vector is binary: 1’s and 0’s correspond to the presence or the absence of one of the 43 features in the process of training and testing the decision tree classifier associated to the objective function. The output of the objective function is the overall accuracy of a 10-fold cross validation training process. In this objective function, a decision tree is used as classifier. We employed meta-heuristic libraries developed in Java for Weka data mining platform [41]. We adopted the following feature selection methods: individual weight of 0.34, inertia weight of 0.33, mutation probability of 0.01, report frequency of 20, social weight of 0.33 [12, 55, 84, 103].

**Fig. 5:**
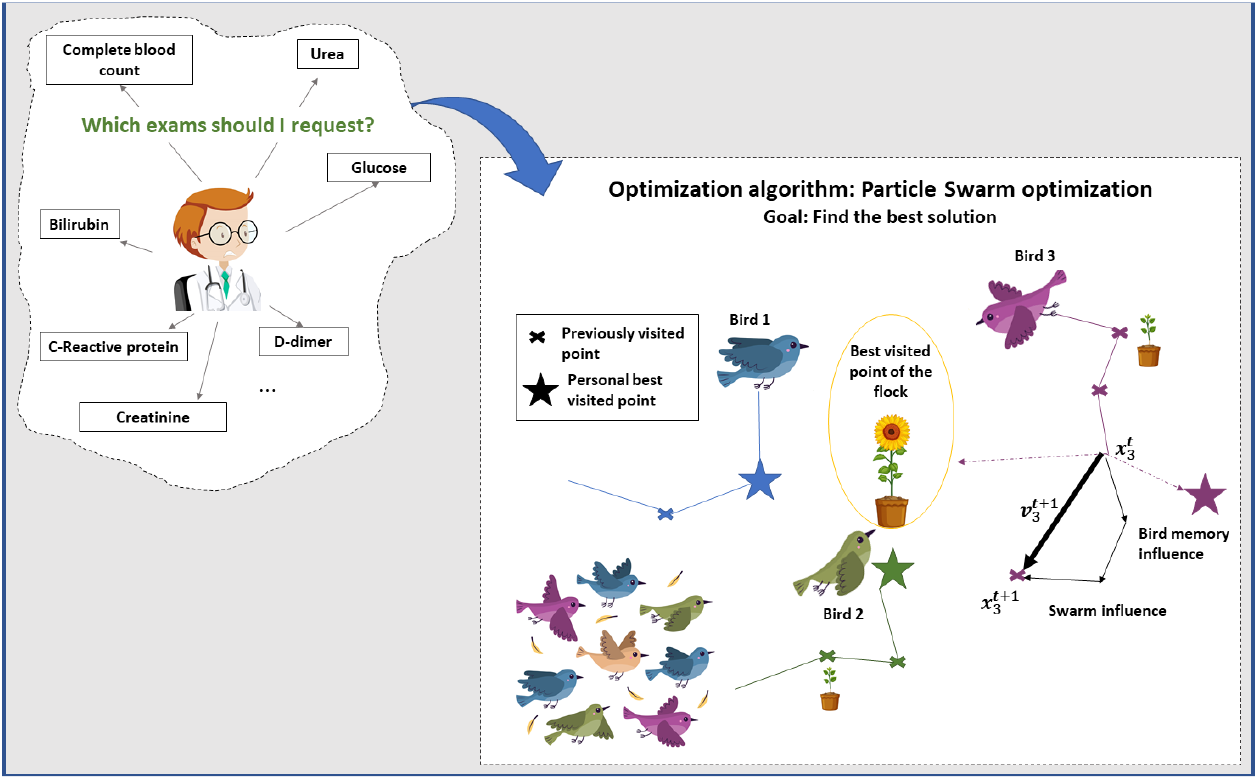
The Particle Swarm Optimization (PSO) algorithm is based on the behavior of flocks of birds. Each bird is modeled by a position vector and a velocity vector. Thus, the movement of a bird in search of resources is governed by a global leader and local leaders whose performance is defined by an objective function. In the problem of selection of the most significant exams, we use a decision tree as an objective function, with the overall accuracy of the training and testing process by 10-fold cross-validation being returned as output. Each bird is a candidate for solving the problem of maximizing accuracy. Position vectors are binary, where each coordinate corresponds to the presence or absence of one of the 43 attributes.

The feature selection implementation resulted two databases: the original database with 43 attributes, SARS-CoV-2, and the dimension-reduced database, SARS-CoV-2 (PSO). Age was not selected: it appeared as not statistically relevant according to PSO selection. The selected exams/features were the following: serum glucose, indirect bilirubin, partial thromboplastin time, lactic dehydrogenase, lipase dosage, D-dimer, ferritin, and troponin.

### 2.4 Classification

#### 2.4.1 Multilayer Perceptron

Multilayer Perceptron (MLP) consists of a generalization of the Perceptron proposed by Franklin Rosenblatt in 1958. Perceptron is the model is the simplest form of a neural network, being able to deal with linearly separable problems. Multilayer Perceptron networks, on the other hand, have several interconnected neurons (or nodes), arranged in layers: the input layer, the hidden layers and the output layer. The input layer only has the network input vector, which is passed on to the next layer. Then, each node in the next layer modifies these input values through non-linear activation functions, generating output signals. In addition, the network nodes are connected by weights, which scales these output signals. Finally, the superposition of several non-linear functions allows the mapping of the input vector to the output vector. As MLPs can have one or multiple hidden layers, this process can be repeated several times, depending on the selected architecture [6, 39, 59, 82].

Through the proper selection of activation functions and synaptic weights, an MLP is able to approximate the inputs at the desired outputs. This search and adjustment of parameters is called the training process. MLPs learn in a supervised manner. During this process, errors between the actual and desired outputs are calculated. These errors are used to adjust the network [39].

In order to adjust these weights, the backpropagation algorithm is the most computationally straightforward and common algorithm. It occurs in two phases: the forward and backward propagation. In the first step, the initial network weights are set to small random values. Then, this first input vector is propagated through the network to obtain an output. This actual output is compared with the desired one, and the error is calculated. In the second phase, the backward propagation, the error signal is propagated back through the network and the connection weights are updated, aiming to minimise the overall error. These steps can be repeated until the overall error is satisfactory [49].

MLPs and other artificial neural networks architectures are commonly used in support diagnosis applications [72], e.g. liver disease dianogis [1], heart diasese diagnosis [48], breast cancer diagnosis over breast thermography [34, 79–81, 87, 91, 92] and mammography images [25, 26, 29, 32, 32, 33, 62, 94], for recognition of intracranial epileptic seizures [86], and multiple sclerosis diagnosis support [23].

#### 2.4.2 Support Vector Machines

Support Vector machines (SVM) were created by Vladimir Vapnik and Alexey Chervonenkis [10, 27] in 1963. Their main purpose is to build a linear decision surface, called a hyperplane. The idea is that this hyperplane should be able to separate classes in the best possible way. The optimal hyperplane is found when the margin of separation between it and a given nearest point is maximum [49].

SVM classifier are known for its good generalization performance. They are employed in several healthcare applications, such as breast cancer diagnosis using thermography and mammography [25, 26, 29, 32, 32–34, 62, 80, 91, 94], diabetes mellitus diagnosis [4], heart valve diseases [22] and pulmonay infections detection [112], and also diagnosis of pulmonary cancer [97]. However, its performance varies depending on the problems complexity. The type of the machine varies with the type of kernel used to build the optimal hyperplane. Table 2 shows the kernel functions used in this study: the polynomial and RBF kernels. For the first case, it was tested exponents of value 1 (linear kernel), 2, and 3.

**Table 2:** Kernel functions of SVM

| Type of SVM | Kernel function |
| --- | --- |
| Polynomial | $K(\mathbf{x}, \mathbf{y}) = (\mathbf{x} \bullet \mathbf{y} + 1)^E$ |
| Radial Basis Function (RBF) | $K(\mathbf{x}, \mathbf{y}) = \exp(-\gamma (\mathbf{x} - \mathbf{y}) \bullet (\mathbf{x} - \mathbf{y}))$ |

#### 2.4.3 Decision Trees

Decision trees are sequential models, which combine several simple tests. They can be understood as a series of questions with “yes” and “no” answers. These tests can be the comparison of a value with a threshold or a categorical attribute compared to a set of possibilities, for instance. Thus, when analyzing the data with these tests, the decision trees will guide to a certain class in classification problems, or to a continuous value, in cases of regression problems. In this way, a decision tree is built with certain questions, called nodes. Essentially, there area four types of nodes: root, parent, child, and leaf. Starting at the root node, an instance is classified. Then, the outcome for this instance is determined ad the process continues through the tree. In addition, one node may connect to another, establishing a parent-child relationship, in which a parent node generate a child node. Finally, the terminal nodes of the tree are the leaf nodes, and they represent the final decision, that is, the predicted class or value. There are several types of decision trees, depending on the tree structure. The most popular ones are Random Tree and Random Forest. Both of them were tested in this study by using multiple parameters [58, 83].

Random Tree uses a tree built by a stochastic process. This method considers only a few randomly selected features in each node of the tree Geurts et al. [40].

In contrast, Random Forest is a model made up of many decision trees. In this case, a set of trees is built and their votes are combined to classify an instance, by means of the majority vote. Each decision tree uses a subset of attributes randomly selected from the original set of attributes [14].

#### 2.4.4 Bayesian Network and Naive Bayes classifier

Bayesian classifiers are based on Bayes’ Decision Theory. Among the most popular Bayes’ classifiers are Naive Bayes and Bayes Nets, also known as Bayesian Networks. Bayesian networks describe the probability distribution over a set of variables. They represent, in a simple way, the causal relationships of the variables of a system using Graph Theory, where the variables are the nodes and the arcs identify the relationships between the variables. In the learning process, it is necessary to calculate the probability distributions and to identify the network structure. Learning the network structure can be considered an optimization problem, where the quality measure of a network structure needs to be maximized [11, 17].

On the other hand, the Naive Bayes classifier is a simple model that considers that the domain variables are conditionally independent, that is, one characteristic is not related to the other. Its learning is done in an inductive way, presenting a set of training data and calculating the conditional probability of each attribute, given a class. Naive Bayes needs to estimate few parameters [11, 18].

#### 2.5 Parameters settings of the classifiers

All experiments were performed using the Weka Java library in 30 runs and 10-fold cross validation. The experiments were made by using the following methods:

– Naive Bayes classifier;
– Bayes Net;
– Multilayer Perceptron: one hidden layer, for 20, 50 and 100 hidden neurons;
– Support Vector Machines: we tested the following kernels, for the parameter *C* varying for 0.01, 0.1, 1, and 10: linear kernel, polynomial kernel with degree varying for 2, 3, 4, and 5; and Radial Basis Function (RBF) kernel, with *γ* of 0.01, 0.25 and 0.5;
– Decision trees: J48 and Random Tree;
– Random Forests, for 10, 20, 30, 40, 50, 60, 70, 80, 90, and 100 trees.

### 2.6 Metrics

We chose seven metrics to evaluate the performance of diagnostic tests: accuracy, precision, sensitivity, specificity, recall, precision and the area under ROC. Accuracy is the probability that the test will provide correct results, that is, be positive in sick patients and negative in healthy patients. In other words, it is the probability of the true positives and true negatives among all the results. The recall and sensitivity metrics can be calculated mathematically in the same way. They are the rate of true positives, and indicate the classifier ability to detect correctly people with Covid-19. However, they are commonly used in different contexts. In machine learning context, the term Recall is common. However, in the medical world, the use of the sensitivity metric is more frequent. Precision, on the other hand, is the fraction of the positive predictions that are actually positive. Specificity is the capacity of classifying healthy patients as negatives. It is the rate of true negatives. The Kappa index is a very good measure that can handle very well both multi-class and imbalanced class problems, as the one proposed here.

Finally, the area under the ROC curve is a measure of a classifier’s discriminating ability. That is, given two classes - a sick individual and a non-sick individual -, chosen at random, the area below the ROC curve that indicates a probability of the latter being correctly classified. If the classifier can not discriminate between these two separately, an area under a curve is equal to 0.5. When this value is the next 1, it indicates that the classifier is able to discriminate these two cases [47].

These metrics allow to discriminate between the target condition and health, in addition to quantifying the diagnostic exactitude [9]. The accuracy, precision, sensitivity, specificity, recall and precision can be calculated according to the equations in Table 3.

**Table 3:** Metrics used to evaluate classifiers performance: overall accuracy, sensitivity (recall), precision, specificity, and kappa index

| Metrics | Equation |
| --- | --- |
| Accuracy | $Accuracy = \frac{TP + TN}{TP + TN + FP + FN}$ |
| Sensitivity and Recall | $Sensitivity = Recall = \frac{TP}{TP + FN}$ |
| Precision | $Precision = \frac{TP}{TP + FP}$ |
| Specificity | $Specificity = \frac{TN}{TN + FP}$ |
| Kappa index | $k = \frac{\rho_o - \rho_e}{1 - \rho_e}$ |
| AUC | $AUC = \int TPR d(FPR)$ |

In Table 3, TP is the true positives, TN is the true negatives, FP is the false positives, and FN the false negatives, *ρ_o_* is observed agreement, or accuracy, and *ρ_e_* is the expected agreement, defined as following:

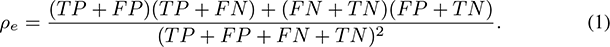

## 3 Results

The results of this research are organized in three parts: in the first part, we present the investigation of the best classification architecture for the original dataset and for the version with the reduced number of attributes through feature selection based on the PSO algorithm. The most suitable classifier was used in the implementation of the Covid-19 diagnostic support web system, Heg.IA web. In the second part, we present the use of decision trees as an alternative to build humanly intelligible models to support clinical diagnosis. These models, although less sophisticated, are important for the clinician to have a better understanding of the hematological parameters that are not only more prevalent in the diagnosis, but which can also be important for monitoring the clinical status of patients with Covid-19. Finally, in the third part, we present the prototype of the Heg.IA web system, in operation in the city of Paudalho, Brazil, since June 2020.

### 3.1 Evaluation of classifiers to support the diagnosis of Covid-19

We investigated the best classifier architectures for classifying patterns of hematological parameters. We investigate multilayer perceptron artificial neural networks, support vector machines, Bayesian networks, Naive Bayes classifiers, simple decision trees and Random Forests. We studied the behavior of these classification architectures for the 6215 patient records, 258 of which were positive for Covid-19, mostly moderate and severe cases of Covid-19. Figures 6, 7, 8, 9, 10, and 11 illustrate the accuracy values for the database with all 43 attributes. Figure 6 presents box plots for accuracy for Naive Bayes, Bayes Net and MLP classifiers. MLPs were tested for just one hidden layer, with 20, 50 and 100 hidden neurons. Figures 7, 8, 9, and 10 illustrate the accuracy behavior for SVMs with polynomial kernels (linear, 2, 3, 4, and 5 degrees) and RBF (for *γ* of 0.01, 0.25 and 0.5), considering C of 0.01, 0.10, 1.00, and 10.00, respectively. Figure 11 presents accuracy box plots for single decision trees J48 and Random Tree, and Random Forests (RF) with 10, 20, 30, 40, 50, 60, 70, 80, 90, and 100 trees. Table 4 shows the mean and standard deviation for accuracy, kappa index, sensitivity, specificity, area under the ROC curve, and F-score results, taking into account the original dataset, with all 43 attributes.

**Fig. 6:**
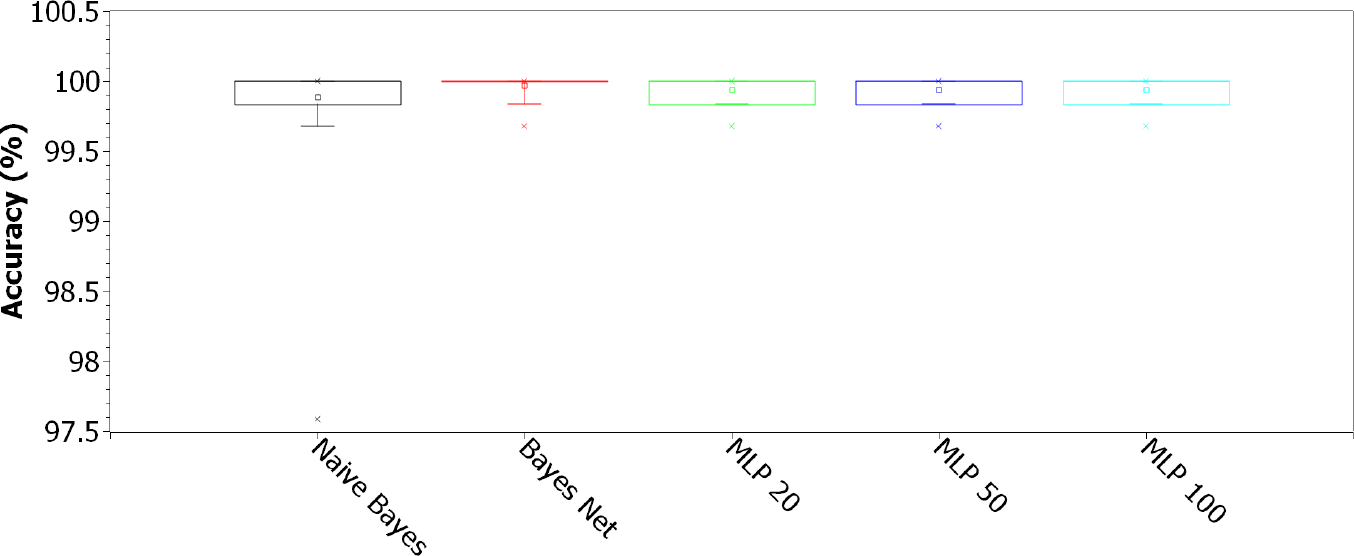
Accuracy for Naive Bayes, Bayes Net and MLP classifiers. MLPs were tested for just one hidden layer, with 20, 50 and 100 hidden neurons, considering the original 43-feature dataset.

**Fig. 7:**
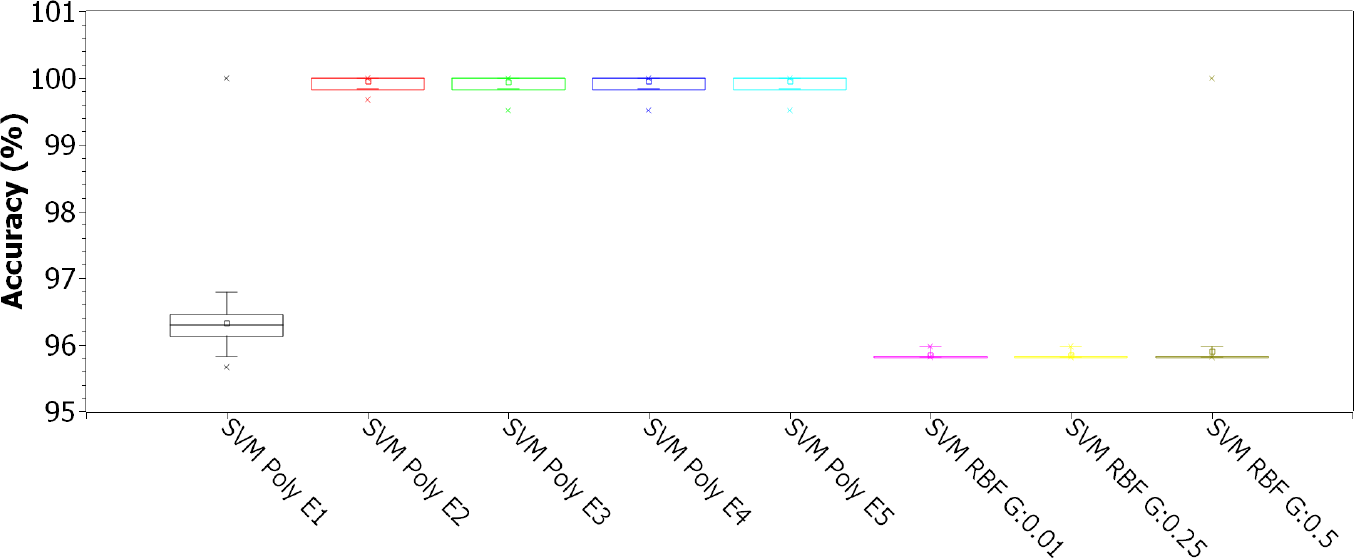
Accuracy for SVMs with polynomial kernels (linear, 2, 3, 4, and 5 degrees) and RBF (for *γ* of 0.01, 0.25 and 0.5), for C of 0.01, considering the original 43-features dataset

**Fig. 8:**
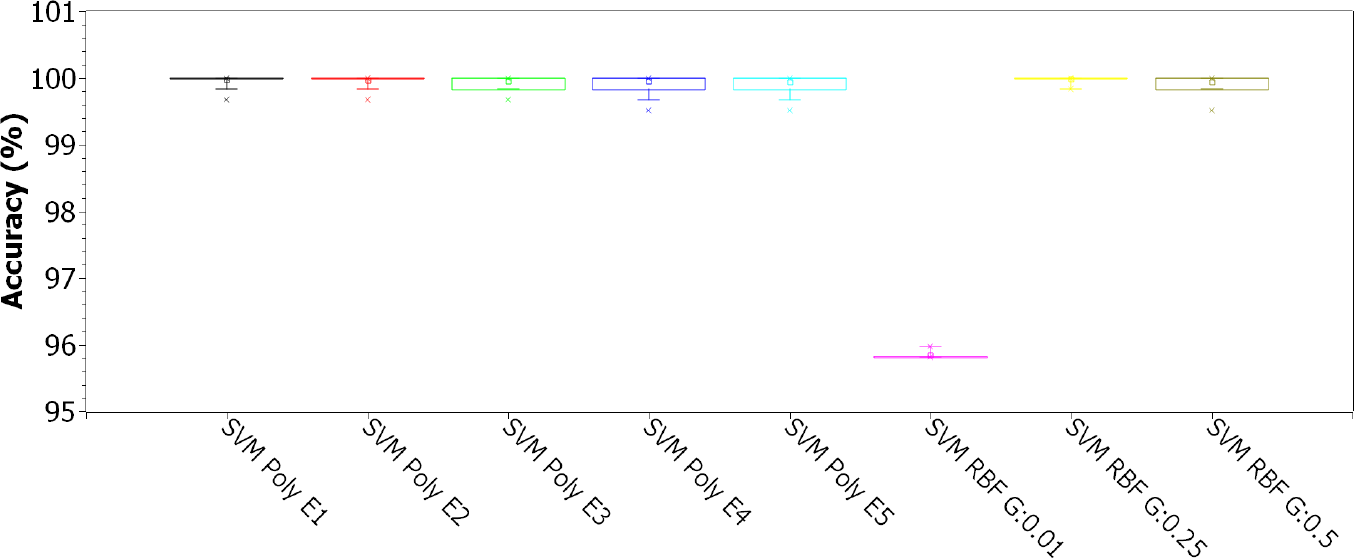
Accuracy for SVMs with polynomial kernels (linear, 2, 3, 4, and 5 degrees) and RBF (for *γ* of 0.01, 0.25 and 0.5), for C of 0.1, considering the original 43-features dataset

**Fig. 9:**
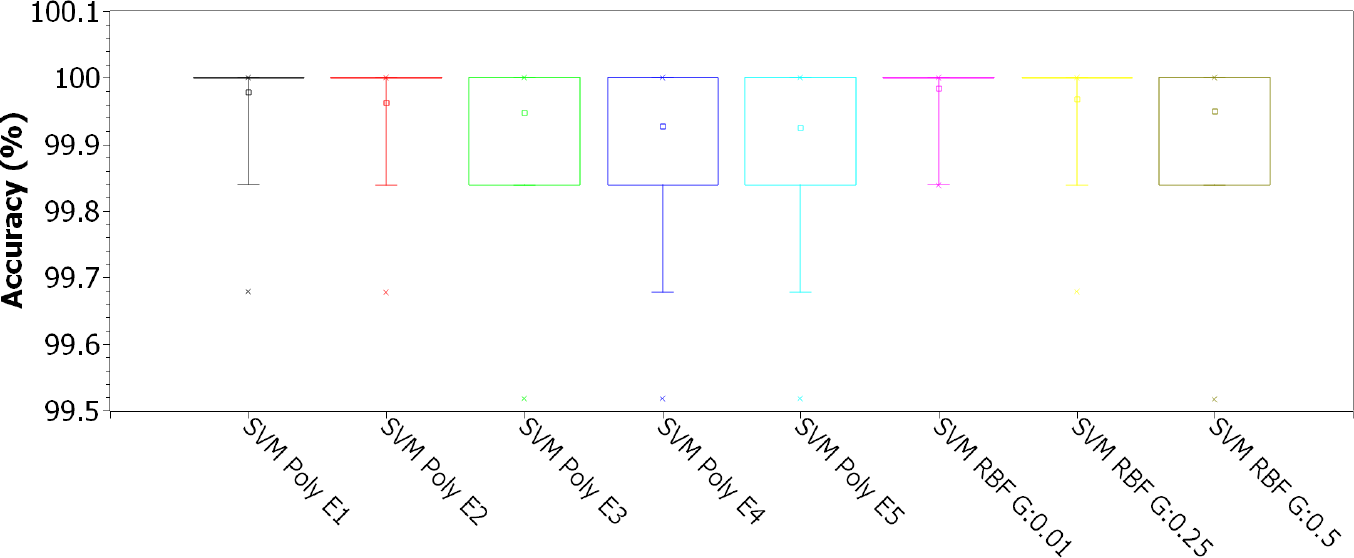
Accuracy for SVMs with polynomial kernels (linear, 2, 3, 4, and 5 degrees) and RBF (for *γ* of 0.01, 0.25 and 0.5), for C of 1.00, considering the original 43-features dataset

**Fig. 10:**
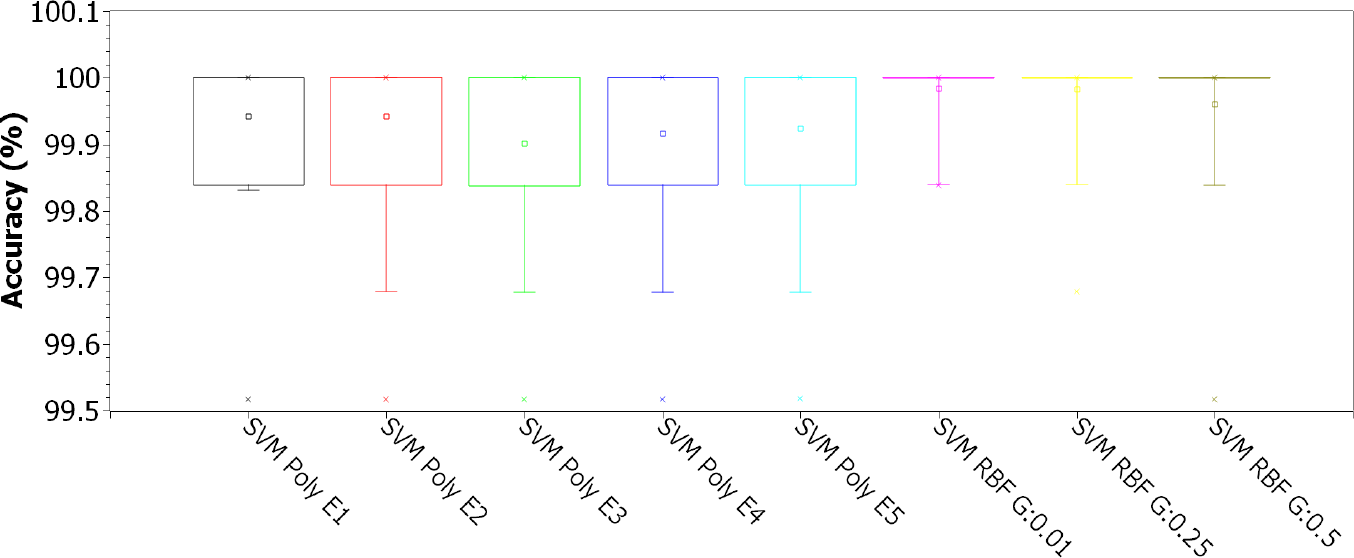
Accuracy for SVMs with polynomial kernels (linear, 2, 3, 4, and 5 degrees) and RBF (for *γ* of 0.01, 0.25 and 0.5), for C of 10.00, considering the original 43-features dataset

**Fig. 11:**
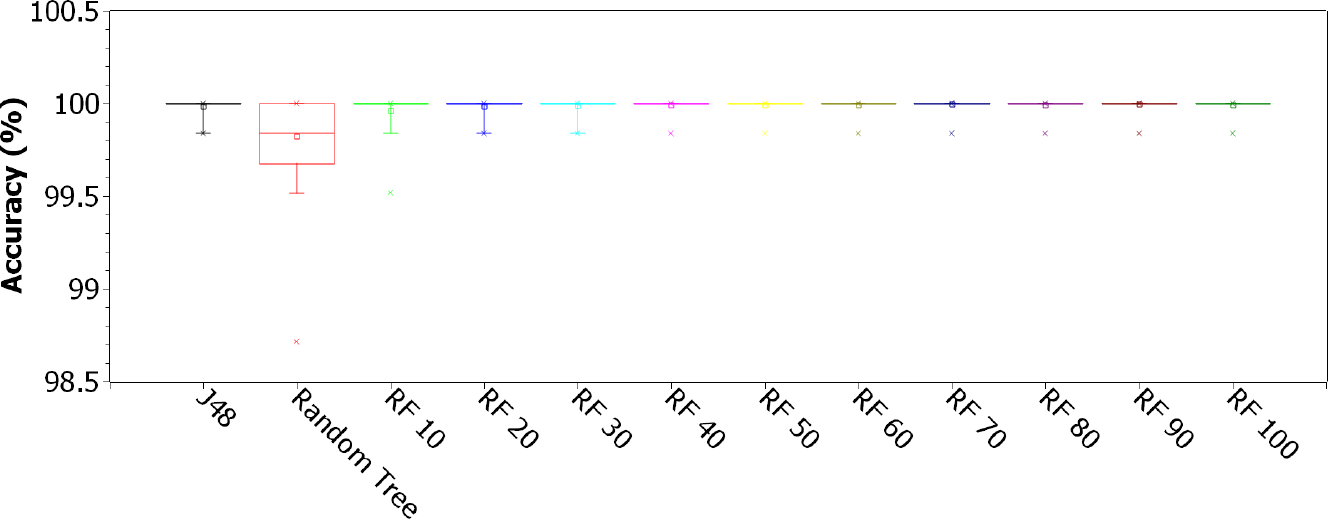
Accuracy for single decision trees J48 and Random Tree, and Random Forests (RF) with 10, 20, 30, 40, 50, 60, 70, 80, 90, and 100 trees, considering the original 43-features dataset

**Table 4:** Sample mean and standard deviation for accuracy, kappa index, sensitivity, specificity, area under ROC curve (AUC), and F1-score, for all classifiers, considering the original 43-feature dataset

| Classifier | SARS-CoV-2 |  |  |  |  |  |  |  |  |  |  |  |
| --- | --- | --- | --- | --- | --- | --- | --- | --- | --- | --- | --- | --- |
|  | Accuracy |  | Kappa |  | Sensitivity |  | Specificity |  | AUC |  | F1 Score |  |
|  | Mean | Std Dev | Mean | Std Dev | Mean | Std Dev | Mean | Std Dev | Mean | Std Dev | Mean | Std Dev |
| Naive Bayes | 99.8890 | 0.2149 | 0.9868 | 0.0230 | 0.9989 | 0.0022 | 0.9997 | 0.0031 | 0.9999 | 0.0011 | 0.9994 | 0.0011 |
| Bayes Network | 99.9716 | 0.0629 | 0.9965 | 0.0077 | 0.9997 | 0.0007 | 1.0000 | 0.0000 | 1.0000 | 0.0000 | 0.9999 | 0.0003 |
| J48 tree | 99.9839 | 0.0483 | 0.9980 | 0.0059 | 0.9998 | 0.0005 | 1.0000 | 0.0000 | 0.9999 | 0.0003 | 0.9999 | 0.0003 |
| Random Tree | 99.8220 | 0.2005 | 0.9775 | 0.0251 | 0.9992 | 0.0014 | 0.9755 | 0.0332 | 0.9873 | 0.0167 | 0.9991 | 0.0010 |
| Random Forest (10 trees) | 99.9619 | 0.0780 | 0.9951 | 0.0101 | 1.0000 | 0.0002 | 0.9913 | 0.0185 | 1.0000 | 0.0000 | 0.9998 | 0.0004 |
| Random Forest (20 trees) | 99.9850 | 0.0469 | 0.9981 | 0.0060 | 1.0000 | 0.0000 | 0.9964 | 0.0113 | 1.0000 | 0.0000 | 0.9999 | 0.0002 |
| Random Forest (30 trees) | 99.9882 | 0.0420 | 0.9985 | 0.0054 | 1.0000 | 0.0000 | 0.9972 | 0.0101 | 1.0000 | 0.0000 | 0.9999 | 0.0002 |
| Random Forest (40 trees) | 99.9925 | 0.0340 | 0.9990 | 0.0043 | 1.0000 | 0.0000 | 0.9982 | 0.0082 | 1.0000 | 0.0000 | 1.0000 | 0.0002 |
| Random Forest (50 trees) | 99.9925 | 0.0340 | 0.9990 | 0.0043 | 1.0000 | 0.0000 | 0.9982 | 0.0082 | 1.0000 | 0.0000 | 1.0000 | 0.0002 |
| Random Forest (60 trees) | 99.9941 | 0.0303 | 0.9992 | 0.0039 | 1.0000 | 0.0000 | 0.9986 | 0.0073 | 1.0000 | 0.0000 | 1.0000 | 0.0002 |
| Random Forest (70 trees) | 99.9946 | 0.0289 | 0.9993 | 0.0037 | 1.0000 | 0.0000 | 0.9987 | 0.0070 | 1.0000 | 0.0000 | 1.0000 | 0.0002 |
| Random Forest (80 trees) | 99.9941 | 0.0303 | 0.9992 | 0.0039 | 1.0000 | 0.0000 | 0.9986 | 0.0073 | 1.0000 | 0.0000 | 1.0000 | 0.0002 |
| Random Forest (90 trees) | 99.9946 | 0.0289 | 0.9993 | 0.0037 | 1.0000 | 0.0000 | 0.9987 | 0.0070 | 1.0000 | 0.0000 | 1.0000 | 0.0002 |
| Random Forest (100 trees) | 99.9930 | 0.0328 | 0.9991 | 0.0042 | 1.0000 | 0.0000 | 0.9983 | 0.0079 | 1.0000 | 0.0000 | 1.0000 | 0.0002 |
| MLP (20 neurons) | 99.9415 | 0.0870 | 0.9928 | 0.0108 | 0.9995 | 0.0008 | 0.9973 | 0.0099 | 0.9997 | 0.0007 | 0.9997 | 0.0005 |
| MLP (50 neurons) | 99.9372 | 0.0909 | 0.9922 | 0.0113 | 0.9995 | 0.0008 | 0.9963 | 0.0114 | 0.9997 | 0.0007 | 0.9997 | 0.0005 |
| MLP (100 neurons) | 99.9372 | 0.0889 | 0.9922 | 0.0110 | 0.9995 | 0.0008 | 0.9961 | 0.0116 | 0.9997 | 0.0007 | 0.9997 | 0.0005 |
| SVM polynomial E1; C = 0.01 | 96.3368 | 0.5989 | 0.1894 | 0.1581 | 0.9998 | 0.0005 | 0.1210 | 0.1454 | 0.5604 | 0.0726 | 0.9813 | 0.0031 |
| SVM polynomial E2; C = 0.01 | 99.9560 | 0.0776 | 0.9945 | 0.0096 | 0.9997 | 0.0007 | 0.9973 | 0.0099 | 0.9985 | 0.0050 | 0.9998 | 0.0004 |
| SVM polynomial E3; C = 0.01 | 99.9394 | 0.0951 | 0.9925 | 0.0118 | 0.9995 | 0.0008 | 0.9961 | 0.0116 | 0.9978 | 0.0058 | 0.9997 | 0.0005 |
| SVM polynomial E4; C = 0.01 | 99.9512 | 0.0870 | 0.9939 | 0.0108 | 0.9997 | 0.0007 | 0.9961 | 0.0116 | 0.9979 | 0.0058 | 0.9997 | 0.0005 |
| SVM polynomial E5; C = 0.01 | 99.9517 | 0.0878 | 0.9940 | 0.0109 | 0.9997 | 0.0007 | 0.9957 | 0.0129 | 0.9977 | 0.0065 | 0.9997 | 0.0005 |
| SVM RBF G = 0.01; C = 0.01 | 95.8488 | 0.0629 | 0.0000 | 0.0000 | 1.0000 | 0.0000 | 0.0000 | 0.0000 | 0.5000 | 0.0000 | 0.9788 | 0.0003 |
| SVM RBF G = 0.25; C = 0.01 | 95.8488 | 0.0629 | 0.0000 | 0.0000 | 1.0000 | 0.0000 | 0.0000 | 0.0000 | 0.5000 | 0.0000 | 0.9788 | 0.0003 |
| SVM RBF G = 0.5; C = 0.01 | 95.9013 | 0.4584 | 0.0130 | 0.1120 | 1.0000 | 0.0001 | 0.0129 | 0.1115 | 0.5065 | 0.0557 | 0.9791 | 0.0023 |
| SVM polynomial E1; C = 0.1 | 99.9705 | 0.0637 | 0.9964 | 0.0078 | 0.9997 | 0.0007 | 1.0000 | 0.0000 | 0.9998 | 0.0003 | 0.9998 | 0.0003 |
| SVM polynomial E2; C = 0.1 | 99.9657 | 0.0723 | 0.9958 | 0.0088 | 0.9996 | 0.0008 | 1.0000 | 0.0000 | 0.9998 | 0.0004 | 0.9998 | 0.0004 |
| SVM polynomial E3; cC = 0.1 | 99.9469 | 0.0875 | 0.9934 | 0.0109 | 0.9996 | 0.0007 | 0.9961 | 0.0116 | 0.9979 | 0.0058 | 0.9997 | 0.0005 |
| SVM polynomial E4; C = 0.1 | 99.9474 | 0.0940 | 0.9935 | 0.0116 | 0.9996 | 0.0008 | 0.9961 | 0.0116 | 0.9979 | 0.0059 | 0.9997 | 0.0005 |
| SVM polynomial E5; C = 0.1 | 99.9383 | 0.1065 | 0.9923 | 0.0132 | 0.9996 | 0.0009 | 0.9955 | 0.0124 | 0.9975 | 0.0063 | 0.9997 | 0.0006 |
| SVM RBF G = 0.01; C = 0.1 | 95.8488 | 0.0629 | 0.0000 | 0.0000 | 1.0000 | 0.0000 | 0.0000 | 0.0000 | 0.5000 | 0.0000 | 0.9788 | 0.0003 |
| SVM RBF G = 0.25; C = 0.1 | 99.9839 | 0.0483 | 0.9980 | 0.0059 | 0.9998 | 0.0005 | 1.0000 | 0.0000 | 0.9999 | 0.0003 | 0.9999 | 0.0003 |
| SVM RBF G = 0.5; C = 0.1 | 99.9362 | 0.0975 | 0.9919 | 0.0124 | 0.9998 | 0.0005 | 0.9885 | 0.0199 | 0.9942 | 0.0099 | 0.9997 | 0.0005 |
| SVM polynomial E1; C = 1 | 99.9796 | 0.0552 | 0.9975 | 0.0068 | 0.9998 | 0.0006 | 1.0000 | 0.0000 | 0.9999 | 0.0003 | 0.9999 | 0.0003 |
| SVM polynomial E2; C = 1 | 99.9635 | 0.0712 | 0.9955 | 0.0087 | 0.9996 | 0.0007 | 1.0000 | 0.0000 | 0.9998 | 0.0004 | 0.9998 | 0.0004 |
| SVM polynomial E3; C = 1 | 99.9480 | 0.0871 | 0.9935 | 0.0108 | 0.9996 | 0.0008 | 0.9968 | 0.0107 | 0.9982 | 0.0054 | 0.9997 | 0.0005 |
| SVM polynomial E4; C = 1 | 99.9271 | 0.1062 | 0.9909 | 0.0132 | 0.9995 | 0.0009 | 0.9932 | 0.0151 | 0.9963 | 0.0076 | 0.9996 | 0.0006 |
| SVM polynomial E5; C = 1 | 99.9244 | 0.1096 | 0.9906 | 0.0136 | 0.9995 | 0.0009 | 0.9932 | 0.0154 | 0.9963 | 0.0078 | 0.9996 | 0.0006 |
| SVM RBF G = 0.01; C = 1 | 99.9839 | 0.0483 | 0.9980 | 0.0059 | 0.9998 | 0.0005 | 1.0000 | 0.0000 | 0.9999 | 0.0003 | 0.9999 | 0.0003 |
| SVM RBF G = 0.25; C = 1 | 99.9689 | 0.0663 | 0.9961 | 0.0083 | 0.9998 | 0.0005 | 0.9964 | 0.0113 | 0.9981 | 0.0056 | 0.9998 | 0.0003 |
| SVM RBF G = 0.5; C = 1 | 99.9512 | 0.0860 | 0.9938 | 0.0109 | 0.9998 | 0.0005 | 0.9921 | 0.0165 | 0.9960 | 0.0083 | 0.9997 | 0.0004 |
| SVM polynomial E1; C = 10 | 99.9421 | 0.0945 | 0.9929 | 0.0115 | 0.9994 | 0.0010 | 1.0000 | 0.0000 | 0.9997 | 0.0005 | 0.9997 | 0.0005 |
| SVM polynomial E2; C = 10 | 99.9426 | 0.0971 | 0.9930 | 0.0119 | 0.9994 | 0.0010 | 0.9996 | 0.0038 | 0.9995 | 0.0021 | 0.9997 | 0.0005 |
| SVM polynomial E3; C = 10 | 99.9018 | 0.1145 | 0.9877 | 0.0143 | 0.9995 | 0.0009 | 0.9890 | 0.0183 | 0.9942 | 0.0092 | 0.9995 | 0.0006 |
| SVM polynomial E4; C = 10 | 99.9163 | 0.1097 | 0.9895 | 0.0137 | 0.9995 | 0.0009 | 0.9915 | 0.0164 | 0.9955 | 0.0082 | 0.9996 | 0.0006 |
| SVM polynomial E5; C = 10 | 99.9244 | 0.1096 | 0.9906 | 0.0136 | 0.9995 | 0.0009 | 0.9932 | 0.0154 | 0.9963 | 0.0078 | 0.9996 | 0.0006 |
| SVM RBF G = 0.01; C = 10 | 99.9839 | 0.0483 | 0.9980 | 0.0059 | 0.9998 | 0.0005 | 1.0000 | 0.0000 | 0.9999 | 0.0003 | 0.9999 | 0.0003 |
| SVM RBF G = 0.25; C = 10 | 99.9828 | 0.0531 | 0.9979 | 0.0065 | 0.9998 | 0.0006 | 1.0000 | 0.0000 | 0.9999 | 0.0003 | 0.9999 | 0.0003 |
| SVM RBF G = 0.5; C = 10 | 99.9603 | 0.0799 | 0.9950 | 0.0101 | 0.9998 | 0.0005 | 0.9943 | 0.0144 | 0.9971 | 0.0072 | 0.9998 | 0.0004 |

Figures 12, 13, 14, 15, 16, and 17 illustrate the accuracy values for the 8-feature database obtained after feature selecion with the PSO algorithm. Figure 12 presents box plots for accuracy for Naive Bayes, Bayes Net and MLP classifiers. MLPs were tested for just one hidden layer, with 20, 50 and 100 hidden neurons. Figures 13, 14, 15, and 16 illustrate the accuracy behavior for SVMs with polynomial kernels (linear, 2, 3, 4, and 5 degrees) and RBF (for *γ* of 0.01, 0.25 and 0.5), considering C of 0.01, 0.10, 1.00, and 10.00, respectively. Figure 17 presents accuracy box plots for single decision trees J48 and Random Tree, and Random Forests (RF) with 10, 20, 30, 40, 50, 60, 70, 80, 90, and 100 trees. Table 5 shows the mean and standard deviation for accuracy, kappa index, sensitivity, specificity, area under the ROC curve, and F-score results, taking into account the 8-feature PSO dimension-reduced dataset.

**Fig. 12:**
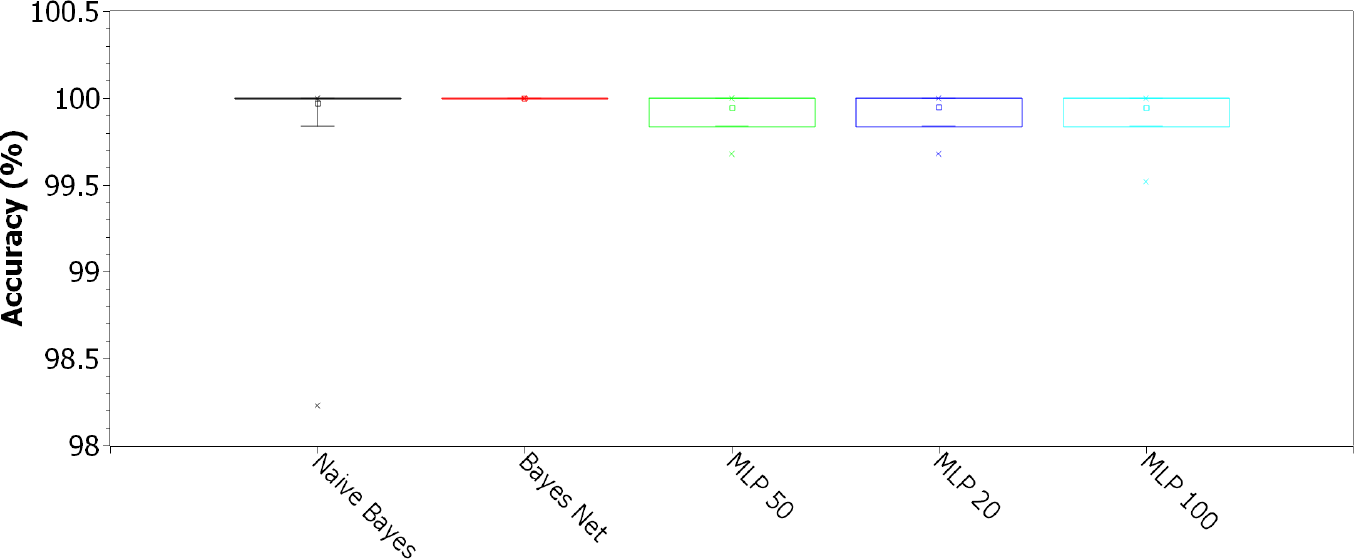
Accuracy for Naive Bayes, Bayes Net and MLP classifiers. MLPs were tested for just one hidden layer, with 20, 50 and 100 hidden neurons, considering the 8-feature PSO dimension-reduced dataset.

**Fig. 13:**
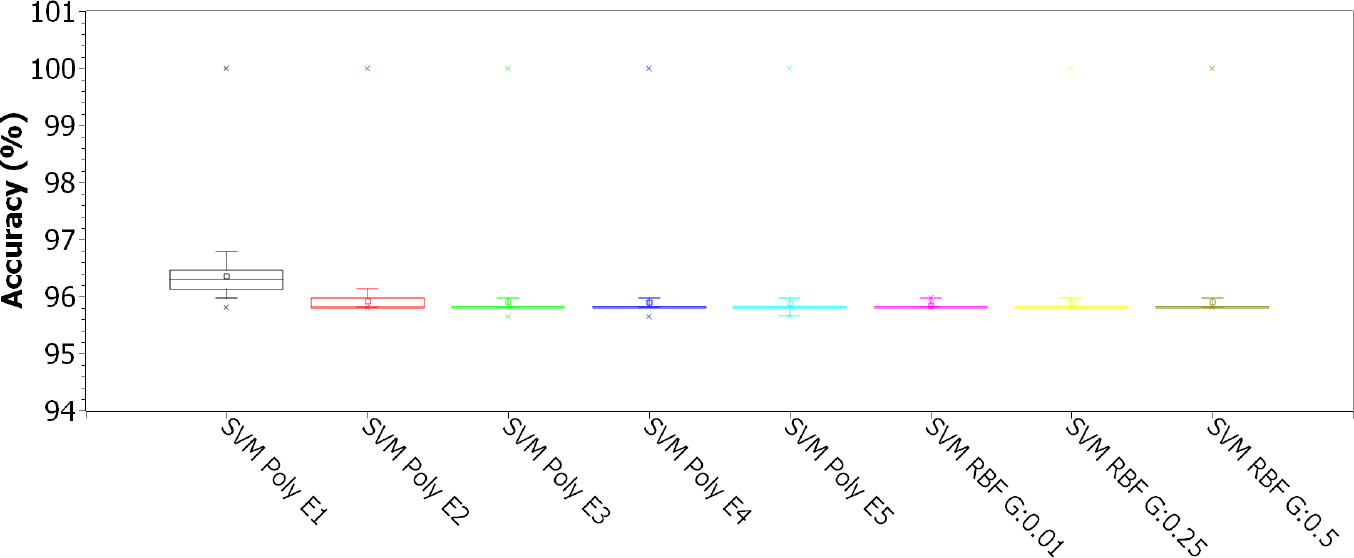
Accuracy for SVMs with polynomial kernels (linear, 2, 3, 4, and 5 degrees) and RBF (for *γ* of 0.01, 0.25 and 0.5), for C of 0.01, considering the 8-feature PSO dimension-reduced dataset

**Fig. 14:**
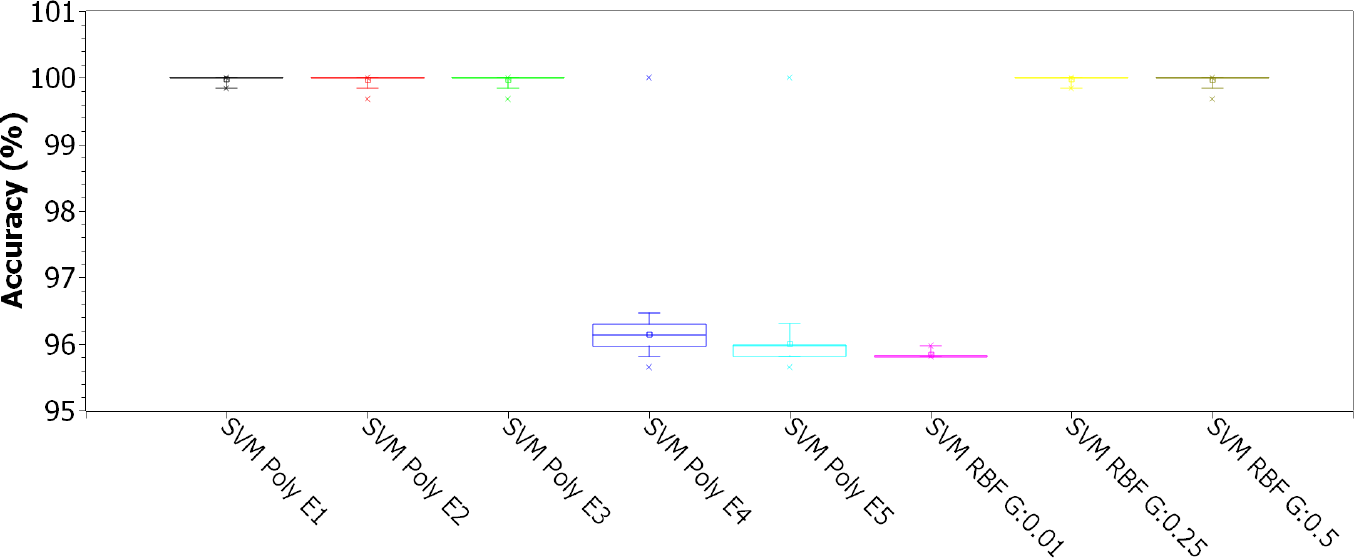
Accuracy for SVMs with polynomial kernels (linear, 2, 3, 4, and 5 degrees) and RBF (for *γ* of 0.01, 0.25 and 0.5), for C of 0.1, considering the 8-feature PSO dimension-reduced dataset

**Fig. 15:**
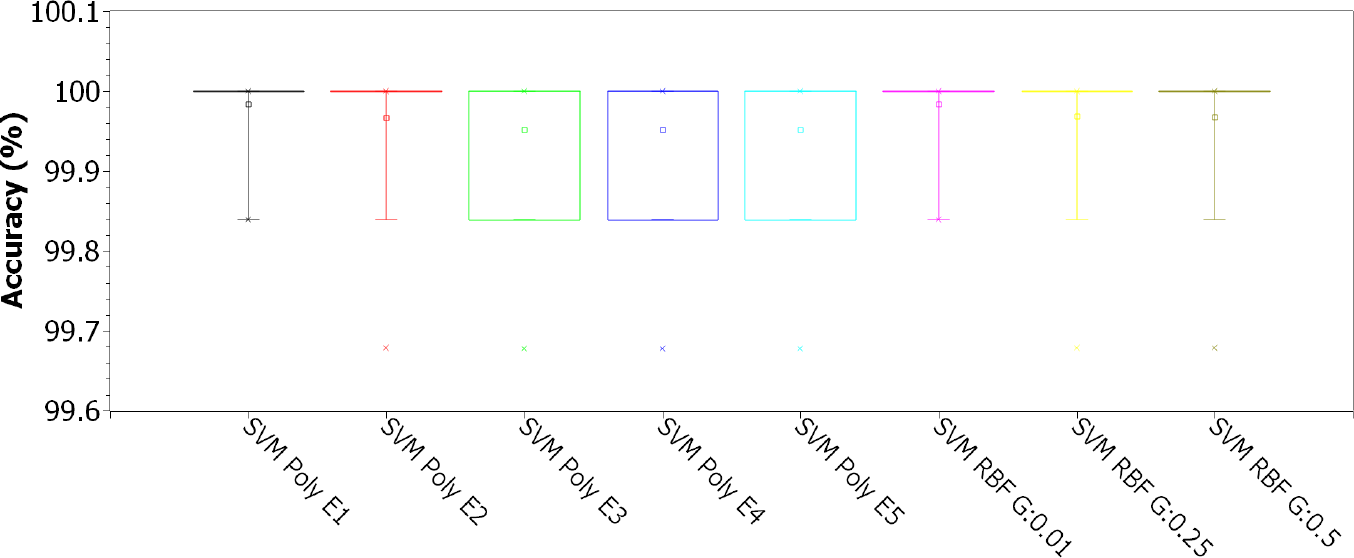
Accuracy for SVMs with polynomial kernels (linear, 2, 3, 4, and 5 degrees) and RBF (for *γ* of 0.01, 0.25 and 0.5), for C of 1.00, considering the 8-feature PSO dimension-reduced dataset

**Fig. 16:**
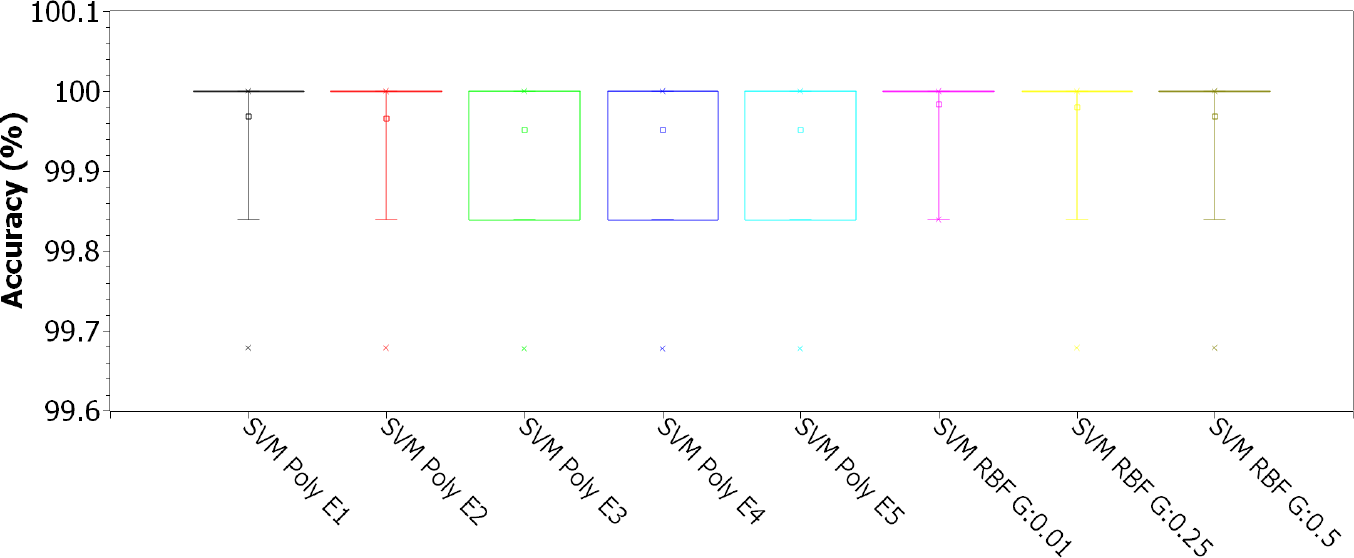
Accuracy for SVMs with polynomial kernels (linear, 2, 3, 4, and 5 degrees) and RBF (for *γ* of 0.01, 0.25 and 0.5), for C of 10.00, considering the 8-feature PSO dimension-reduced dataset

**Fig. 17:**
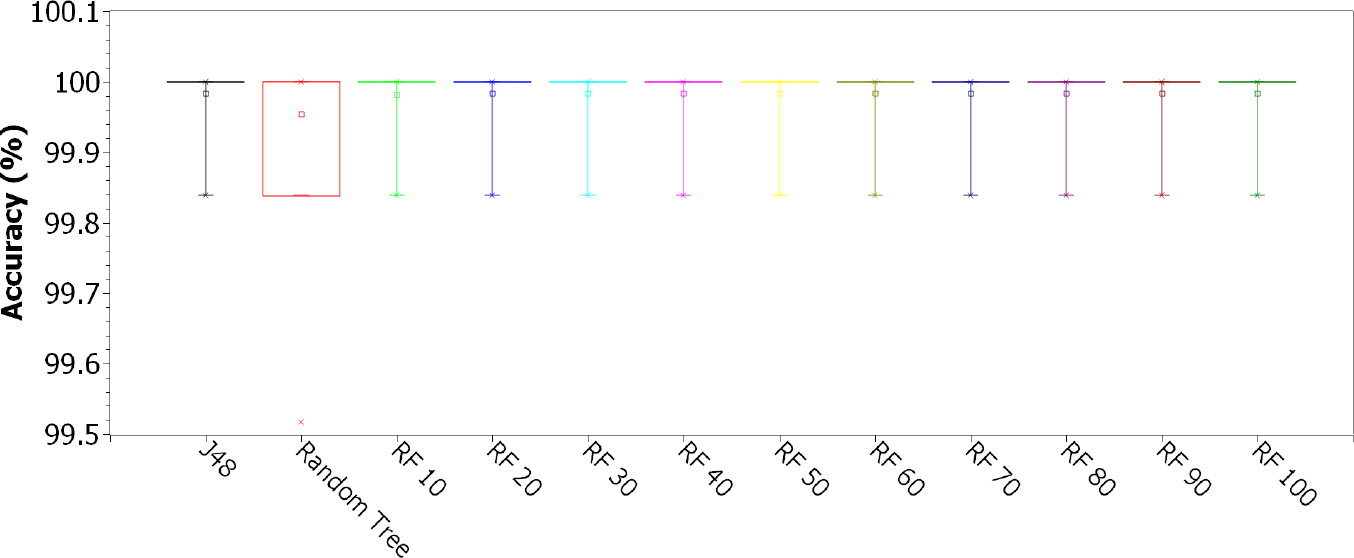
Accuracy for single decision trees J48 and Random Tree, and Random Forests (RF) with 10, 20, 30, 40, 50, 60, 70, 80, 90, and 100 trees, considering the 8-feature PSO dimension-reduced dataset

**Table 5:** Sample mean and standard deviation for accuracy, kappa index, sensitivity, specificity, area under ROC curve (AUC), and F1-score, for all classifiers, considering the 8-feature PSO dimension-reduced dataset

| Classifier | SARS-CoV-2 PSO |  |  |  |  |  |  |  |  |  |  |  |
| --- | --- | --- | --- | --- | --- | --- | --- | --- | --- | --- | --- | --- |
|  | Accuracy |  | Kappa |  | Sensitivity |  | Specificity |  | AUC |  | F1 Score |  |
|  | Mean | Std Dev | Mean | Std Dev | Mean | Std Dev | Mean | Std Dev | Mean | Std Dev | Mean | Std Dev |
| Naive Bayes | 99.9689 | 0.1369 | 0.9963 | 0.0149 | 0.9997 | 0.0014 | 1.0000 | 0.0000 | 1.0000 | 0.0001 | 0.9998 | 0.0007 |
| Bayes Network | 100.0000 | 0.0000 | 1.0000 | 0.0000 | 1.0000 | 0.0000 | 1.0000 | 0.0000 | 1.0000 | 0.0000 | 1.0000 | 0.0000 |
| J48 tree | 99.9839 | 0.0483 | 0.9980 | 0.0059 | 0.9998 | 0.0005 | 1.0000 | 0.0000 | 0.9999 | 0.0003 | 0.9999 | 0.0003 |
| Random Tree | 99.9539 | 0.0849 | 0.9942 | 0.0107 | 0.9998 | 0.0006 | 0.9939 | 0.0151 | 0.9969 | 0.0076 | 0.9998 | 0.0004 |
| Random Forest (10 trees) | 99.9812 | 0.0517 | 0.9977 | 0.0064 | 0.9998 | 0.0005 | 0.9992 | 0.0054 | 1.0000 | 0.0001 | 0.9999 | 0.0003 |
| Random Forest (20 trees) | 99.9839 | 0.0483 | 0.9980 | 0.0059 | 0.9998 | 0.0005 | 1.0000 | 0.0000 | 1.0000 | 0.0000 | 0.9999 | 0.0003 |
| Random Forest (30 trees) | 99.9839 | 0.0483 | 0.9980 | 0.0059 | 0.9998 | 0.0005 | 1.0000 | 0.0000 | 1.0000 | 0.0000 | 0.9999 | 0.0003 |
| Random Forest (40 trees) | 99.9839 | 0.0483 | 0.9980 | 0.0059 | 0.9998 | 0.0005 | 1.0000 | 0.0000 | 1.0000 | 0.0000 | 0.9999 | 0.0003 |
| Random Forest (50 trees) | 99.9839 | 0.0483 | 0.9980 | 0.0059 | 0.9998 | 0.0005 | 1.0000 | 0.0000 | 1.0000 | 0.0000 | 0.9999 | 0.0003 |
| Random Forest (60 trees) | 99.9839 | 0.0483 | 0.9980 | 0.0059 | 0.9998 | 0.0005 | 1.0000 | 0.0000 | 1.0000 | 0.0000 | 0.9999 | 0.0003 |
| Random Forest (70 trees) | 99.9839 | 0.0483 | 0.9980 | 0.0059 | 0.9998 | 0.0005 | 1.0000 | 0.0000 | 1.0000 | 0.0000 | 0.9999 | 0.0003 |
| Random Forest (80 trees) | 99.9839 | 0.0483 | 0.9980 | 0.0059 | 0.9998 | 0.0005 | 1.0000 | 0.0000 | 1.0000 | 0.0000 | 0.9999 | 0.0003 |
| Random Forest (90 trees) | 99.9839 | 0.0483 | 0.9980 | 0.0059 | 0.9998 | 0.0005 | 1.0000 | 0.0000 | 1.0000 | 0.0000 | 0.9999 | 0.0003 |
| Random Forest (100 trees) | 99.9839 | 0.0483 | 0.9980 | 0.0059 | 0.9998 | 0.0005 | 1.0000 | 0.0000 | 1.0000 | 0.0000 | 0.9999 | 0.0003 |
| MLP (20 neurons) | 99.9496 | 0.0845 | 0.9937 | 0.0106 | 0.9997 | 0.0007 | 0.9955 | 0.0132 | 0.9999 | 0.0005 | 0.9997 | 0.0004 |
| MLP (50 neurons) | 99.9458 | 0.0888 | 0.9932 | 0.0111 | 0.9997 | 0.0007 | 0.9946 | 0.0145 | 0.9999 | 0.0005 | 0.9997 | 0.0005 |
| MLP (100 neurons) | 99.9458 | 0.0888 | 0.9932 | 0.0112 | 0.9997 | 0.0007 | 0.9943 | 0.0154 | 0.9999 | 0.0005 | 0.9997 | 0.0005 |
| SVM polynomial E1; C = 0.01 | 96.3626 | 0.5919 | 0.1952 | 0.1561 | 1.0000 | 0.0001 | 0.1240 | 0.1440 | 0.5620 | 0.0720 | 0.9814 | 0.0030 |
| SVM polynomial E2; C = 0.01 | 95.9281 | 0.4686 | 0.0245 | 0.1154 | 1.0000 | 0.0002 | 0.0195 | 0.1152 | 0.5098 | 0.0575 | 0.9792 | 0.0024 |
| SVM polynomial E3; C = 0.01 | 95.9078 | 0.4678 | 0.0160 | 0.1137 | 1.0000 | 0.0002 | 0.0149 | 0.1150 | 0.5074 | 0.0574 | 0.9791 | 0.0024 |
| SVM polynomial E4; C = 0.01 | 95.9024 | 0.4675 | 0.0136 | 0.1133 | 1.0000 | 0.0002 | 0.0136 | 0.1149 | 0.5068 | 0.0574 | 0.9791 | 0.0024 |
| SVM polynomial E5; C = 0.01 | 95.8890 | 0.4708 | 0.0131 | 0.1133 | 0.9998 | 0.0005 | 0.0135 | 0.1149 | 0.5066 | 0.0574 | 0.9790 | 0.0024 |
| SVM RBF G=0.01; C = 0.01 | 95.8488 | 0.0629 | 0.0000 | 0.0000 | 1.0000 | 0.0000 | 0.0000 | 0.0000 | 0.5000 | 0.0000 | 0.9788 | 0.0003 |
| SVM RBF G = 0.25; C = 0.01 | 95.9019 | 0.4631 | 0.0130 | 0.1125 | 1.0000 | 0.0000 | 0.0128 | 0.1105 | 0.5064 | 0.0552 | 0.9791 | 0.0024 |
| SVM RBF G=0.5; C = 0.01 | 95.9067 | 0.4728 | 0.0148 | 0.1146 | 1.0000 | 0.0001 | 0.0142 | 0.1151 | 0.5071 | 0.0575 | 0.9791 | 0.0024 |
| SVM polynomial E1; C = 0.1 | 99.9839 | 0.0483 | 0.9980 | 0.0059 | 0.9998 | 0.0005 | 1.0000 | 0.0000 | 0.9999 | 0.0003 | 0.9999 | 0.0003 |
| SVM polynomial E2; C = 0.1 | 99.9673 | 0.0687 | 0.9960 | 0.0084 | 0.9997 | 0.0007 | 1.0000 | 0.0000 | 0.9998 | 0.0004 | 0.9998 | 0.0004 |
| SVM polynomial E3; C = 0.1 | 99.9673 | 0.0687 | 0.9960 | 0.0084 | 0.9997 | 0.0007 | 1.0000 | 0.0000 | 0.9998 | 0.0004 | 0.9998 | 0.0004 |
| SVM polynomial E4; C = 0.1 | 96.1480 | 0.4815 | 0.1227 | 0.1305 | 0.9998 | 0.0005 | 0.0763 | 0.1190 | 0.5381 | 0.0594 | 0.9803 | 0.0025 |
| SVM polynomial E5; C = 0.1 | 96.0097 | 0.4765 | 0.0659 | 0.1237 | 0.9998 | 0.0005 | 0.0430 | 0.1171 | 0.5214 | 0.0585 | 0.9796 | 0.0024 |
| SVM RBF G=0.01; C = 0.1 | 95.8488 | 0.0629 | 0.0000 | 0.0000 | 1.0000 | 0.0000 | 0.0000 | 0.0000 | 0.5000 | 0.0000 | 0.9788 | 0.0003 |
| SVM RBF G=0.25; C = 0.1 | 99.9839 | 0.0483 | 0.9980 | 0.0059 | 0.9998 | 0.0005 | 1.0000 | 0.0000 | 0.9999 | 0.0003 | 0.9999 | 0.0003 |
| SVM RBF G=0.5; C = 0.1 | 99.9689 | 0.0663 | 0.9961 | 0.0083 | 0.9998 | 0.0005 | 0.9964 | 0.0113 | 0.9981 | 0.0057 | 0.9998 | 0.0003 |
| SVM polynomial E1; C = 1 | 99.9839 | 0.0483 | 0.9980 | 0.0059 | 0.9998 | 0.0005 | 1.0000 | 0.0000 | 0.9999 | 0.0003 | 0.9999 | 0.0003 |
| SVM polynomial E2; C = 1 | 99.9673 | 0.0687 | 0.9960 | 0.0084 | 0.9997 | 0.0007 | 1.0000 | 0.0000 | 0.9998 | 0.0004 | 0.9998 | 0.0004 |
| SVM polynomial E3; C = 1 | 99.9517 | 0.0816 | 0.9941 | 0.0100 | 0.9995 | 0.0009 | 1.0000 | 0.0000 | 0.9997 | 0.0004 | 0.9997 | 0.0004 |
| SVM polynomial E4; C = 1 | 99.9517 | 0.0816 | 0.9941 | 0.0100 | 0.9995 | 0.0009 | 1.0000 | 0.0000 | 0.9997 | 0.0004 | 0.9997 | 0.0004 |
| SVM polynomial E5; C = 1 | 99.9517 | 0.0816 | 0.9941 | 0.0100 | 0.9995 | 0.0009 | 1.0000 | 0.0000 | 0.9997 | 0.0004 | 0.9997 | 0.0004 |
| SVM RBF G=0.01; C = 1 | 99.9839 | 0.0483 | 0.9980 | 0.0059 | 0.9998 | 0.0005 | 1.0000 | 0.0000 | 0.9999 | 0.0003 | 0.9999 | 0.0003 |
| SVM RBF G=0.25; C = 1 | 99.9684 | 0.0667 | 0.9960 | 0.0084 | 0.9998 | 0.0005 | 0.9963 | 0.0115 | 0.9980 | 0.0057 | 0.9998 | 0.0003 |
| SVM RBF G=0.5; C = 1 | 99.9678 | 0.0671 | 0.9960 | 0.0084 | 0.9998 | 0.0005 | 0.9961 | 0.0117 | 0.9980 | 0.0058 | 0.9998 | 0.0003 |
| SVM polynomial E1; C = 10 | 99.9689 | 0.0676 | 0.9962 | 0.0083 | 0.9997 | 0.0007 | 1.0000 | 0.0000 | 0.9998 | 0.0004 | 0.9998 | 0.0004 |
| SVM polynomial E2; C = 10 | 99.9657 | 0.0711 | 0.9958 | 0.0087 | 0.9996 | 0.0007 | 1.0000 | 0.0000 | 0.9998 | 0.0004 | 0.9998 | 0.0004 |
| SVM polynomial E3; C = 10 | 99.9517 | 0.0816 | 0.9941 | 0.0100 | 0.9995 | 0.0009 | 1.0000 | 0.0000 | 0.9997 | 0.0004 | 0.9997 | 0.0004 |
| SVM polynomial E4; C = 10 | 99.9517 | 0.0816 | 0.9941 | 0.0100 | 0.9995 | 0.0009 | 1.0000 | 0.0000 | 0.9997 | 0.0004 | 0.9997 | 0.0004 |
| SVM polynomial E5; C = 10 | 99.9517 | 0.0816 | 0.9941 | 0.0100 | 0.9995 | 0.0009 | 1.0000 | 0.0000 | 0.9997 | 0.0004 | 0.9997 | 0.0004 |
| SVM RBF G=0.01; C = 10 | 99.9839 | 0.0483 | 0.9980 | 0.0059 | 0.9998 | 0.0005 | 1.0000 | 0.0000 | 0.9999 | 0.0003 | 0.9999 | 0.0003 |
| SVM RBF G=0.25; C = 10 | 99.9807 | 0.0540 | 0.9976 | 0.0067 | 0.9998 | 0.0005 | 0.9992 | 0.0054 | 0.9995 | 0.0027 | 0.9999 | 0.0003 |
| SVM RBF G=0.5; C = 10 | 99.9684 | 0.0667 | 0.9960 | 0.0084 | 0.9998 | 0.0005 | 0.9963 | 0.0115 | 0.9980 | 0.0057 | 0.9998 | 0.0003 |

### 3.2 Humanly intelligible models to support the clinical diagnosis of Covid-19

Figure 18 shows the J48 decision tree trained on the 8-feature dataset: serum glucose, partial thromboplastin time, troponin, lipase dosage, lactic dehydrogenase, ferritin, indirect bilirubin, and D-dimer. Training was done using 10-fold cross validation. The result shows that, for all 6215 records, the accuracy was 99.9839%, the sensitivity, specificity, AUC, and F-score were maximum. Only a single instance was misclassified: a negative record for Covid-19 out of the total of 5457 negatives was classified as positive. All Covid-19 positive records were classified correctly.

**Fig. 18:**
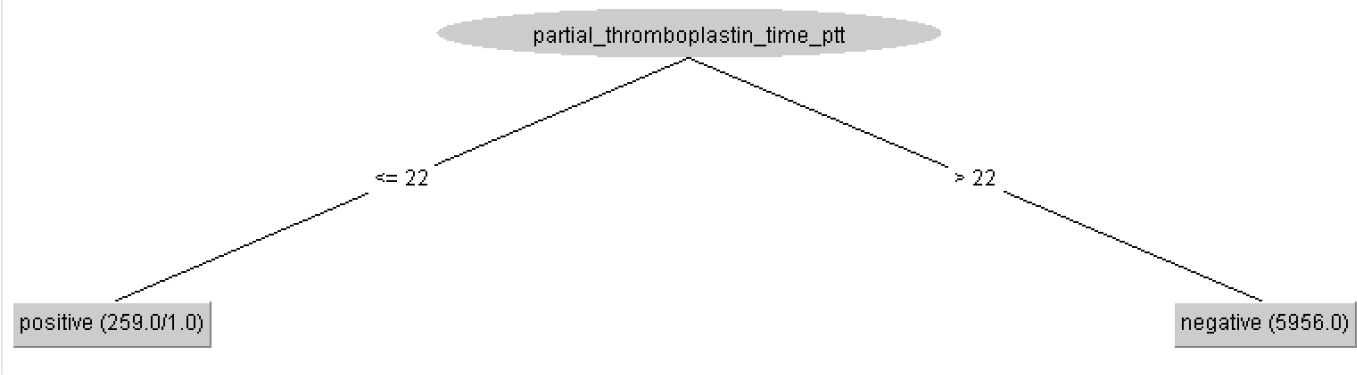
Clinical decision protocol based on J48 decision tree trained on the 8-feature dataset: serum glucose, partial thromboplastin time, troponin, lipase dosage, lactic dehydrogenase, ferritin, indirect bilirubin, and D-dimer. Training was done using 10-fold cross validation. Reduced partial thromboplastin time (< 22) is associated to symptomatic Covid-19 case. The accuracy was 99.9839%, the sensitivity, specificity, AUC, and F-score were maximum. We had 1 false positive.

To assess the influence of the other 7 hematological parameters on the diagnosis of Covid-19, we retrained the J48 decision tree after removing the partial thromboplastin time. Training was done using 10-fold cross validation. Figure 19 illustrates the resulting decision-making protocol, where only the following parameters are statistically relevant: ferritin, troponin, lipase dosage, and serum glucose. The accuracy obtained was 99.8552%, sensitivity of 0.999, specificity of 0.985, AUC of 0.985, and F-score of 0.999. We had 5 false positives and 4 false negatives.

**Fig. 19:**
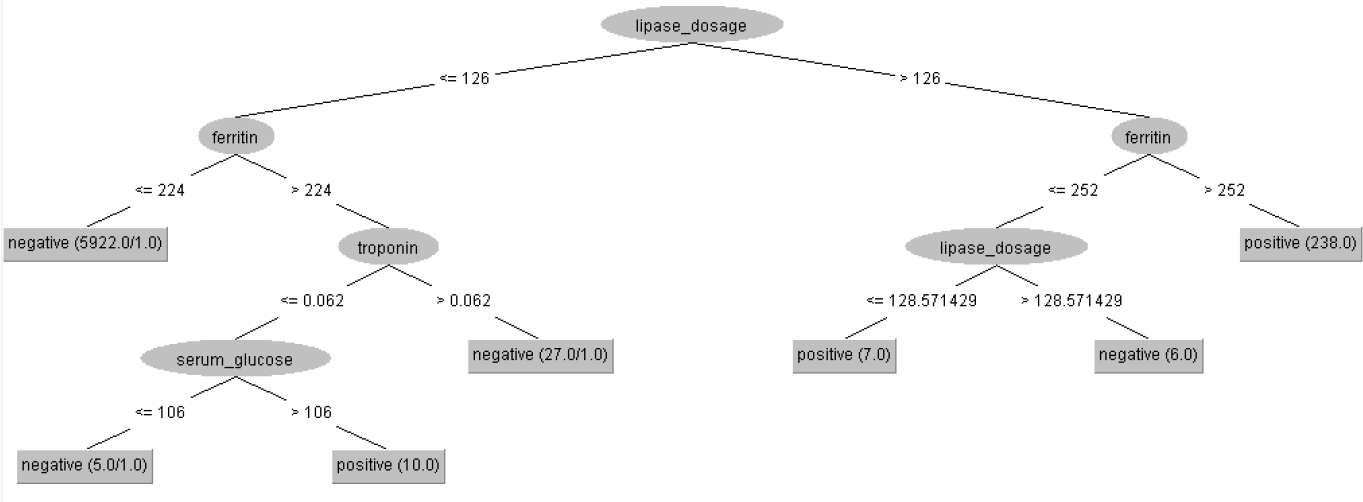
Clinical decision protocol based on J48 decision tree trained on the 7-feature dataset. Only the following were statistically relevant: lipase dosage, ferritin, troponin, and serum glucose. Training was done using 10-fold cross validation. Lipase dosage was dominant in this decision protocol. The accuracy was 99.8552%, sensitivity of 0.999, specificity of 0.985, AUC of 0.985, and F-score of 0.999. We had 5 false positives and 4 false negatives.

By removing the partial thromboplastin time and lipase dosage, we can assess the importance of the other 6 hematological parameters. We train a J48 decision tree again using 10-fold cross validation. The results are shown in Figure 20. The hematological parameters considered most relevant this time were: ferritin, troponin, D-dimer, and serum glucose. The accuracy obtained was 99.8713%, with a sensitivity of 0.999, specificity of 0.981, F-score of 0.999, and AUC of 0.990. We get 3 false positives and 5 false negatives.

**Fig. 20:**
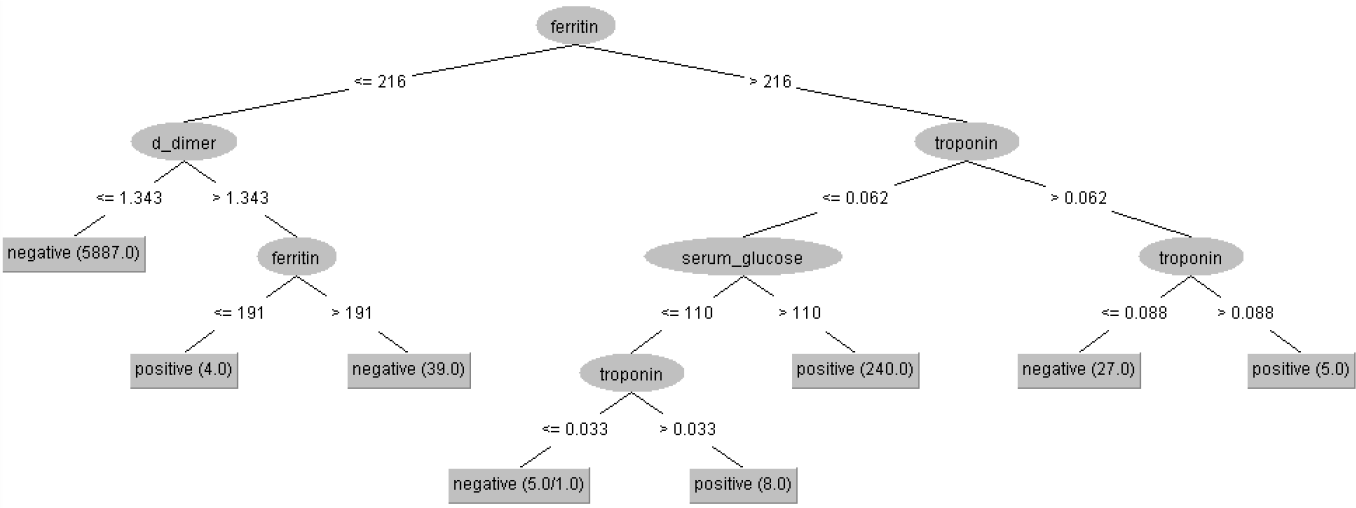
Clinical decision protocol based on J48 decision tree trained on the 6-feature dataset. Only the following were statistically relevant: ferritin, troponin, D-dimer, and serum glucose. Training was done using 10-fold cross validation. The accuracy obtained was 99.8713%, with a sensitivity of 0.999, specificity of 0.981, F-score of 0.999, and AUC of 0.990. We get 3 false positives and 5 false negatives.

We retrained a J48 decision tree removing the partial thromboplastin time, ferritin, and lipase dosage from the database. The training was done using 10-fold cross validation. The decision protocol is shown in Figure 21. All 5 remaining hematological parameters were considered relevant in the diagnostic decision: serum glucose, D-dimer, troponin, lactic dehydrogenase, and indirect bilirubin. The accuracy obtained was 99.7908%, with a sensitivity of 0.998, specificity of 0.955, AUC of 0.982, and an F-score of 0.998. We got 1 false positive and 12 false negatives.

**Fig. 21:**
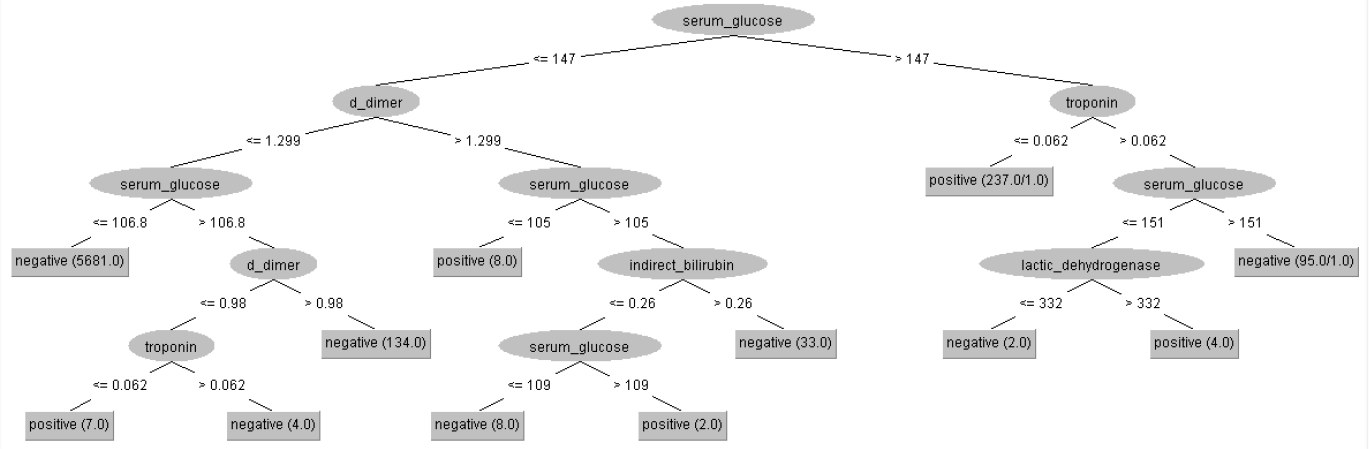
Clinical decision protocol based on J48 decision tree trained on the 5-feature dataset. All remaining features were statistically relevant: serum glucose, D-dimer, troponin, lactic dehydrogenase, and indirect bilirubin. Training was done using 10-fold cross validation. The accuracy obtained was 99.7908%, with a sensitivity of 0.998, specificity of 0.955, AUC of 0.982, and an F-score of 0.998. We got 1 false positive and 12 false negatives.

### 3.3 Heg.IA Web Application

After selecting the best classifier, the Heg.IA web system was developed. It can be accessed through the link: https://hegia.ufpe.br/welcome. Its front-end was developed using the library React.js. This library is based on pure JavaScript. It is open source and used to create user interfaces, more specifically, single page application (SPA) web platforms. As for data access and manipulation of application state, we used the Redux-Saga structure, a powerful tool that allows us to manage masterfully asynchronous queries, receiving API data, and trigger actions to the application of state safely and easily to maintain. Furthermore, our back-end was developed in Python (version 3.7.7). Only the Random Forest classifier was implemented in this final solution. Figure 22 illustrates how Random Forest was be applied as a classifier for the Covid-19 rapid diagnosis proposal we suggest in this work.

**Fig. 22:**
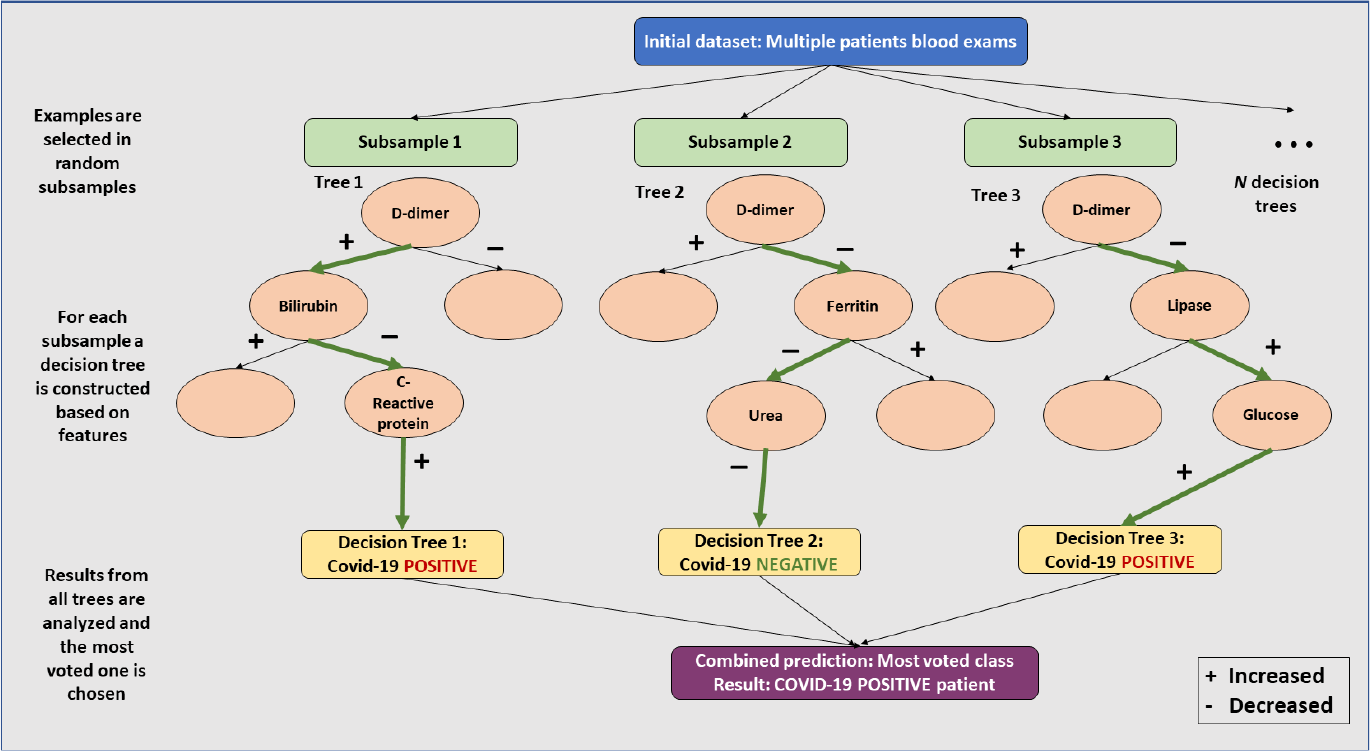
Random Forest classifier architecture adapted to Covid-19 rapid diagnosis as implemented in Heg.IA web proposal

On the initial screen, as shown on Figure 23, it is possible to visualize a brief description of the intelligent system, as well as the supporters of this initiative: The Federal University of Pernambuco (UFPE) and the Department of Biomedical Engineering at UFPE. To get to know the members of the project’s development team and their respective functions, it is possible to access the ‘About’ option on the top menu of the screen. The options ‘Login’ and ‘Consult’ are also available. For the ‘Login’ option, health professionals, especially medical laboratory professionals and nurses, will be able to access their private account or register a new account, in cases of first access. In the Consult option, it is possible to view the report with the diagnosis for a specific patient, as long as the user has the patient’s personal locator.

**Fig. 23:**
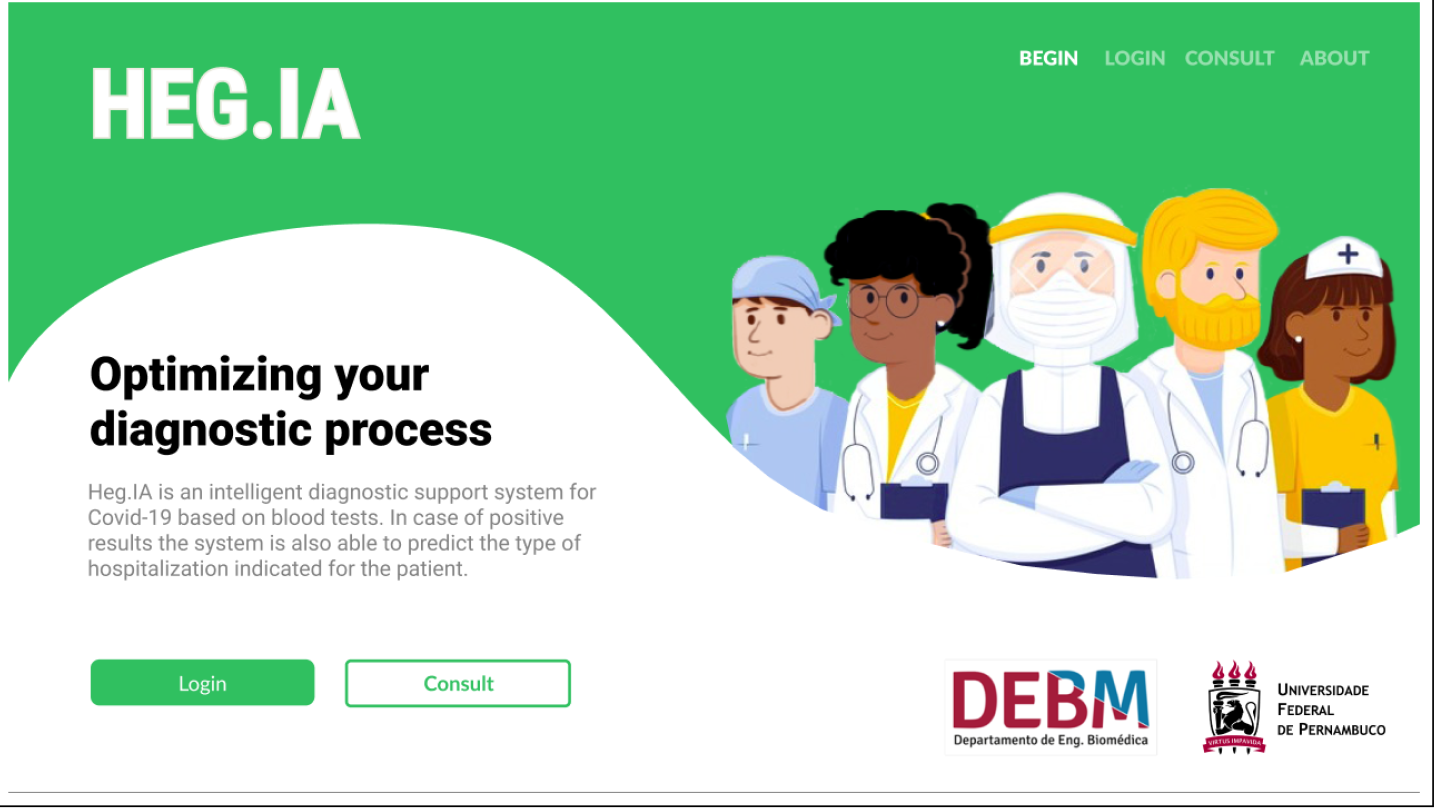
Heg.IA homepage: an intelligent web-based system for diagnosing Covid-19 through blood tests. On this initial screen, it is possible for the user (nurses and medical laboratory professionals) to login in his personal account. There are also the ‘Consult’ and ‘About’ options, available for all users, including patients and physicians. They allow the user to visualize the diagnostic report, and to get to know the involved team in this project, respectively.

After logging into the system, the user can register a patient or view the complete history of registered patients. In the case of a new registration, personal information such as full name, ID, date of birth, telephone, sex and full home address will be requested (Figure 24). In the following, the user will be directed to the screen shown in the Figure 25. In this screen, the results of the Complete blood count (CBC) with differential must be entered. The units and reference values are available next to each of the hematological parameters. After filling in the CBC, the user will be directed to the screens for the other blood tests and arterial blood gas tests, as shown in the Figures 26 and 27. Thus, the list of tests required to make the predictions will be complete. The user can then check the parameters entered in the screen ‘Let’s check it out?’, as shown in the Figure 28. If he realizes that he made a typo, he can go back to the previous steps and correct it.

**Fig. 24:**
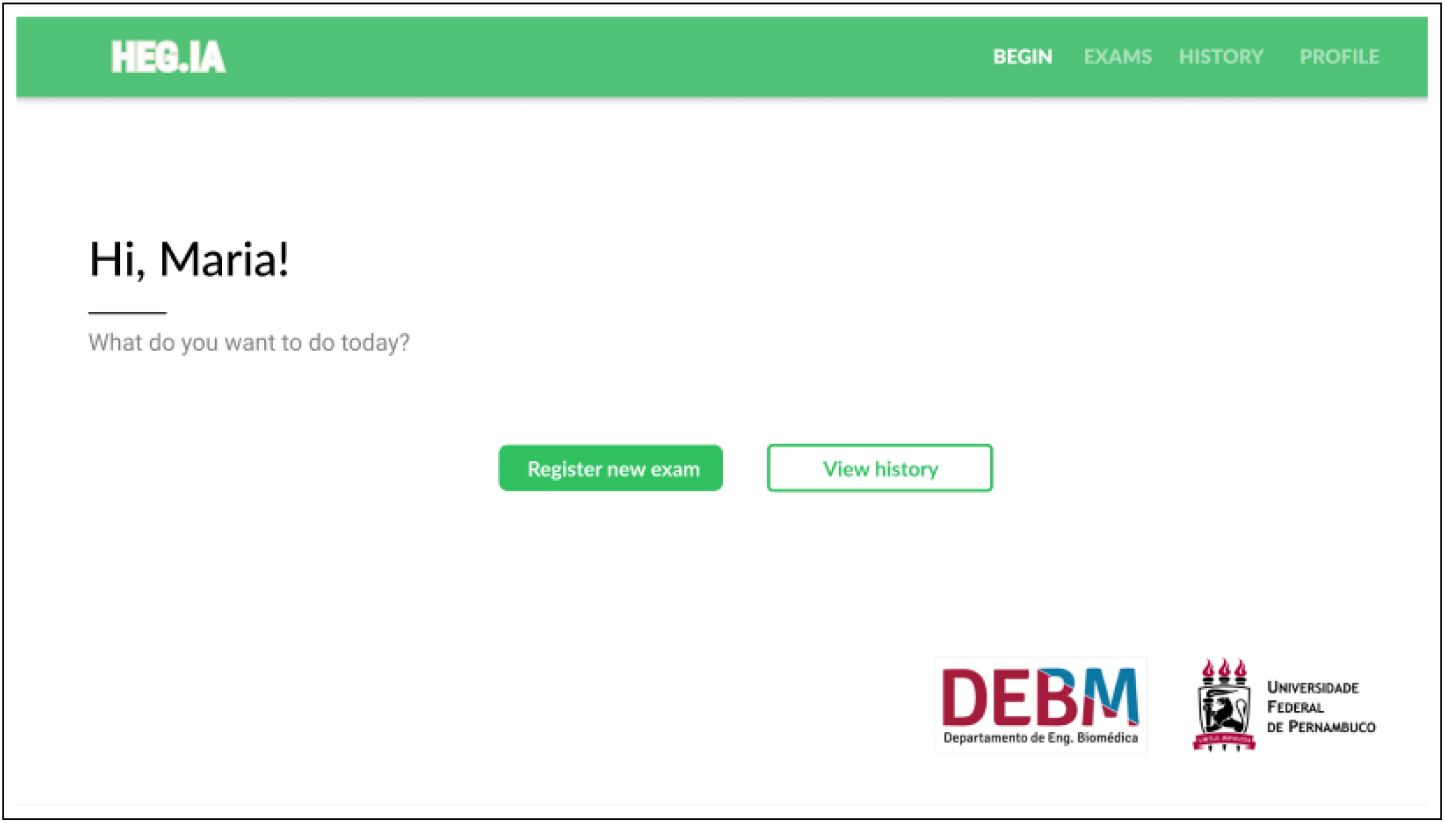
On this screen, the logged in user will be able to register new patients or access the complete history of patients already registered.

**Fig. 25:**
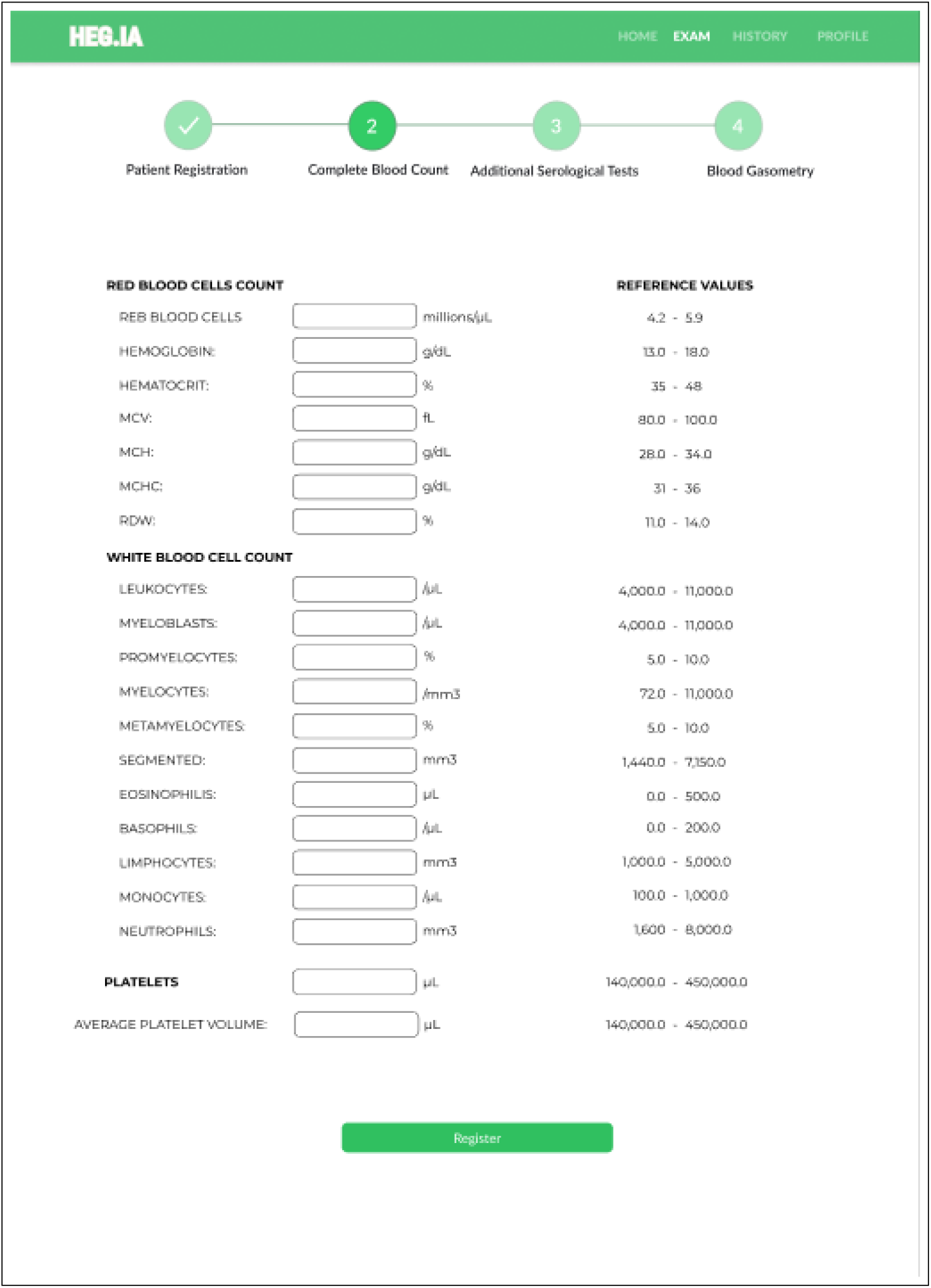
Complete Blood Count screen: After the patient’s registration, the results of the patient’s Complete Blood Count with differential can be inserted. The units and reference values can be viewed next to each hematological parameter.

**Fig. 26:**
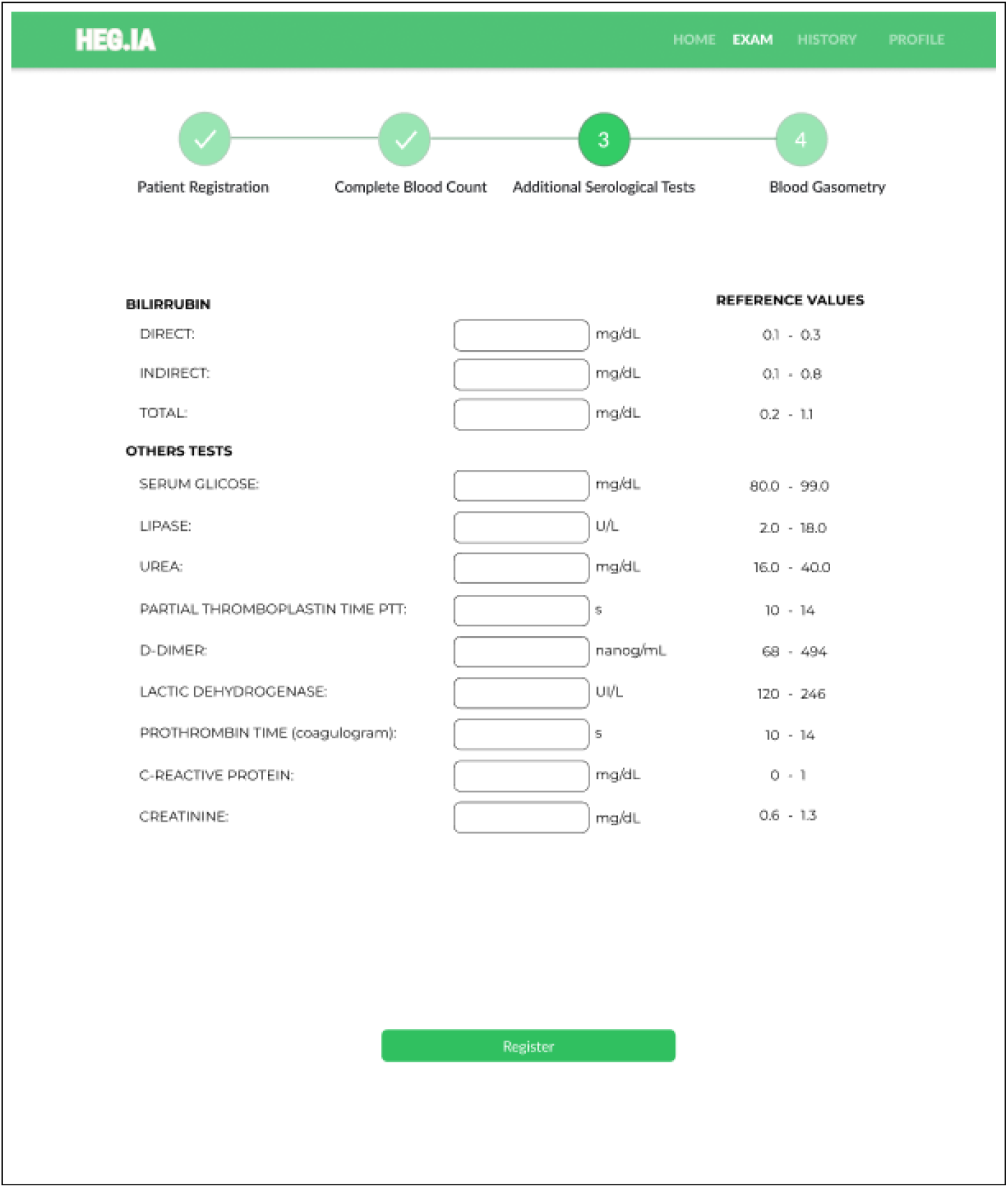
Additional Serological Tests Screen: Additional tests, such as total, direct and indirect bilirubin, can be inserted in this screen. In addition, serum glicose, dosage lipase, urea, PTT, D-dimer, lactic dehydrogenase, prothrombin time, CRP, and creatinine results can also be included here. The units and reference values can be viewed next to each hematological parameter.

**Fig. 27:**
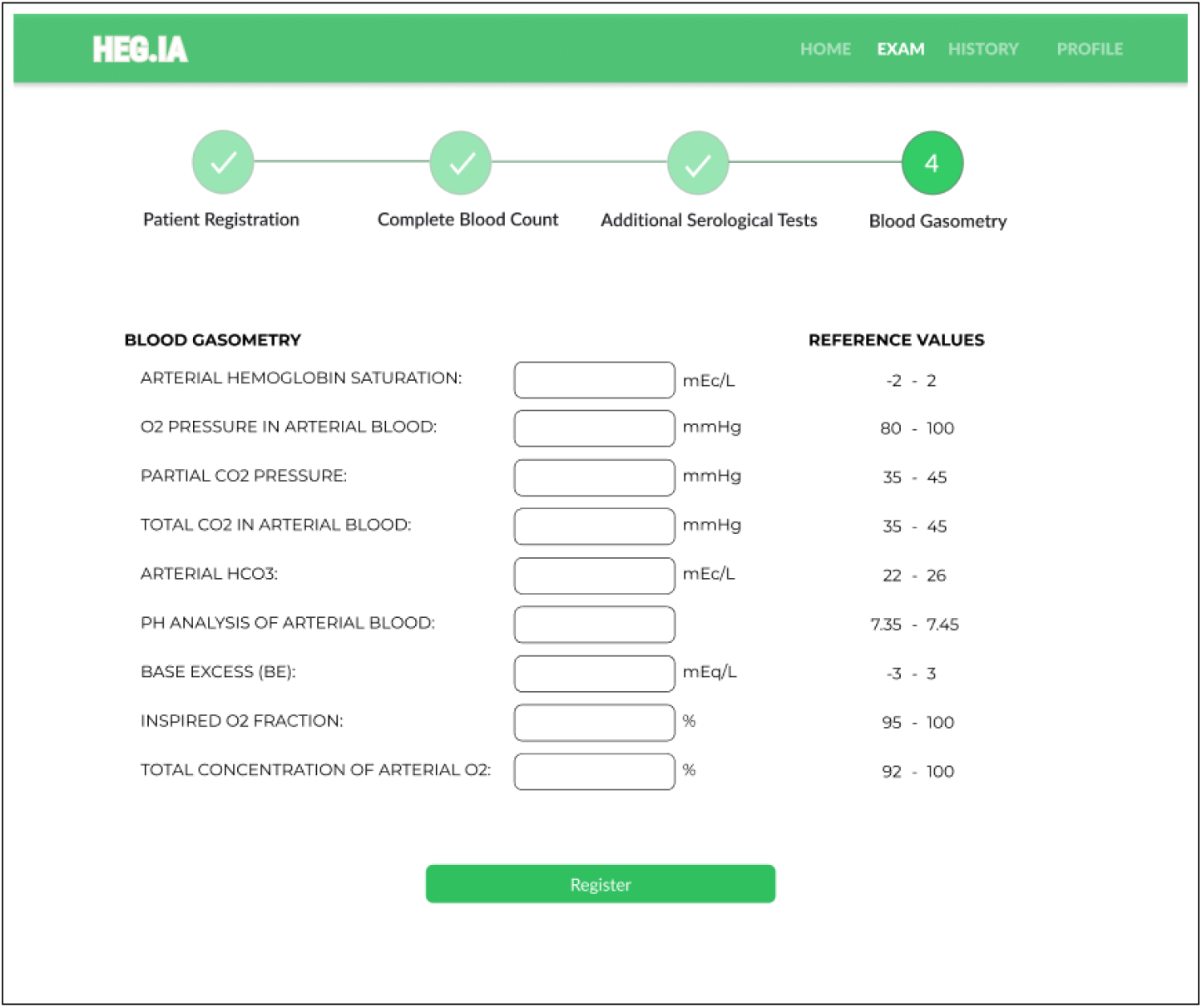
Blood Gasometry Screen: Arterial gasometry can be inserted in this screen, finalizing the list of necessary exams. The units and reference values for each parameter are available.

**Fig. 28:**
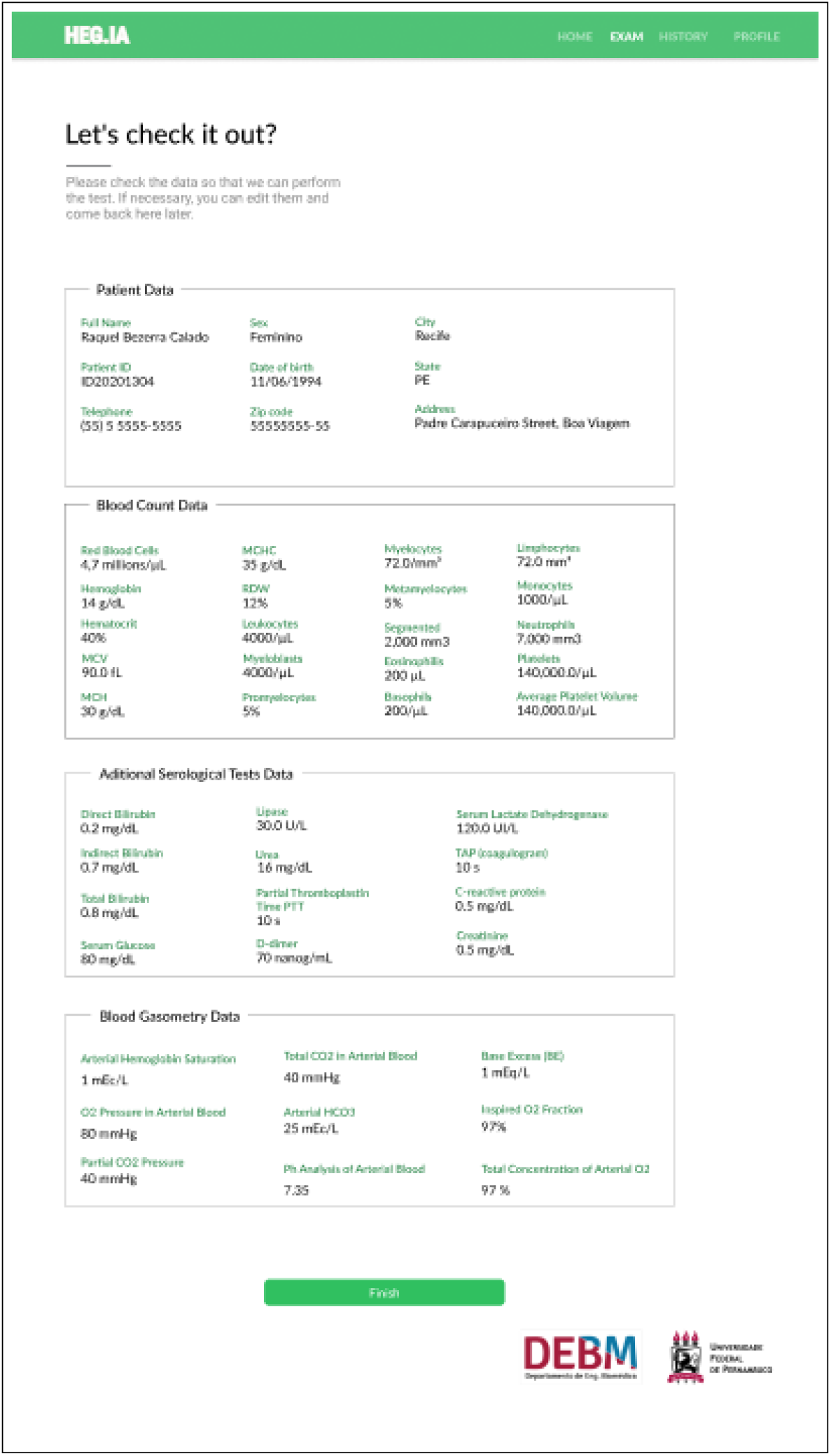
In the screen, it is possible for the user to check the patient’s personal information and the values of the hematological parameters inserted. If there is a typo, the user can return to the previous screens to correct it. If they are correct, it is possible to select the “Finish” option in order to access the report.

Finally, the report will be available immediately, similarly to that shown in the Figure 29. The diagnostic report will indicate the positive or negative diagnosis for Covid-19. Hospitalization predictions are also reported, indicating the best type of hospitalization for the patient: regular ward, semi-ICU or ICU. Information on accuracy, kappa index, sensitivity and specificity of the determination of each of these scenarios are also available, in order to assist the physician’s decision making. In addition to viewing the report, it is also possible to print it.

**Fig. 29:**
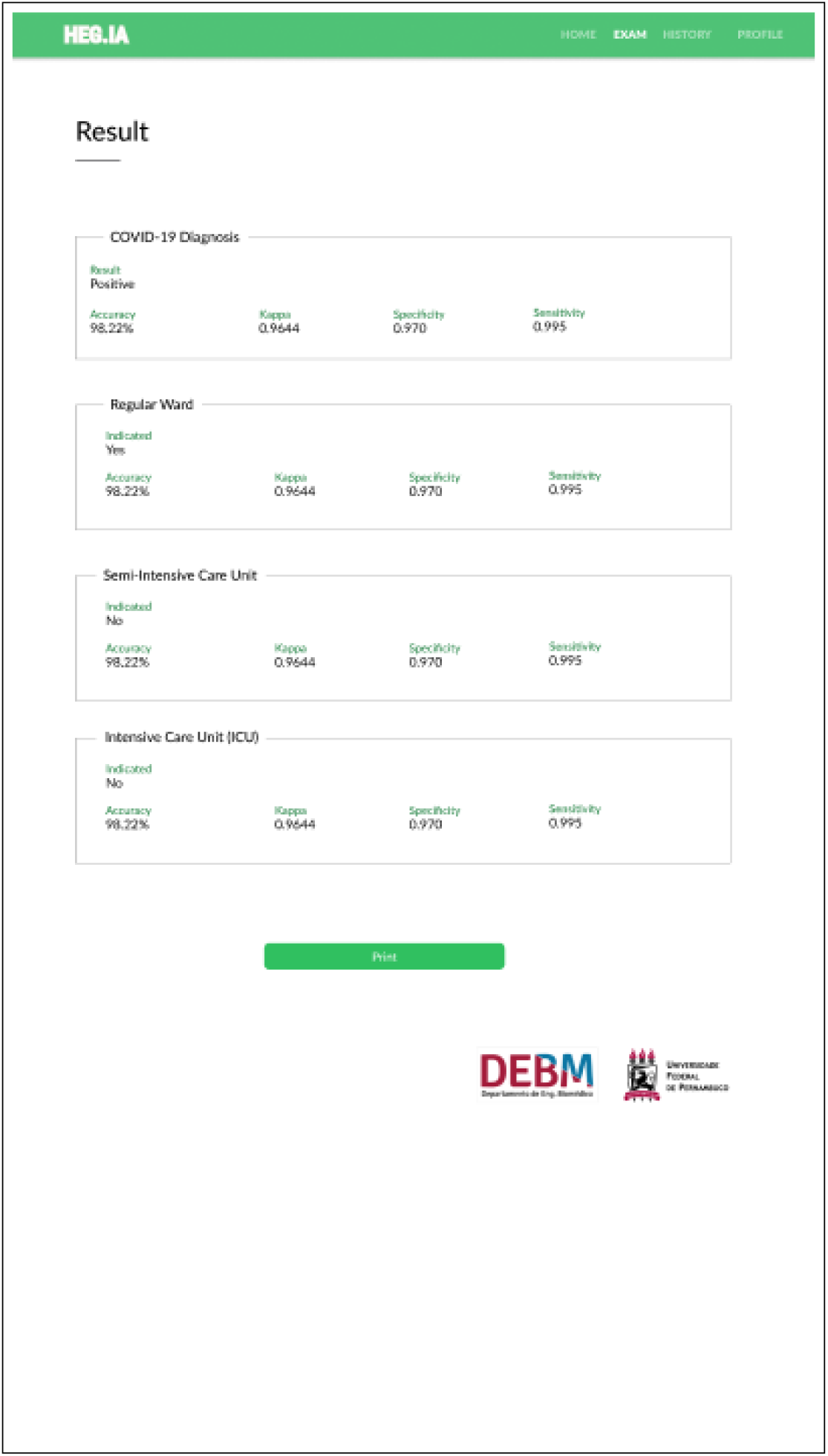
Results screen: In this screen it is possible to view the patient’s diagnostic report. In the report, the diagnosis for Covid-19 is available, as well as the hospitalization predictions, indicating whether the patient should be admitted to the regular ward, semi-intensive care unit, or to the ICU. Information on accuracy, kappa index, sensitivity and specificity of the determination of each of these scenarios are also available, in order to assist the physician’s decision making. In addition to viewing the report, it is also possible to print it.

## 4 Discussion

### 4.1 Evaluation of classifiers to support the diagnosis of Covid-19

When we analyze the results of the box plots in Figure 6 for the MLP neural network and the Bayesian methods, we can see that the accuracy results are quite accurate, with results concentrated approximately between 99.55% and 100%. Results with MLP are statistically equivalent. These results, in turn, are also statistically similar to the result of the Naive Bayes classifier. Bayes Net results are different and statistically superior to these, with a concentration close to 100% accuracy, first quartile starting at 99.53% and one point out at 99.52%. Looking at Table 4, we can see that Bayes Net’s performance is optimal (nearly 1.0000 or 100.00%) for all metrics, with standard deviation less than 0.0001. The results for Naive Bayes and the three MLP configurations are also considered close to optimal, with the sample means of all metrics greater than 99%, but with sample standard deviations much higher than those of Bayes Net, at least 100 times greater for accuracy, kappa and specificity.

Analyzing the results for the SVMs (Figures 7, 8, 9, and 10), considering C as 0.01 (Figure 7), we notice that the accuracy values are all greater than 95%. The results for the RBF kernel are statistically similar and stable, concentrated around 95.5%. For the linear kernel, the accuracy results vary around 96.2% with less than 1% variation. The best results were obtained for polynomial kernels with degrees 2, 3, 4 and 5, with accuracies concentrated in the third quartile, between 99.8% and 100%. According to Figure 8, for C of 0.1, all configurations have accuracies concentrated in the third quartile, between 99.8% and 100%, except for the configuration with RBF kernel and *γ* of 0.01, where the accuracy appears concentrated around 95.8%. For C of 1.00, as shown in Figure 9, all accuracy values are greater than 99.65%. The best settings are observed for linear, 2-degree polynomial, and RBF kernels with *γ* of 0.01 and 0.25: the first quartile is between 99.84% and 100%, while the other results are concentrated in 100% accuracy. For C of 10.00, the best accuracy results are those obtained with the RBF kernel (see Figure 10): the first quartile is between 99.84% and 100%, while the other results are concentrated in 100% of accuracy. Analyzing the kappa results in Table 4, we can see that the SMVs can only deal well with the high base unbalance for C equal to 1.00 or 10.00.

Figure 11 shows the box plots of the accuracy results for the decision tree-based methods. We investigate the J48 and Random Tree simple decision trees. We also evaluated the use of Random Forests with 10, 20, 30, 40, 50, 60, 70, 80, 90, and 100 trees. The results with Random Tree show that the absolute majority of the accuracy results for this classification architecture are concentrated between 99.54% and 100% (second, third and fourth quartiles). The first quartile is located between 99.50% and 99.54%, with a point out at 98.54%. This shows that Random Tree decision trees are very suitable for this problem, as the worst accuracy result is 98.54% while the absolute majority of the results are between 99.54% and 100%. Table 4 shows that these accuracy results for Random Tree really correspond to good classification results and that there was no bias due to base imbalance: the kappa index was 0.98 *±* 0.03, the sensitivity was 0.999 *±*0.001, the specificity was 0.98 *±* 0.03, the AUC was 0.99 *±* 0.02, and the F-score was 0.999 *±* 0.0001. Considering the accuracy, the results for the J48 tree and the Random Forests with 10, 20 and 30 trees are statistically similar: 75% of the results (second, third and fourth quartiles) are concentrated in exactly 100%, while the first quartile it is located between 99.56% and 100% (20% of the results for the Random Forest with 10 trees, 25% of the results for the other methods mentioned). Specifically for Random Forest with 10 trees, maximum 5% of the results are located at 99.50%. These results are considered very good, but the best results were those obtained with Random Forests with 40, 50, 60, 70, 80, 90, and 100 trees: 95% of accuracy results correspond to 100% accuracy, while at most 5% of the results correspond to 99.56% accuracy. Given that the best results were obtained with Random Forests and these classifiers are sufficiently robust to noisy data (possible typing errors of hematological parameters and missing data that need to be statistically estimated), we decided to adopt a Random Forest with 100 trees to implement the system. support for Heg.IA web diagnostics. Another advantage of Random Forests and decision trees in general is their low computational cost of processing, although the memory consumption can be considerable. Also, sorting with decision trees does not require input attributes to be rescheduled through normalization or standardization, a necessary preprocessing stage when we have to work with neural networks, support vector machines or Bayesian methods.

These results show that the 43 attributes (age and 42 hematological attributes) adopted in this work can be used to diagnose patients with symptoms of Covid-19 with a precision close to that obtained with RT-PCR, as shown in Table 4, for the results with Random Forests. Therefore, the Heg.IA web system can be an important aid for the development of a rapid protocol for the clinical diagnosis of Covid-19, providing more certainty for clinical practice. These results also show that these 42 hematological parameters, given the high accuracy, sensitivity, specificity and AUC values obtained, comparable to those obtained with RT-PCR, can be very important for monitoring Covid-19-positive patients.

The feature selection method based on the PSO algorithm returned the following eight attributes that were statistically most relevant for classification: serum glucose, lactic dehydrogenase, lipase dosage, partial thromboplastin time, troponin, ferritin, D-dimer, and indirect bilirubin. Figures 12, 13, 14, 15, 16, and 17 present the accuracy behavior according to the set of classifiers we adopted herein this work. Table 5 presents detailed results as sample average and standard deviation regarding accuracy, sensitivity, specificity, AUC, and F-score.

Analyzing the box plots in Figure 12, we can see that Bayesian methods and MLP networks returned very good results. For the Naive Bayes classifier, 75% of the experiments returned the maximum accuracy, 100% (second, third and fourth quartiles), while at least 20% of the experiments were located in accuracies between 99.56% and 100%, with a maximum of 5% of the 300 experiments returning 98.25%. The MLP settings had statistically similar accuracy results, with at least 95% of the experiments returning accuracies between 99.56% and 100%, and at most 5% of the experiments with an accuracy of 99.55% (20 and 50 hidden neurons) or 99% (100 neurons). The best results were obtained with the Bayes Net classifier, with all accuracy results concentrated at 100%. These results are confirmed by Table 5, which shows the sample means of the quality metrics reaching a maximum, with sample standard deviations less than 0.0001.

Figures 13, 14, 15, and 16 illustrate the box plots of the accuracy results for the use of SVMs, varying the kernels (linear, polynomial from degrees 2 to 5, and RBF), for C from 0.01, 0, 1, 1.0 and 10.0. For C equal to 0.01, the best results were obtained with linear kernel, with most experiments returning accuracies between 96% and 96.5%, with up to 5% of experiments reaching 100% (see Figure 13). For the grade 2 polynomial kernel, the experiment results were concentrated between 95.6% and 96%, with up to 5% of the experiments reaching 100% (see Figure 13). The results with polynomial kernel of grades 3, 4 and 5, and RBF are statistically similar, with 75% of the experiments resulting in accuracies of 95.8%, at least 20% of the experiments with an accuracy between 95.8% and 96%, and up to 5% of the experiments reaching a maximum of 100% accuracy (see Figure 13). For experiments with C equal to 0.1, the results for the linear kernels, polynomial of degrees 2 and 3, and RBF with *γ* of 0.25 and 0.5, were significantly improved: 75% of the experiments achieved the maximum accuracy of 100% (second, third and fourth quartiles), at least 20% of the experiments were between 99.8% and 100%, while up to 5% of the experiments returned an accuracy equal to 99.6% (see Figure 14). For C equal to 1.0, all results were considerably improved for all settings: 75% of the experiments reached 100% accuracy (second, third and fourth quartiles); 25% of the linear kernel and RBF experiments with *γ* of 0.01 were between 99.84% and 100% accuracy (first quartile); at least 20% of the experiments with grade 2 polynomial kernel and with RBF with *γ* of 0.25 and 0.5 were between 99.84% and 100% accuracy (first quartile); the results for polynomial kernel of degrees 3, 4 and 5 are statistically equivalent: at least 50% of the experiments reached accuracies between 99.85% and 100% (first and second quartiles), another 50% of the experiments reached the maximum accuracy, and up to 5% of the experiments returned an accuracy of 99.68%; the results for linear kernel, degree 2 polynomial, and RBF are statistically similar (see Figure 15). For C equal to 10.00, there was no noticeable improvement compared to C equal to 1.00 (see Figure 16). Table 5 shows that, in this database with the 8 statistically most relevant hematological parameters, all tested SVM configurations were robust to high database imbalance only for C equal to 1.0 and 10.0, according to values of specificity and AUC, with grade 4 and 5 polynomial kernels being the least robust configurations.

Figure 17 shows the box plots of the accuracy results for the decision tree-based methods. We investigate the J48 and Random Tree simple decision trees. We also evaluated the use of Random Forests with 10, 20, 30, 40, 50, 60, 70, 80, 90, and 100 trees. The results for the Random Tree are considered quite good: at least 50% of the experiments returned accuracies between 99.84% and 100% (first and second quartile), 50% of the experiments reached 100% accuracy (third and fourth quartile), and at most 5% of the experiments resulted in 99.52% accuracy. However, the best results were achieved with the other configurations, which were statistically equivalent: 75% of the experiments reached the maximum accuracy (second, third and fourth quartile), while 25% of the experiments ranged between 99.84% and 100% (first quartile). Table 5 details that Random Forests and the J48 decision tree are well suited to support the diagnosis of Covid-19 using the eight selected hematological parameters: accuracy of 99.98% *±* 0.05, sensitivity of 0.998 *±* 0.006, specificity of 0.9998 *±* 0.0005, maximum specificity and AUC with sample standard deviation less than 0.0001, and F-score of 0.9999 *±* 0.0003.

These results show that the eight hematological parameters chosen by the PSO algorithm are very important for the clinical diagnosis of Covid-19 in symptomatic patients, reaching results comparable to those obtained with RT-PCR, the gold standard method for the diagnosis of Covid-19. These results also show that these hematological parameters are strongly related to Covid-19 itself, and should not only be part of Covid-19 clinical diagnosis protocols, but also protocols for patient follow-up and assessment of patient progress and severity of disease, as the review of the state of the art demonstrates.

### 4.2 Humanly intelligible models to support the clinical diagnosis of Covid-19

Figure 18 shows that, for the symptomatic case records of the public health system in the municipality of Paudalho, Brazil, the most important factor for a positive diagnosis for Covid-19 is the low partial thromboplastin time (less than or equal to 22). This is indicative of the possibility of early blood clotting, a clinical occurrence strongly associated with thrombopenias, thrombosis and blood clot formation. Since all patient records used in this research correspond to symptomatic patients who sought medical care, it is reasonable to consider that all positive cases of covid-19 are moderate or severe. This raises the hypothesis that, for symptomatic Covid-19 patients in moderate or severe condition, treatments with anticoagulants such as low molecular weight heparin may be successful. In fact, the literature and clinical practice report that these treatments are already being used with relative success, but they are restricted to very severe cases, often in intubated patients. For this protocol, the accuracy was 99.98%, whilst the sensitivity, specificity, AUC, and F-score were maximum (approximately 1.00).

Figure 19 shows the decision protocol for clinical diagnosis generated automatically by decision tree J48 after excluding partial thromboplastin time. The protocol shows the importance of lipase dosing, ferritin, troponin, and serum glucose for the clinical diagnosis, monitoring, and evaluation of Covid-19. Most patients with Covid-19 (238 out of 258) had high levels of lipase dosage (greater than 126) and ferritin (greater than 252). These appear to be critically ill patients, as ferritin indicates that the patient is highly symptomatic, whereas high-dose lipase may indicate cases of pancreatitis. A few patients were positive for Covid-19 for low ferritin (less than 252) and lipase dosage between 126 and 129 (7 patients). Other 10 patients had low lipase dosage (less than 126), but high ferritin (greater than 224), low troponin (less than 0.062) and high serum glucose (greater than 106). This may indicate the influence of high glucose levels in the worsening of Covid-19 symptoms, but it may be associated with milder or moderate cases. However, to be more certain, it would be necessary to have access to the clinical histories of these patients, which was not possible in this study. For this protocol, accuracy was 99.8552%, and we got sensitivity of 0.999, specificity of 0.985, AUC of 0.985, and F-score of 0.999.

Figure 20 shows the decision protocol obtained by removing the partial thromboplastin time and lipase dosage. In this case, we can observe that most patients with Covid-19 had high ferritin (greater than 216), low troponin (less than 0.062), and high serum glucose (greater than 110). A few patients had different situations: 5 patients with Covid-19 had high ferritin (greater than 216) and high troponin (greater than 0.088); 8 patients had high ferritin (greater than 216), low serum glucose (less than 110), and troponin between 0.033 and 0.062. However, for 4 patients, D-dimer was high (greater than 1.343) and ferritin was low (lower than 191), which may indicate complicators due to previous heart disease, but these conditions would need to be verified in the patients’ clinical histories. For this protocol, we got accuracy of 99.8713%, sensitivity of 0.999, specificity of 0.981, F-score of 0.999, and AUC of 0.990.

Figure 21 shows the decision protocol obtained automatically by the J48 decision tree after removing the partial thromboplastin time, lipase dosage, and ferritin. For this protocol, most patients with Covid-19 (237) had high serum glucose (greater than 147) and low but positive troponin (less than 0.062). As this hematologic parameter is often negative for healthy people, this condition may indicate a potential for Covid-19 to negatively alter cardiac function, possibly inducing myocardial damage and, depending on the patient’s history, myocardial infarction. The other patients did not present a specific rule for the relationship between hematological parameters. Its conditions were expressed in many rules. Some patients appear to be heart disease (4 patients) because they have high troponin. These patients had high lactic dehydrogenase (greater than 332) and serum glucose between 147 and 151. One group of patients (8 patients) had high D-dimer (greater than 1,299) and low serum glucose (less than 105). Another group, with more specific conditions (7 patients), had low troponin (less than 0.062), low D-dimer (less than 0.98), and serum glucose between 106.8 and 147. Finally, only 2 patients had this condition: serum glucose between 109 and 147, low indirect bilirubin (less than 0.26), and high D-dimer (greater than 1.299). For this protocol, we have accuracy of 99.79%, sensitivity of 0.998, specificity of 0.955, AUC of 0.982, and F-score of 0.998.

## 5 Conclusion

The disease caused by the new type of coronavirus, the Covid-19, has posed a global public health challenge. Since this virus has a strong human-to-human transmission ability, it has already led to millions of infected people and thousands of deaths since the beginning of the outbreak in December 2019.

In this study, we developed a web system, Heg.IA, which seeks to optimize the diagnosis of Covid-19 by combining blood tests and artificial intelligence. From the system, a healthcare professional may have a diagnostic report after providing patient’s age and 42 hematological parameters from common blood tests. The system is able to indicate if the patient is infected with SARS-CoV-2 virus. The proposed system is based on decision trees and achieved great performance of accuracy, kappa statistic, sensitivity, precision, specificity and area under ROC for all tested scenarios. Considering SARS-CoV-2 detection, the system may play an important role as a highly efficient rapid test to aid at the clinical diagnosis as a way to provide rapid clinical diagnosis protocols.

The experimental results demonstrate that the eight hematological parameters selected by PSO, i.e. serum glucose, lactic dehydrogenase, lipase dosage, partial thromboplastin time, troponin, ferritin, D-dimer, and indirect bilirubin, are very significant for the clinical diagnosis of Covid-19 in symptomatic patients. Classification results are comparable to those obtained with RT-PCR. Consequently, serum glucose, lactic dehydrogenase, lipase dosage, partial thromboplastin time, troponin, ferritin, D-dimer, and indirect bilirubin are closely related to Covid-19 dynamics. The experimental results are suggestive for the use of these eight hematological parameters as part of Covid-19 clinical diagnosis protocols, as well as patient monitoring assessment of patient progress and severity of disease, confirming state-of-the-art results. These parameters may even be important for the construction of new specific treatments for Covid-19, which take into account the clinical and biochemical interpretation of these parameters.

The change in serum glucose by Covid-19 may explain the fact that diabetes is considered a comorbidity that can increase the chances of worsening the patient’s death by Covid-19 Indirect biblirubin is formed by the breakdown of red blood cells and is then transported to the liver. Bilirubin can be related to several complications in the blood, changes in heart function and liver problems. This, in turn, may be strongly related to how the chances of aggravation and death in patients with heart disease and liver disease may increase. Troponin alterations, also considered statistically relevant in this research, also show how patients with heart disease can have their chances of worsening the disease. D-dimer is one of the breakdown products of fibrin, a protein that is involved in clot formation. Therefore, when there are changes in the coagulation process, it is possible that there is a greater amount of circulating D-dimer. Lactic dehydrogenase is an enzyme present in all cells, but in greater concentration in the lungs, heart, skeletal muscle and liver. High levels of lactic dehydrogenase in patients with Covid-19 increase the chances of aggravation of the infection and, therefore, the probability of death. Lipase is an enzyme that is part of the digestive process. It works by breaking down fat molecules to be more easily absorbed by the intestine. The lipase test is a clinical analysis that helps identify changes in the levels of this enzyme in the individual’s body. High levels of lipase are associated with acute pancreatitis. Several studies show that Covid-19 also affects the pancreas in moderate and severe cases. State-of-the-art investigations show that the occurrence of pancreatitis at different levels is reasonably common in patients positive for Covid-19 and may be a highly important clinical occurrence in intubated patients.

Reduced partial thromboplastin time indicates the possibility of early blood clotting. It is strongly related to the occurrence of thrombosis, thrombopenias and other problems related to early coagulation, common to moderate and severe cases of Covid-19 and in post-Covid-19 complicators. Ferritin is a protein produced mainly by the liver, whose basic functions are: transporting iron and mediating the inflammation process. High ferritin can be a symptom of inflammation or infections, as it is an acute phase protein, which may be increased in the following situations: Covid-19, hemolytic anemia, megaloblastic anemia, alcoholic liver disease, Hodgkin’s lymphoma, myocardial infarction, leukemia, and hemochromatosis. Symptoms of excess ferritin are joint pain, tiredness, shortness of breath or abdominal pain, i.e. symptoms common to Covid-19. This information, combined with the results obtained with decision trees, suggest the clinical use of partial time thromboplastin and ferritin both for the rapid clinical diagnosis of symptomatic cases of Covid-19 without the aid of diagnostic support systems and for the assessment of severity of the disease and the follow-up of the patient’s clinical situation.

These results may also indicate the possibility of constructing specific treatments for moderate and severe cases of Covid-19, which can be based on therapies with anticoagulants, IL-6 blocking drugs. Another possibility may be to adapt therapies aimed at reducing ferritin for Covid-19 cases, respecting the degree of disease severity and patient limitations.

## Data Availability

Data will be available on demand.

## Acknowledgements

The authors are grateful to the Federal University of Pernambuco, Google Cloud COVID-19 Research Grant, and the Brazilian research agencies CAPES and CNPq, for the partial financial support of this research.

We thank the Mayor of the City of Paudalho, Mr. Marcelo Gouveia, for the collaboration and partnership with our research team. We are also grateful to the Health Department of the Municipality of Paudalho and all the medical, nurses and biomedical professionals of the Municipal Public Health System who collaborated with this research.

## Conflict of Interest

All authors declare they have no conflicts of interest.

## Compliance with Ethical Standards

This study was funded by the Federal University of Pernambuco, Google Cloud COVID-19 Research Grant, and the Brazilian research agencies CAPES and CNPq.

All procedures performed in studies involving human participants were in accordance with the ethical standards of the institutional and/or national research committee and with the 1964 Helsinki declaration and its later amendments or comparable ethical standards.

## Notes

### Competing Interest Statement

The authors have declared no competing interest.

## References

1. M. Abdar, N. Y. Yen, and J. C.-S. Hung. Improving the diagnosis of liver disease using multilayer perceptron neural network and boosted decision trees. Journal of Medical and Biological Engineering, 38(6):953–965, 2018.

2. J. Ahmadzadeh, K. Mobaraki, S. J. Mousavi, J. Aghazadeh-Attari, M. Mirza-Aghazadeh-Attari, and I. Mohebbi. The risk factors associated with mers-cov patient fatality: A global survey. Diagnostic Microbiology and Infectious Disease, 96 (3):114876, 2020.

3. E. I. Azhar, S. A. El-Kafrawy, S. A. Farraj, A. M. Hassan, M. S. Al-Saeed, A. M. Hashem, and T. A. Madani. Evidence for camel-to-human transmission of mers coronavirus. New England Journal of Medicine, 370(26):2499–2505, 2014.

4. N. Barakat, A. P. Bradley, and M. N. H. Barakat. Intelligible support vector machines for diagnosis of diabetes mellitus. IEEE transactions on information technology in biomedicine, 14(4):1114–1120, 2010.

5. V. A. d. F. Barbosa, J. C. Gomes, M. A. de Santana, E. d. A. Jeniffer, R. G. de Souza, R. E. de Souza, and W. P. dos Santos. Heg.IA: An intelligent system to support diagnosis of Covid-19 based on blood tests. Research on Biomedical Engineering, 2021: 1–18, 2021.

6. V. A. F. Barbosa, M. A. Santana, M. K. S. Andrade, R. C. F. Lima, and W. P. Santos. Deep-wavelet neural networks for breast cancer early diagnosis using mammary termographies. In H. Das, C. Pradhan, and N. Dey, editors, Deep Learning for Data Analytics: Foundations, Biomedical Applications, and Challenges. Academic Press, London, 1st edition, 2020.

7. S. Bataille, N. Pedinielli, and J.-P. Bergounioux. Could ferritin help the screening for COVID-19 in hemodialysis patients? Kidney international, 98(1):235–236, 2020.

8. C. T. Bauch and T. Oraby. Assessing the pandemic potential of mers-cov. The Lancet, 382(9893):662–664, 2013.

9. L. Borges. Medidas de acurácia diagnóstica na pesquisa cardiovascular. International Journal of Cardiovascular Sciences, 29(3):218–22, 2016.

10. B. E. Boser, I. M. Guyon, and V. N. Vapnik. A training algorithm for optimal margin classifiers. In Proceedings of the fifth annual workshop on Computational learning theory, pages 144–152, 1992.

11. R. R. Bouckaert. Bayesian network classifiers in weka for version 3-5-7. Artificial Intelligence Tools, 11(3):369–387, 2008.

12. D. Bratton and J. Kennedy. Defining a standard for particle swarm optimization. In 2007 IEEE Swarm Intelligence Symposium, pages 120–127. IEEE, 2007.

13. Brazilian Ministry of Health. Guidelines for the diagnosis and treatment of COVID-19. Brazilian Society of Clinical Analyzes, 2020. URL www.sbac.org.br/blog/2020/04/09/diretrizes-para-diagnostico-e-tratamento-da-covid-19/. Last accessed: 2020 June. 03.

14. L. Breiman. Random forests. Machine Learning, 45(1):5–32, 2001.

15. M. Cascella, M. Rajnik, A. Cuomo, S. C. Dulebohn, and R. Di Napoli. Features, evaluation and treatment coronavirus (covid-19). In StatPearls [Internet]. Stat Pearls Publishing, 2020.

16. S. Chatterjee, T. Sengupta, S. Majumder, and R. Majumder. COVID-19: a probable role of the anticoagulant ProteinS in managing COVID-19-associated coagulopathy. Aging (Albany NY), 12(16):15954, 2020.

17. J. Cheng and R. Greiner. Comparing bayesian network classifiers. In Proceedings of the Fifteenth conference on Uncertainty in artificial intelligence, pages 101–108. Morgan Kaufmann Publishers Inc., 1999.

18. J. Cheng and R. Greiner. Learning Bayesian belief network classifiers: Algorithms and System. Advances in Artificial Intelligence, 2056(1):141–151, 2001.

19. L. Cheng, H. Li, L. Li, C. Liu, S. Yan, H. Chen, and Y. Li. Ferritin in the coronavirus disease 2019 (covid-19): A systematic review and meta-analysis. Journal of Clinical Laboratory Analysis, 34(10):e23618, 2020.

20. D. K. Chu, L. L. Poon, M. M. Gomaa, M. M. Shehata, R. A. Perera, D. A. Zeid, A. S. El Rifay, L. Y. Siu, Y. Guan, R. J. Webby, et al. Mers coronaviruses in dromedary camels, egypt. Emerging Infectious Diseases, 20(6):1049, 2014.

21. M. Ciotti, S. Angeletti, M. Minieri, M. Giovannetti, D. Benvenuto, S. Pascarella, C. Sagnelli, M. Bianchi, S. Bernardini, and M. Ciccozzi. Covid-19 outbreak: an overview. Chemotherapy, 64(5-6):215–223, 2019.

22. E. Çomak, A. Arslan, and I. Türkoğlu. A decision support system based on support vector machines for diagnosis of the heart valve diseases. Computers in biology and Medicine, 37(1):21–27, 2007.

23. O. Commowick, A. Istace, M. Kain, B. Laurent, F. Leray, M. Simon, S. C. Pop, P. Girard, R. Ameli, J.-C. Ferré, et al. Objective evaluation of multiple sclerosis lesion segmentation using a data management and processing infrastructure. Scientific Reports, 8(1):1–17, 2018.

24. J. M. Connors and J. H. Levy. Covid-19 and its implications for thrombosis and anticoagulation. *Blood*, The Journal of the American Society of Hematology, 135(23): 2033–2040, 2020.

25. F. R. Cordeiro, W. P. Santos, and A. G. Silva-Filho. A semi-supervised fuzzy growcut algorithm to segment and classify regions of interest of mammographic images. Expert Systems with Applications, 65:116–126, 2016.

26. F. R. Cordeiro, W. P. d. Santos, and A. G. Silva-Filho. Analysis of supervised and semi-supervised growcut applied to segmentation of masses in mammography images. Computer Methods in Biomechanics and Biomedical Engineering: Imaging & Visualization, 5(4):297–315, 2017.

27. C. Cortes and V. Vapnik. Support-vector networks. Machine learning, 20(3):273–297, 1995.

28. V. A. Crooks, G. J. Andrews, and J. Pearce. Routledge Handbook of Health Geography. Routledge, 2018.

29. T. Cruz, T. Cruz, and W. Santos. Detection and classification of lesions in mammographies using neural networks and morphological wavelets. IEEE Latin America Transactions, 16(3):926–932, 2018.

30. S. Dahan, G. Segal, I. Katz, T. Hellou, M. Tietel, G. Bryk, H. Amital, Y. Shoenfeld, and A. Dagan. Ferritin as a Marker of Severity in COVID-19 Patients: A Fatal Correlation. The Israel Medical Association journal: IMAJ, 22(8):494–500, 2020.

31. C. L. de Lima, C. C. da Silva, A. C. G. da Silva, E. Luiz Silva, G. S. Marques, L. J. B. de Araújo, L. A. Albuquerque Júnior, S. B. J. de Souza, M. A. de Santana, J. C. Gomes, V. A. d. F. Barbosa, A. Musah, P. Kostkova, W. P. dos Santos, and A. G. da Silva Filho. Covid-sgis: A smart tool for dynamic monitoring and temporal forecasting of covid-19. Frontiers in Public Health, 8:761, 2020.

32. S. M. de Lima, A. G. da Silva-Filho, and W. P. dos Santos. A methodology for classification of lesions in mammographies using zernike moments, elm and svm neural networks in a multi-kernel approach. In 2014 IEEE International Conference on Systems, Man, and Cybernetics (SMC), pages 988–991. IEEE, 2014.

33. S. M. de Lima, A. G. da Silva-Filho, and W. P. dos Santos. Detection and classification of masses in mammographic images in a multi-kernel approach. Computer Methods and Programs in Biomedicine, 134:11–29, 2016.

34. J. de Vasconcelos, W. dos Santos, and R. de Lima. Analysis of methods of classification of breast thermographic images to determine their viability in the early breast cancer detection. IEEE Latin America Transactions, 16(6):1631–1637, 2018.

35. E. De Wit, N. Van Doremalen, D. Falzarano, and V. J. Munster. Sars and mers: recent insights into emerging coronaviruses. Nature Reviews Microbiology, 14(8):523, 2016.

36. M. Döhla, C. Boesecke, B. Schulte, C. Diegmann, E. Sib, E. Richter, M. Eschbach-Bludau, S. Aldabbagh, B. Marx, A.-M. Eis-Hübinger, et al. Rapid point-of-care testing for SARS-CoV-2 in a community screening setting shows low sensitivity. Public Health, 182:170–172, 2020.

37. B. E. Fan, V. C. L. Chong, S. S. W. Chan, G. H. Lim, K. G. E. Lim, G. B. Tan, S. S. Mucheli, P. Kuperan, and K. H. Ong. Hematologic parameters in patients with COVID-19 infection. American Journal of Hematology, 2020(04), 2020.

38. Y. Gao, T. Li, M. Han, X. Li, D. Wu, Y. Xu, Y. Zhu, Y. Liu, X. Wang, and L. Wang. Diagnostic utility of clinical laboratory data determinations for patients with the severe COVID-19. Journal of Medical Virology, 2020.

39. M. W. Gardner and S. Dorling. Artificial neural networks (the multilayer perceptron)—a review of applications in the atmospheric sciences. Atmospheric environment, 32(14-15):2627–2636, 1998.

40. P. Geurts, D. Ernst, and L. Wehenkel. Extremely randomized trees. Machine Learning, 63(1):3–42, 2006.

41. S. Gnanambal, M. Thangaraj, V. Meenatchi, and V. Gayathri. Classification algorithms with attribute selection: an evaluation study using weka. International Journal of Advanced Networking and Applications, 9(6):3640–3644, 2018.

42. J. C. Gomes, V. A. d. F. Barbosa, M. A. de Santana, J. Bandeira, M. J. S. Valença, R. E. de Souza, A. M. Ismael, and W. P. dos Santos. Ikonos: An intelligent tool to support diagnosis of covid-19 by texture analysis of x-ray images. Research on Biomedical Engineering, 2020:1–14, 2020.

43. J. C. Gomes, L. H. d. S. Silva, J. Ferreira, A. A. F. Junior, A. L. d. S. Rocha, L. Castro, N. R. C. da Silva, B. J. T. Fernandes, and W. P. dos Santos. Optimizing the molecular diagnosis of Covid-19 by combining RT-PCR and a pseudo-convolutional machine learning approach to characterize virus DNA sequences. bioRxiv, 2020.

44. J. Gómez-Pastora, M. Weigand, J. Kim, X. Wu, J. Strayer, A. F. Palmer, M. Zborowski, M. Yazer, and J. J. Chalmers. Hyperferritinemia in critically ill covid-19 patients–is ferritin the product of inflammation or a pathogenic mediator? Clinica Chimica Acta; International Journal of Clinical Chemistry, 2020(509):249–251, 2020.

45. G. Gunčar, M. Kukar, M. Notar, M. Brvar, P. Cernelc, M. Notar, and M. Notar. An application of machine learning to haematological diagnosis. Scientific Reports, 8(1): 1–12, 2018.

46. L. Guo, L. Ren, S. Yang, M. Xiao, D. Chang, F. Yang, C. S. Dela Cruz, Y. Wang, C. Wu, Y. Xiao, L. Zhang, L. Han, S. Dang, Y. Xu, Q.-W. Yang, S.-Y. Xu, H.-D. Zhu, Y.-C. Xu, Q. Jin, L. Sharma, L. Wang, and J. Wang. Profiling Early Humoral Response to Diagnose Novel Coronavirus Disease (COVID-19). Clinical Infectious Diseases, 2020(03), 2020.

47. D. J. Hand. Measuring classifier performance: a coherent alternative to the area under the roc curve. Machine Learning, 77(1):103–123, 2009.

48. T. T. Hasan, M. H. Jasim, and I. A. Hashim. Heart disease diagnosis system based on multi-layer perceptron neural network and support vector machine. Int J Curr Eng Technol, 77(55):2277–4106, 2017.

49. S. Haykin. Neural networks: principles and practice. Bookman, 11:900, 2001.

50. T. Hoffman, K. Nissen, J. Krambrich, B. Rönnberg, D. Akaberi, M. Esmaeilzadeh, E. Salaneck, J. Lindahl, and Å. Lundkvist. Evaluation of a covid-19 igm and igg rapid test; an efficient tool for assessment of past exposure to sars-cov-2. Infection Ecology & Epidemiology, 10(1):1754538, 2020.

51. T. Iba, J. H. Levy, M. Levi, and J. Thachil. Coagulopathy in covid-19. Journal of Thrombosis and Haemostasis, 18(9):2103–2109, 2020.

52. I. B. d. G. e. E. IBGE. Censo brasileiro de 2010, 2010.

53. Kaggle. Diagnosis of COVID-19 and its clinical spectrum. Kaggle, 2020. URL www.kaggle.com/einsteindata4u/covid19. Last accessed: 2020 Apr. 07.

54. K. Kappert, A. Jahic, and R. Tauber. Assessment of serum ferritin as a biomarker in covid-19: bystander or participant? insights by comparison with other infectious and non-infectious diseases. Biomarkers, pages 1–10, 2020.

55. J. Kennedy and R. Eberhart. Particle swarm optimization. In Proceedings of ICNN’95-International Conference on Neural Networks, volume 4, pages 1942–1948. IEEE, 1995.

56. K. F. Kernan and J. A. Carcillo. Hyperferritinemia and inflammation. International Immunology, 29(9):401–409, 2017.

57. F. Klok, M. Kruip, N. Van der Meer, M. Arbous, D. Gommers, K. Kant, F. Kaptein, J. van Paassen, M. Stals, M. Huisman, et al. Incidence of thrombotic complications in critically ill ICU patients with COVID-19. Thrombosis Research, 191:145–147, 2020.

58. S. B. Kotsiantis. Decision trees: a recent overview. Artificial Intelligence Review, 39 (4):261–283, 2013.

59. B. Lerner, M. Levinstein, B. Rosenberg, H. Guterman, L. Dinstein, and Y. Romem. Feature selection and chromosome classification using a multilayer perceptron neural network. In Proceedings of 1994 IEEE International Conference on Neural Networks (ICNN’94), volume 6, pages 3540–3545. IEEE, 1994.

60. Z. Li, Y. Yi, X. Luo, N. Xiong, Y. Liu, S. Li, R. Sun, Y. Wang, B. Hu, W. Chen, et al. Development and clinical application of a rapid igm-igg combined antibody test for sars-cov-2 infection diagnosis. Journal of medical virology, 2020.

61. D. Liao, F. Zhou, L. Luo, M. Xu, H. Wang, J. Xia, Y. Gao, L. Cai, Z. Wang, P. Yin, et al. Haematological characteristics and risk factors in the classification and prognosis evaluation of covid-19: a retrospective cohort study. The Lancet Haematology, 7(9): e671–e678, 2020.

62. S. Lima, W. Azevedo, F. Cordeiro, A. Silva-Filho, and W. Santos. Feature extraction employing fuzzy-morphological decomposition for detection and classification of mass on mammograms. In Conference proceedings:… Annual International Conference of the IEEE Engineering in Medicine and Biology Society. IEEE Engineering in Medicine and Biology Society. Annual Conference, volume 2015, pages 801–804, 2015.

63. Z. Lin, F. Long, Y. Yang, X. Chen, L. Xu, and M. Yang. Serum ferritin as an independent risk factor for severity in covid-19 patients. Journal of Infection, 81(4):647–679, 2020.

64. B. Liu, M. Li, Z. Zhou, X. Guan, and Y. Xiang. Can we use interleukin-6 (IL-6) blockade for coronavirus disease 2019 (COVID-19)-induced cytokine release syndrome (CRS)? Journal of Autoimmunity, page 102452, 2020.

65. J. Liu, S. Li, J. Liu, B. Liang, X. Wang, H. Wang, W. Li, Q. Tong, J. Yi, L. Zhao, et al. Longitudinal characteristics of lymphocyte responses and cytokine profiles in the peripheral blood of SARS-CoV-2 infected patients. EBioMedicine, page 102763, 2020.

66. T. Liu, J. Zhang, Y. Yang, H. Ma, Z. Li, J. Zhang, J. Cheng, X. Zhang, Y. Zhao, Z. Xia, et al. The role of interleukin-6 in monitoring severe case of coronavirus disease 2019. EMBO Molecular Medicine, 12(7):e12421, 2020.

67. Y. Liu, Y. Liu, B. Diao, F. Ren, Y. Wang, J. Ding, and Q. Huang. Diagnostic indexes of a rapid igg/igm combined antibody test for sars-cov-2. medRxiv, 2020, 2020.

68. H. Long, L. Nie, X. Xiang, H. Li, X. Zhang, X. Fu, H. Ren, W. Liu, Q. Wang, and Q. Wu. D-dimer and prothrombin time are the significant indicators of severe COVID-19 and poor prognosis. BioMed Research International, 2020, 2020.

69. Y. Luo, P. Szolovits, A. S. Dighe, and J. M. Baron. Using machine learning to predict laboratory test results. American Journal of Clinical Pathology, 145(6):778–788, 2016.

70. I. M. Mackay and K. E. Arden. Mers coronavirus: diagnostics, epidemiology and transmission. Virology Journal, 12(1):1–21, 2015.

71. F. S. Menezes-Rodrigues, J. G. Padrão Tavares, M. Pires de Oliveira, R. Guzella de Carvalho, P. Ruggero Errante, M. Omar Taha, D. José Fagundes, and A. Caricati-Neto. Anticoagulant and antiarrhythmic effects of heparin in the treatment of COVID-19 patients. Journal of Thrombosis and Haemostasis, 18(8):2073–2075, 2020.

72. P. Naraei, A. Abhari, and A. Sadeghian. Application of multilayer perceptron neural networks and support vector machines in classification of healthcare data. In 2016 Future Technologies Conference (FTC), pages 848–852. IEEE, 2016.

73. E. M. Negri, B. M. Piloto, L. K. Morinaga, C. V. P. Jardim, S. A. E.-D. Lamy, M. A. Ferreira, E. A. D’Amico, and D. Deheinzelin. Heparin therapy improving hypoxia in COVID-19 patients: a case series. Frontiers in Physiology, 11, 2020.

74. W. H. Organization. Advice on the use of point-of-care immunodiagnostic tests for COVID-19: scientific brief, 8 April 2020, 2020. URL https://www.who.int/news-room/commentaries/detail/advice-on-the-use-of-point-of-care-immunodiagnostic-tests-for-covid-19. Last accessed: March 26, 2021.

75. W. H. Organization. Weekly epidemiological update on COVID-19 – 16 March 2021, 2021. URL https://www.who.int/publications/m/item/weekly-epidemiological-update---16-march-2021. Last accessed: March 20, 2021.

76. M. Panigada, N. Bottino, P. Tagliabue, G. Grasselli, C. Novembrino, V. Chantarangkul, A. Pesenti, F. Peyvandi, and A. Tripodi. Hypercoagulability of COVID-19 patients in intensive care unit: a report of thromboelastography findings and other parameters of hemostasis. Journal of Thrombosis and Haemostasis, 18(7):1738–1742, 2020.

77. V. Pavoni, L. Gianesello, M. Pazzi, C. Stera, T. Meconi, and F. C. Frigieri. Evaluation of coagulation function by rotation thromboelastometry in critically ill patients with severe covid-19 pneumonia. Journal of Thrombosis and Thrombolysis, 50:281–286, 2020.

78. N. C. Peeri, N. Shrestha, M. S. Rahman, R. Zaki, Z. Tan, S. Bibi, M. Baghbanzadeh, N. Aghamohammadi, W. Zhang, and U. Haque. The SARS, MERS and novel coronavirus (COVID-19) epidemics, the newest and biggest global health threats: what lessons have we learned? International Journal of Epidemiology, 2020, 2020.

79. J. M. S. Pereira, M. A. Santana, R. C. F. Lima, S. M. L. Lima, and W. P. Santos. Method for classification of breast lesions in thermographic images using elm classifiers. In W. P. dos Santos, M. A. de Santana, and W. W. A. da Silva, editors, Understanding a Cancer Diagnosis, pages 117–132. Nova Science, New York, 1 edition, 2020.

80. J. M. S. Pereira, M. A. Santana, R. C. F. Lima, and W. P. Santos. Lesion detection in breast thermography using machine learning algorithms without previous segmentation. In W. P. dos Santos, M. A. de Santana, and W. W. A. da Silva, editors, Understanding a Cancer Diagnosis, pages 81–94. Nova Science, New York, 1 edition, 2020.

81. J. M. S. Pereira, M. A. Santana, W. W. A. Silva, R. C. F. Lima, S. M. L. Lima, and W. P. Santos. Dialectical optimization method as a feature selection tool for breast cancer diagnosis using thermographic images. In W. P. dos Santos, M. A. de Santana, and W. W. A. da Silva, editors, Understanding a Cancer Diagnosis, pages 95–118. Nova Science, New York, 1 edition, 2020.

82. S. L. Phung, A. Bouzerdoum, and D. Chai. Skin segmentation using color pixel classification: analysis and comparison. IEEE Transactions on Pattern Analysis and Machine Intelligence, 27(1):148–154, 2005.

83. V. Podgorelec, P. Kokol, B. Stiglic, and I. Rozman. Decision trees: an overview and their use in medicine. Journal of Medical Systems, 26(5):445–463, 2002.

84. R. Poli, J. Kennedy, and T. Blackwell. Particle swarm optimization. Swarm intelligence, 1(1):33–57, 2007.

85. L. Prior, D. Manley, and C. E. Sabel. Biosocial health geography: New ‘exposomic’ geographies of health and place. Progress in Human Geography, 43(3):531–552, 2019.

86. S. Raghu and N. Sriraam. Optimal configuration of multilayer perceptron neural network classifier for recognition of intracranial epileptic seizures. Expert Systems with Applications, 89:205–221, 2017.

87. A. L. Rodrigues, M. A. de Santana, W. W. Azevedo, R. S. Bezerra, V. A. Barbosa, R. C. de Lima, and W. P. dos Santos. Identification of mammary lesions in thermographic images: feature selection study using genetic algorithms and particle swarm optimization. Research on Biomedical Engineering, 35(3):213–222, 2019.

88. C. Rosário, G. Zandman-Goddard, E. G. Meyron-Holtz, D. P. D’Cruz, and Y. Shoenfeld. The hyperferritinemic syndrome: macrophage activation syndrome, Still’s disease, septic shock and catastrophic antiphospholipid syndrome. BMC Medicine, 11 (1):1–11, 2013.

89. H. A. Rothan and S. N. Byrareddy. The epidemiology and pathogenesis of coronavirus disease (covid-19) outbreak. Journal of Autoimmunity, 109:102433, 2020.

90. D. Sahu and T. Agrawal. Is it the Covid-19 happy hypoxia syndrome or the Covid-19 infodemic syndrome? Diabetes & Metabolic Syndrome, 14(5):1399, 2020.

91. M. A. Santana, J. M. S. Pereira, R. C. F. Lima, and W. P. Santos. Breast lesions classification in frontal thermographic images using intelligent systems and moments of haralick and zernike. In W. P. dos Santos, M. A. de Santana, and W. W. A. da Silva, editors, Understanding a Cancer Diagnosis, pages 65–80. Nova Science, New York, 1 edition, 2020.

92. M. A. d. Santana, J. M. S. Pereira, F. L. d. Silva, N. M. d. Lima, F. N. d. Sousa, G. M. S. d. Arruda, R. d. C. F. d. Lima, W. W. A. d. Silva, and W. P. d. Santos. Breast cancer diagnosis based on mammary thermography and extreme learning machines. Research on Biomedical Engineering, 34:45–53, 01 2018. ISSN 2446-4740.

93. C. Shi, W. Tingting, J.-P. Li, M. A. Sullivan, C. Wang, H. Wang, B. Deng, and Y. Zhang. Comprehensive Landscape of Heparin Therapy for Covid-19. Carbohydrate Polymers, page 117232, 2020.

94. W. W. A. Silva, M. A. Santana, A. G. Silva Filho, S. M. L. Lima, and W. P. Santos. Morphological extreme learning machines applied to the detection and classification of mammary lesions. In T. K. Gandhi, S. Bhattacharyya, S. De, D. Konar, and S. Dey, editors, Advanced Machine Vision Paradigms for Medical Image Analysis. Elsevier, London, 2020.

95. F. Soares, A. Villavicencio, F. S. Fogliatto, M. H. P. Rigatto, M. J. Anzanello, M. A. Idiart, and M. Stevenson. A novel specific artificial intelligence-based method to identify COVID-19 cases using simple blood exams. medRxiv, 2020.

96. L. Spiezia, A. Boscolo, F. Poletto, L. Cerruti, I. Tiberio, E. Campello, P. Navalesi, and P. Simioni. COVID-19-related severe hypercoagulability in patients admitted to intensive care unit for acute respiratory failure. Thrombosis and haemostasis, 120(6): 998, 2020.

97. T. Sun, J. Wang, X. Li, P. Lv, F. Liu, Y. Luo, Q. Gao, H. Zhu, and X. Guo. Comparative evaluation of support vector machines for computer aided diagnosis of lung cancer in ct based on a multi-dimensional data set. Computer methods and programs in biomedicine, 111(2):519–524, 2013.

98. L. Tan, Q. Wang, D. Zhang, J. Ding, Q. Huang, Y.-Q. Tang, Q. Wang, and H. Miao. Lymphopenia predicts disease severity of COVID-19: a descriptive and predictive study. Signal Transduction and Targeted Therapy, 5(1):1–3, 2020.

99. N. Tang, H. Bai, X. Chen, J. Gong, D. Li, and Z. Sun. Anticoagulant treatment is associated with decreased mortality in severe coronavirus disease 2019 patients with coagulopathy. Journal of Thrombosis and Haemostasis, 18(5):1094–1099, 2020.

100. L. Tanner, M. Schreiber, J. G. Low, A. Ong, T. Tolfvenstam, Y. L. Lai, L. C. Ng, Y. S. Leo, L. T. Puong, S. G. Vasudevan, et al. Decision tree algorithms predict the diagnosis and outcome of dengue fever in the early phase of illness. PLoS Neglected Tropical Diseases, 2(3), 2008.

101. S. Tural Onur, S. Altın, S. N. Sokucu, B. I. Fikri, T. Barça, E. Bolat, and M. Toptas. Could ferritin level be an indicator of COVID-19 disease mortality? Journal of Medical Virology, 93(3):1672–1677, 2021.

102. A. J. Turner, J. A. Hiscox, and N. M. Hooper. Ace2: from vasopeptidase to sars virus receptor. Trends in pharmacological sciences, 25(6):291–294, 2004.

103. F. Van den Bergh and A. P. Engelbrecht. A cooperative approach to particle swarm optimization. IEEE Transactions on Evolutionary Computation, 8(3):225–239, 2004.

104. M. Vargas-Vargas and C. Cortés-Rojo. Ferritin levels and covid-19. Revista Panamericana de Salud Pública, 44:e72, 2020.

105. M. Viecca, D. Radovanovic, G. B. Forleo, and P. Santus. Enhanced platelet inhibition treatment improves hypoxemia in patients with severe Covid-19 and hypercoagulability. A case control, proof of concept study. Pharmacological Research, 158:104950, 2020.

106. H. N. Vilar and R. M. de Medeiros. Índice de aridez na Zona da Mata no Estado de Pernambuco-Brasil. Journal of Environmental Analysis and Progress, 4(1):14–20, 2019.

107. D. Wang, B. Hu, C. Hu, F. Zhu, X. Liu, J. Zhang, B. Wang, H. Xiang, Z. Cheng, Y. Xiong, et al. Clinical characteristics of 138 hospitalized patients with 2019 novel coronavirus–infected pneumonia in Wuhan, China. Jama, 323(11):1061–1069, 2020.

108. J. Wang, N. Hajizadeh, E. E. Moore, R. C. McIntyre, P. K. Moore, L. A. Veress, M. B. Yaffe, H. B. Moore, and C. D. Barrett. Tissue plasminogen activator (tPA) treatment for Covid-19 associated acute respiratory distress syndrome (ARDS): a case series. Journal of Thrombosis and Haemostasis, 18(7):1752–1755, 2020.

109. X. Wang, J. Yang, X. Teng, W. Xia, and R. Jensen. Feature selection based on rough sets and particle swarm optimization. Pattern Recognition Letters, 28(4):459–471, 2007.

110. F. L. Wright, T. O. Vogler, E. E. Moore, H. B. Moore, M. V. Wohlauer, S. Urban, T. L. Nydam, P. K. Moore, and R. C. McIntyre Jr. Fibrinolysis shutdown correlation with thromboembolic events in severe COVID-19 infection. Journal of the American College of Surgeons, 231(2):193–203, 2020.

111. Y.-C. Wu, C.-S. Chen, and Y.-J. Chan. The outbreak of covid-19: an overview. Journal of the Chinese Medical Association, 83(3):217, 2020.

112. J. Yao, A. Dwyer, R. M. Summers, and D. J. Mollura. Computer-aided diagnosis of pulmonary infections using texture analysis and support vector machine classification. Academic radiology, 18(3):306–314, 2011.

113. Y.-Y. Zheng, Y.-T. Ma, J.-Y. Zhang, and X. Xie. COVID-19 and the cardiovascular system. Nature Reviews Cardiology, 17(5):259–260, 2020.

114. Z. Zhu, T. Cai, L. Fan, K. Lou, X. Hua, Z. Huang, and G. Gao. Clinical value of immune-inflammatory parameters to assess the severity of coronavirus disease 2019. International Journal of Infectious Diseases, 95:332–339, 2020.

